# Remdesivir for the treatment of hospitalised patients with COVID-19: final results from the DisCoVeRy randomised, controlled, open-label trial

**DOI:** 10.1101/2022.03.30.22273206

**Authors:** Florence Ader, Maude Bouscambert-Duchamp, Maya Hites, Nathan Peiffer-Smadja, Julien Poissy, Drifa Belhadi, Alpha Diallo, Christelle Delmas, Juliette Saillard, Aline Dechanet, Claire Fougerou, Minh-Patrick Lê, Gilles Peytavin, Noémie Mercier, Priyanka Velou, Sarah Tubiana, Xavier Lescure, Emmanuel Faure, Saad Nseir, Jean-Christophe Richard, Florent Wallet, François Goehringer, Benjamin Lefèvre, Antoine Kimmoun, François Raffi, Benjamin Gaborit, Jean Reignier, Jean-Philippe Lanoix, Claire Andrejak, Yoann Zerbib, Firouzé Bani-Sadr, Bruno Mourvilliers, François Danion, Yvon Ruch, Raphaël Clere-Jehl, Vincent Le Moing, Kada Klouche, Karine Lacombe, Guillaume Martin-Blondel, Fanny Vardon-Bounes, André Cabié, Jean-Marie Turmel, Lionel Piroth, Mathieu Blot, Élisabeth Botelho-Nevers, Amandine Gagneux-Brunon, Guillaume Thiery, François Bénézit, Rostane Gaci, Joy Mootien, Sébastien Gallien, Denis Garot, Kevin Bouiller, Loïc Epelboin, Stéphane Jauréguiberry, Alexandre Gaymard, Gil Verschelden, Sandra Braz, Joao Miguel Ferreira Ribeiro, Michael Joannidis, Thérèse Staub, Antoine Altdorfer, Richard Greil, Alexander Egle, Jérémie Guedj, Marion Noret, Roberto Roncon-Albuquerque, Jose-Artur Paiva, Bruno Lina, Dominique Costagliola, Yazdan Yazdanpanah, Charles Burdet, France Mentré

## Abstract

**Background:** The antiviral efficacy of remdesivir is still controversial. We aimed at evaluating its clinical effectiveness in hospitalised patients with COVID-19, with indication of oxygen and/or ventilator support. Following prior publication of preliminary results, here we present the final results after completion of data monitoring.

**Methods:** In this European multicentre, open-label, parallel-group, randomised, controlled trial (DisCoVeRy, NCT04315948; EudraCT2020-000936-23), participants were randomly allocated to receive usual standard of care (SoC) alone or in combination with remdesivir, lopinavir/ritonavir, lopinavir/ritonavir and IFN-β-1a, or hydroxychloroquine. Adult patients hospitalised with COVID-19 were eligible if they had clinical evidence of hypoxemic pneumonia, or required oxygen supplementation. Exclusion criteria included elevated liver enzyme, severe chronic kidney disease, any contra-indication to one of the studied treatments or their use in the 29 days before randomization, or use of ribavirin, as well as pregnancy or breast-feeding. Here, we report results for remdesivir + SoC *versus* SoC alone. Remdesivir was administered as 200 mg infusion on day 1, followed by once daily infusions of 100 mg up to 9 days, for a total duration of 10 days. It could be stopped after 5 days if the participant was discharged. Treatment assignation was performed *via* web-based block randomisation stratified on illness severity and administrative European region. The primary outcome was the clinical status at day 15 measured by the WHO 7-point ordinal scale, assessed in the intention-to-treat population.

**Findings:** Between March 22^nd^, 2020 and January 21^st^, 2021, 857 participants were randomised to one of the two arms in 5 European countries and 843 participants were included for the evaluation of remdesivir (control, n=423; remdesivir, n=420).

At day 15, the distribution of the WHO ordinal scale was as follow in the remdesivir and control groups, respectively: Not hospitalized, no limitations on activities: 62/420 (14.8%) and 72/423 (17.0%); Not hospitalized, limitation on activities: 126/420 (30%) and 135/423 (31.9%); Hospitalized, not requiring supplemental oxygen: 56/420 (13.3%) and 31/423 (7.3%); Hospitalized, requiring supplemental oxygen: 75/420 (17.9%) and 65/423 (15.4%); Hospitalized, on non-invasive ventilation or high flow oxygen devices: 16/420 (3.8%) and 16/423 (3.8%); Hospitalized, on invasive mechanical ventilation or ECMO: 64/420 (15.2%) and 80/423 (18.9%); Death: 21/420 (5%) and 24/423 (5.7%). The difference between treatment groups was not statistically significant (OR for remdesivir, 1.02, 95% CI, 0.62 to 1.70, P=0.93). There was no significant difference in the occurrence of Serious Adverse Events between treatment groups (remdesivir, n=147/410, 35.9%, *versus* control, n=138/423, 32.6%, p=0.29).

**Interpretation:** Remdesivir use for the treatment of hospitalised patients with COVID-19 was not associated with clinical improvement at day 15.

**Funding:** European Union Commission, French Ministry of Health, DIM One Health Île-de-France, REACTing, Fonds Erasme-COVID-ULB; Belgian Health Care Knowledge Centre (KCE), AGMT gGmbH, FEDER “European Regional Development Fund”, Portugal Ministry of Health, Portugal Agency for Clinical Research and Biomedical Innovation. Remdesivir was provided free of charge by Gilead.

## Introduction

The evaluation of repurposed drugs for severe acute respiratory syndrome coronavirus 2 (SARS-CoV-2)-associated coronavirus disease 2019 (COVID-19) has been conducted in several large-scale randomised clinical trials. Among them, the DisCoVeRy trial has investigated the efficacy and the safety of lopinavir/ritonavir, lopinavir/ritonavir plus IFN-β-1a, hydroxychloroquine, and remdesivir as compared to standard of care in adults hospitalised with COVID-19 (1). Results for lopinavir/ritonavir, lopinavir/ritonavir plus IFN-β-1a and hydroxychloroquine have been reported (2,3).

Remdesivir is a small molecule, formulated with sulfobutylether B-cyclodextrin sodium for injection, dialyzable, known to penetrate well into deep compartments and devoid of drug interactions *via* CYP450 (4,5). It is a nucleotide analogue prodrug, intracellularly metabolised to an analogue of adenosine triphosphate, which inhibits RNA polymerase activity in some pathogenic coronaviruses (6). It has shown evidence of antiviral activity against SARS-CoV-2 in preclinical models, both *in vitro* and *in vivo* (7,8), supporting its evaluation in COVID-19. In a randomised clinical trial in China including 237 COVID-19 patients, remdesivir was associated with a shorter time to clinical improvement in patients that started treatment within 10 days of symptom onset (9). In the Adaptive Covid-19 Treatment Trial (ACTT 1) including 1 062 patients, remdesivir was associated with a shorter time to recovery (10 *vs*. 15 days compared to placebo), but was not associated with a decrease in mortality (10), resulting in emergency use authorization (EUA). Similarly, the international Solidarity consortium trial sponsored by the World Health Organization (WHO), which included 2,750 patients on remdesivir found no benefit of remdesivir on in-hospital mortality across various health-care settings (11). Overall, these mixed results have not led so far to a consensus on the use of remdesivir for COVID-19 patients.

As an add-on trial, the DisCoVeRy trial shared with the WHO Solidarity consortium patients’ baseline characteristics, as well as the dates of hospital discharge and eventual need for oxygen therapy either through standard device, high flow device, non-invasive ventilation, mechanical ventilation or ECMO, or death (11). DisCoVeRy was designed to further document clinical outcomes, virological kinetics, treatment pharmacokinetics and related safety data. Preliminary results have been published previously (12), and we present here the final results, after completion of the data monitoring.

## Methods

### Study design

DisCoVeRy is a phase 3, open-label, adaptive, multicentre, randomised, controlled, superiority trial for evaluating the efficacy and safety of repurposed drugs in adults hospitalised for COVID-19 (1). It was conducted across 48 sites in 5 European countries (France, Belgium, Portugal, Austria, and Luxembourg). The trial was approved by the Ethics Committee (CPP Ile-de-France-III, approval #20.03.06.51744), and is sponsored by the Institut national de la santé et de la recherche médicale (Inserm, France); it was conducted in accordance with the Declaration of Helsinki. Written informed consent was obtained from all included participants (or their legal representatives if unable to consent). The present analysis is based on the protocol v11.0 of December 12^th^, 2020.

### Participants

Hospitalised participants ≥18 years of age with laboratory-confirmed SARS-CoV-2 infection and illness of any duration could be enrolled if they presented at least one of the following: clinical assessment (evidence of rales/crackles on exam) and SpO2 ≤ 94% on room air, or requirement of supplemental oxygen, high flow oxygen devices, non-invasive ventilation and/or mechanical ventilation. Women of childbearing potential must agree to use at least one primary form of contraception for the duration of the study. Written informed consent was obtained from all participants or from their legal representative if they were unable to provide consent. Participants were excluded from enrolment if they had liver enzymes (ALT/AST) > 5 times the upper limit of normal, a stage 4 severe chronic kidney disease or requiring dialysis (eGFR < 30 mL/min), or if a transfer to another hospital, which is not a study site within 72 hours was anticipated. Participants with contraindication to any study medication including allergy, treated with one of the evaluated antivirals in the past 29 days or who used ribavirin in the 29 days and/or concomitantly to randomisation were not eligible, as were pregnant or breast-feeding women.

The criterion of laboratory-confirmed SARS-CoV-2 infection was initially restricted to 72 hours prior to randomisation but was further extended to 9 days in the protocol v10.0 of October, 1^st^ 2020. Participants were included in 48 centres (France, n=39, Austria, n=3 Belgium, n=3, Portugal, n=2 and Luxembourg, n=1).

### Randomisation and masking

Participants were randomly assigned in a 1:1:1:1:1 ratio when 5 arms were initially implemented, and then in a 1:1 ratio to receive either standard of care (SoC, control arm) or SoC + remdesivir, once the other three treatment arms had been stopped for futility (2). Participants allocated to Standard of Care alone or in combination with remdesivir were recruited contemporaneously.

Randomisation was performed in the electronic Case Report Form to ensure appropriate allocation concealment and used computer-generated blocks of various sizes; it was stratified on severity of disease at inclusion and on European administrative region of inclusion. Disease was defined as moderate in participants not receiving supplemental oxygen or requiring supplemental oxygen through face mask or nasal prongs (*i*.*e*., ordinal scale value of 3 or 4); it was defined as severe in participants requiring non-invasive ventilation, high flow oxygen device, invasive mechanical ventilation or ECMO (*i*.*e*., ordinal scale value of 5 or 6). Allocated treatment was not masked to participants nor study investigator.

### Procedures

Remdesivir was administrated intravenously at a loading dose of 200 mg on day 1 followed by a 100 mg 1-hour infusion once-daily for a total duration of 10 days. Its cessation was allowed after 5 days if the participant was discharged from the hospital.

Corticosteroids and anticoagulants were added to the SoC on October 1^st^, 2020 (protocol v10.0). The suggested corticosteroids regimen was dexamethasone 6 mg once daily for 10 days or until discharge (13,14). In participants critically ill participants with Acute Respiratory Distress Syndrome (ARDS) requiring Intensive Care Unit (ICU), a standard ARDS dexamethasone regimen could be proposed at clinician’s discretion (dexamethasone 20 mg once daily for 5 days, followed by 10 mg once daily for 5 days) (15). Dosage regimens of anticoagulation was administered according to local protocols for venous thromboembolism prophylaxis and/or therapy (16,17). Other supportive treatments, such as immunomodulatory agents, were allowed in all arms and left to the investigator’s discretion. No participant received a SARS-CoV-2 vaccine during the trial.

Participants were assessed daily while hospitalised, and at days 3, 5, 8, 11, 15±2 and 29±3 if discharged. Clinical data, concomitant medications, adverse events (AEs), blood cell counts, serum creatinine and liver aminotransferases were collected. Nasopharyngeal (NP) swab specimens were collected for SARS-CoV-2 real-time (RT) PCR at days 3, 5, 8, 11, 15±2 and 29±3. Blood samples were collected at the discretion of the investigator in charge for measurement of remdesivir and its metabolite GS-441524 in plasma post-infusion (up to 30 minutes after completion of first infusion) and at trough (up to 4 hours before infusion on days 2, 5, and 8, respectively).

### Outcomes

The primary outcome measure was the clinical status at day 15 as measured on the 7-point ordinal scale of the WHO Master Protocol (v3.0, March 3, 2020): 1. Not hospitalized, no limitation on activities; 2. Not hospitalized, limitation on activities; 3. Hospitalized, not requiring supplemental oxygen; 4. Hospitalized, requiring supplemental oxygen; 5. Hospitalized, on non-invasive ventilation or high flow oxygen devices; 6. Hospitalized, on invasive mechanical ventilation or extracorporeal membrane oxygenation (ECMO); 7. Dead. Secondary efficacy outcome measures included: clinical status and change from baseline of the clinical status at days 3, 5, 8, 11 and 29; time to an improvement of one and two categories as measured on the 7-point ordinal scale or hospital discharge until day 29; change from baseline of the National Early Warning Score 2 (NEWS-2) at days 3, 5, 8, 11, 15 and 29; time to NEWS2 ≤2 or hospital discharge until day 29; time to hospital discharge until day 29 and duration of hospitalisation; time to new mechanical ventilation, ECMO or death until day 29; oxygenation- and ventilator-free days until day 29; in-hospital mortality and mortality at days 28 and 90. Exploratory outcome measures included the proportion of subjects with SARS-CoV-2 detectable in NP swabs at six timepoints from baseline to day 29; the decrease of the normalized SARS-CoV-2 viral load in NP swabs from baseline to day 15; the post-infusion plasma concentration of remdesivir and GS-441524 at day 1 and the trough at days 2, 5 and 8.

Safety outcomes included the cumulative incidence of any grade 3 or 4 AE or of any Serious Adverse Event (SAE) and the grade changes in the biological and inflammatory patterns of participants over time, coded using the medical dictionary for regulatory affairs, v23.0 and graded according to the Division of AIDS (DAIDS, Table for Grading the Severity of Adult and Paediatric Adverse Events, v2.1, July 2017).

#### Virological methods

Systematic determination of the normalized viral load blinded to treatment arm was performed on NPS specimens by RNA extraction on the EMAG® platform (bioMerieux, Marcy-l’Étoile, France) following manufacturer’s instructions. The SARS-CoV-2 load was measured by quantitative RT-PCR, according to a scale of calibrated in-house plasmid, using the RT-PCR RdRp-IP4 developed by the Institut Pasteur (Paris, France) (18). The amplification protocol was developed using QuantStudio 5 rtPCR Systems (Thermo Fisher Scientific, Waltham, Massachusetts, USA). The number of cells in sample (quality criteria for NPS and normalization tool or viral load determination) was checked using the CELL Control r-gene® kit (Argene-BioMérieux, Marcy-l’Étoile, France). If cell quantification was below 500 cells/reaction, the quality of the sample was considered too low to be measured. We computed a normalized SARS-CoV-2 load (in log10 of RNA copies per 10 000 cells) by dividing the viral load by the number of cells. All viral loads strictly below 1 log10 RNA copies/10 000 cells were considered under the limit of detection and were reported as a negative result. Any point of kinetics corresponding to a rebound of SARS-CoV-2 detection was tested again for confirmation.

#### Pharmacological methods

Concentrations of remdesivir and its metabolite GS-441524 were determined in plasma using an UPLC-MS/MS (Waters Model Xevo TQ-D) method after precipitation of plasma proteins (19). The active triphosphorylated metabolite GS-443902 has not been determined in the peripheral blood mononuclear cells, as this represented a heavy workload for centres in the context of the pandemic. The lower limit of quantification of both remdesivir and GS-441524 was 1 ng/mL.

### Sample size calculation

The sample size was determined assuming the following scenario under SoC for each item of the ordinal scale at day 15: 1, 42%; 2, 38%; 3, 8%; 4, 7%; 5, 2%; 6, 1%; 7, 2%. At the time of the trial design (March 2020), there was a significant uncertainty with these assumptions. We powered the study for an odds ratio of 1.5 (an odds ratio higher than 1 indicates superiority of the experimental treatment over the control for each ordinal scale category), with 90% power and using an overall one-sided type I error rate of 0.05. This size effect appeared statistically relevant, meaning that 52% of patients would be discharged with no limitation of activity at day 15 in the remdesivir arm, instead of 42% of patients in the control arm. We determined that the inclusion of 450 participants in each treatment arm was required; this number was increased to 475 participants per arm to account for unevaluable participants.

#### Interim analyses

An independent data safety and monitoring board (DSMB) externally reviewed the trial data at regular intervals regarding treatment efficacy, safety and futility.

Following cessation of hydroxychloroquine on June 17^th^, 2020 and of both lopinavir/ritonavir containing arms on June 27^th^, 2020, the trial continued the evaluation of remdesivir. On January 13^th^, 2021, the DisCoVeRy DSMB recommended to suspend participant recruitment based on the evaluation of an interim report of 842 randomised participants, of whom 776 participants had been evaluated at day 15 (389 on remdesivir and 387 on SoC, respectively). Calculating conditional power based on the intended recruitment of 900 participants (i.e. an additional 124 evaluable participants), the DSMB estimated the chances of reaching 5% significance on the originally hypothesised odds ratio of 1.5 to be 0.02% at the end of the trial. They also found no evidence of efficacy on the WHO scale at day 29, nor on mortality at day 29 and noticed the low recruitment rate in the trial over the last six weeks. The decision was endorsed by the DisCoVeRy steering committee on January 19^th^, 2021 with subsequent cessation of participant recruitment on January 21^st^, 2021. Since April, 28^th^ 2021, participants enrolled in the trial are randomized (1:1) to receive either AZ7442, a combination of two long-acting antibodies derived from convalescent patients, or placebo.

#### Statistical analyses

The intention-to-treat population included all randomised participants with a positive SARS-CoV-2 PCR obtained in the last 9 days, for whom a valid consent form was obtained and who did not receive any investigational treatment in the last 29 days.

The modified intention-to-treat population included participants from the intention-to-treat population who received at least one dose of the treatment allocated by randomisation. Efficacy analyses were performed on the intention-to-treat population. Safety analyses were performed on the modified intention-to-treat population. Analyses were stratified by baseline severity but not by region of inclusion due to a low number of inclusions in some regions; all tests were two-sided with a type-I error of 0.05. When one endpoint reached statistical significativity, we performed a non-prespecified subgroup analysis according to baseline severity.

For the 7-point ordinal scale, missing data were imputed using the last observation carried forward method, except in the case of known death or hospital discharge, in which case the ordinal scale was imputed to the value of 7 (death) or 2 (not hospitalised, limitation of activities), respectively. For NEWS, oxygenation and mechanical ventilation outcomes, missing data were treated using the last observation carried forward method, except on the day of death, in which case participants were imputed to the worst NEWS value, or considered to require oxygen or mechanical ventilation. For time-to-event analyses, participants were censored at day 29, at their date of loss of follow-up, or of study withdrawal, whichever occurred first. For outcomes in which death was not included, participants who died before day 29 were censored at day 29. Missing SARS-CoV-2 viral loads were not imputed. For the analysis of viral load by mixed models, undetectable viral load values (i.e. values < 1 log10 copies/10 000 cells) were imputed to half the LoD hence 0.7 log10 copies/10 000 cells. In case of several consecutive undetectable values, only the first one was replaced, and the subsequent ones discarded (until the next detectable value if values were available afterwards).

For the 7-point ordinal scale, data were analysed using a proportional odds model. Time-to-event data were analysed using a Cox proportional hazards model. An analysis of covariance was performed for the comparison of oxygenation- and ventilator-free days between arms; in-hospital mortality, 28-day mortality and the number of participants with detectable SARS-CoV-2 in respiratory tract specimens at each time point were analysed using a Cochran-Mantel-Haenszel test. For safety endpoints, the number of participants with at least one AE, with at least one grade 3 or 4 AE and with at least one SAE were compared between groups using a Cochran-Mantel-Haenszel test. Pre-specified subgroup analyses for the primary outcome were performed using proportional odds models across the following subgroups: age (< 50 years, 50-69 years, ≥ 70 years); sex (female, male); duration of symptoms prior to randomisation (≤ 7 days, 8-14 days, >14 days); disease severity (moderate, severe); country. The evolution of the viral load since randomization was analysed using a mixed-effects linear model with a test of treatment effect on the slope, and a non-prespecified subgroup analysis was performed across duration of symptoms prior to randomisation (≤7 days, 8-14 days,>14 days) and disease severity at randomization.

All analyses were performed using SAS v9.4 (SAS Institute, Cary, NC, USA). This trial is registered with the European Clinical Trials Database, 2020-000936-23, and ClinicalTrials.gov, NCT04315948.

## Results

Between March 22^nd^, 2020 and January 21^st^, 2021, 857 participants were randomized to one of the 2 arms across 48 sites in 5 European countries (France, n=724; Belgium, n=51; Portugal, n=36; Austria, n=31; Luxembourg, n=15); 14 participants were excluded from analysis (no valid written informed consent, n=8; previously treated with a study treatment, n=1; lack of positive PCR, n=5;), and 843 were evaluable for analysis: control arm, n=423, remdesivir arm, n=420 (Figure 1). Among participants from the remdesivir arm, the median duration of treatment was 9 days (IQR, 5; 10).

**Figure 1.**
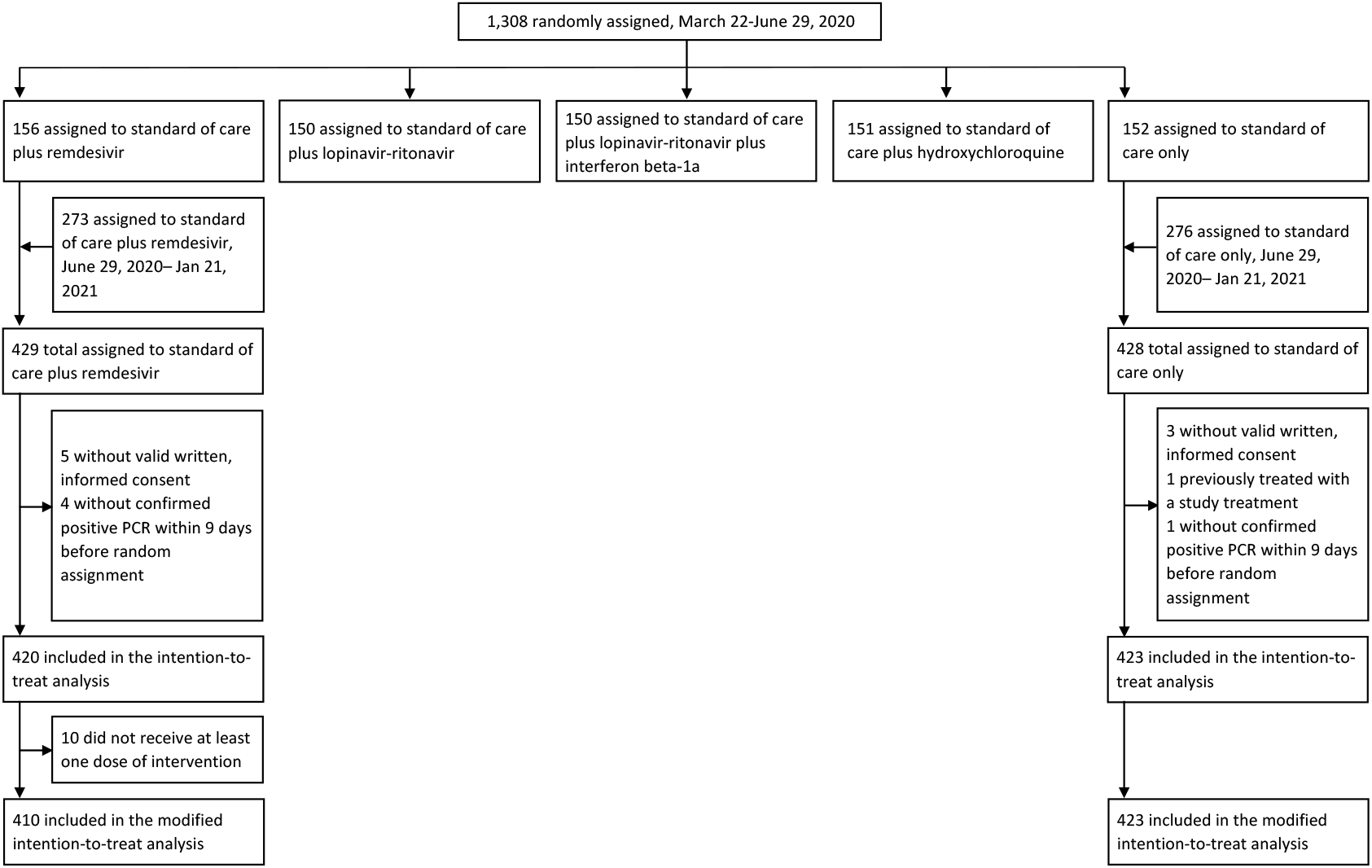
Trial profile. Participants from all groups received the standard of care, in addition to the treatment allocated by randomization. No participant assigned to SoC received remdesivir

Participants’ baseline characteristics are presented in Table 1 and Supplementary Table S1, without notable imbalance between arms. The median age was 64 years (IQR, 54; 73) and 588/843 participants (69.5%) were male, with a median time from symptom onset to randomization of 9 days (IQR, 7; 12). Overall, 618/840 (73.6%) had at least one comorbid condition. The most frequent underlying conditions were obesity (n=284/833, 34.1%), chronic cardiac disease (n=234/839, 28%) and diabetes mellitus (n=225/840, 27%). Upon randomisation, 332/843 participants (39.4%) had a severe COVID-19. Concomitant treatments are presented in Supplementary Table S2. Overall, systemic corticosteroids were administered to 296/843 participants (35.1%); they were more often administrated in participants included after July 1^st^, 2020 (Supplementary Table S3).

**Table 1.**
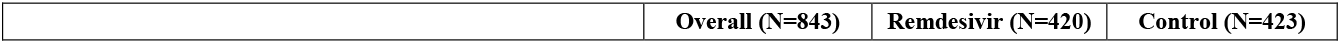
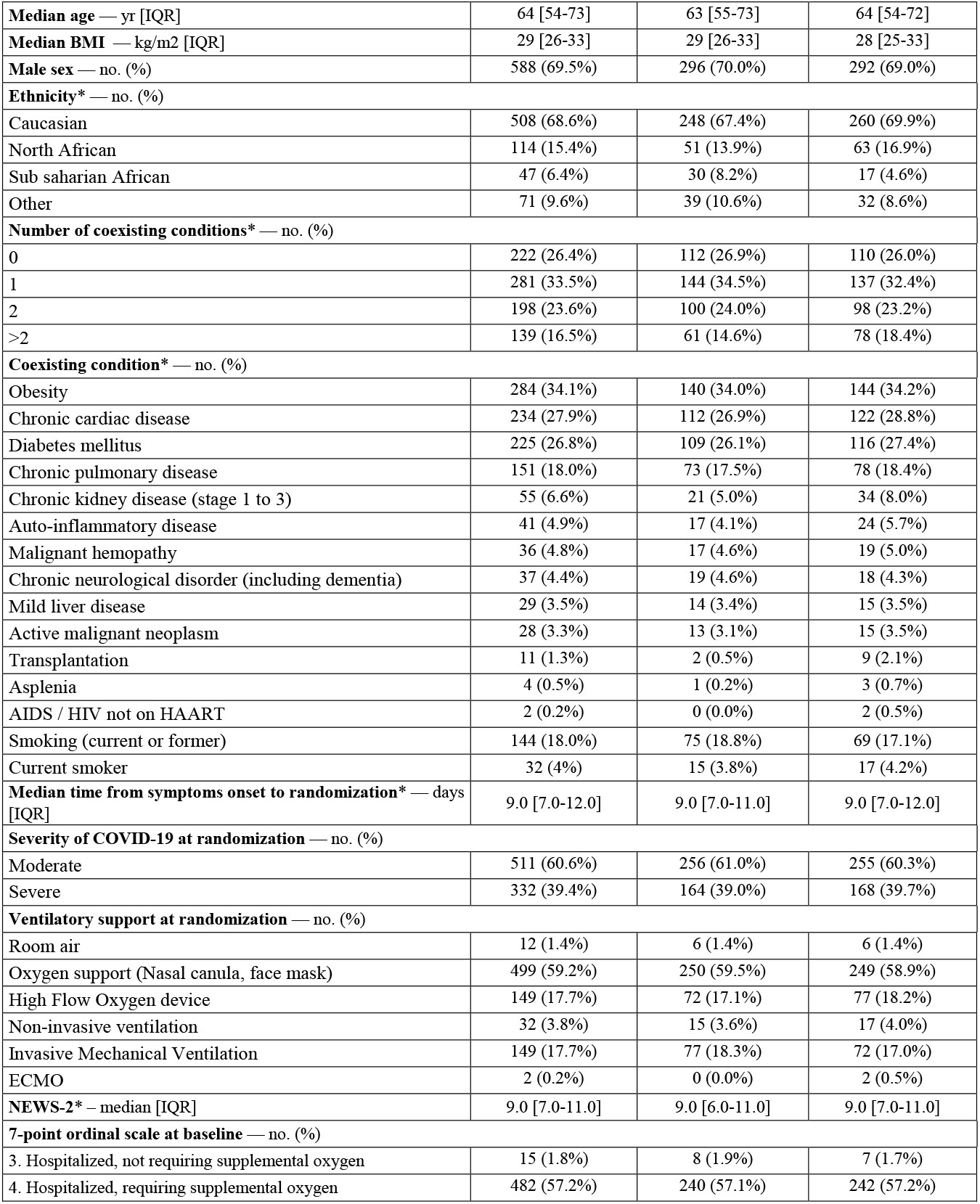
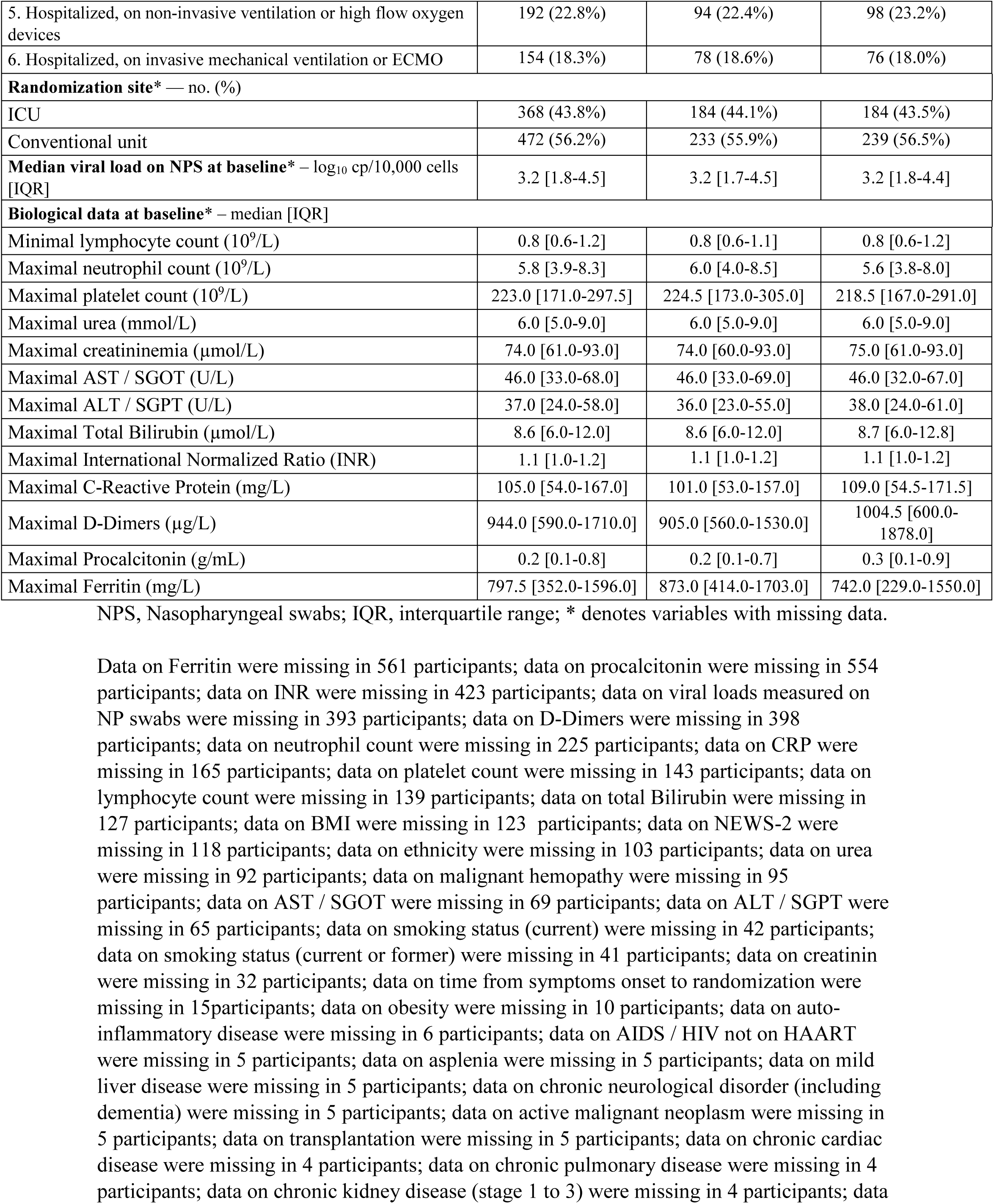
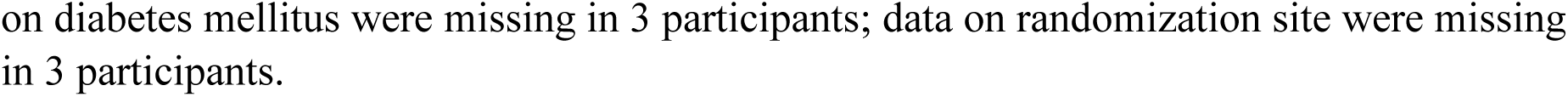
Baseline characteristics of participants included in the intention-to-treat population of the DisCoVeRy trial, overall and according to randomization arm.

|  | Overall (N=843) | Remdesivir (N=420) | Control (N=423) |
| --- | --- | --- | --- |
| **Median age** — yr [IQR] | 64 [54-73] | 63 [55-73] | 64 [54-72] |
| **Median BMI** — kg/m2 [IQR] | 29 [26-33] | 29 [26-33] | 28 [25-33] |
| **Male sex** — no. (%) | 588 (69.5%) | 296 (70.0%) | 292 (69.0%) |
| **Ethnicity\*** — no. (%) |  |  |  |
| Caucasian | 508 (68.6%) | 248 (67.4%) | 260 (69.9%) |
| North African | 114 (15.4%) | 51 (13.9%) | 63 (16.9%) |
| Sub saharian African | 47 (6.4%) | 30 (8.2%) | 17 (4.6%) |
| Other | 71 (9.6%) | 39 (10.6%) | 32 (8.6%) |
| **Number of coexisting conditions\*** — no. (%) |  |  |  |
| 0 | 222 (26.4%) | 112 (26.9%) | 110 (26.0%) |
| 1 | 281 (33.5%) | 144 (34.5%) | 137 (32.4%) |
| 2 | 198 (23.6%) | 100 (24.0%) | 98 (23.2%) |
| >2 | 139 (16.5%) | 61 (14.6%) | 78 (18.4%) |
| **Coexisting condition\*** — no. (%) |  |  |  |
| Obesity | 284 (34.1%) | 140 (34.0%) | 144 (34.2%) |
| Chronic cardiac disease | 234 (27.9%) | 112 (26.9%) | 122 (28.8%) |
| Diabetes mellitus | 225 (26.8%) | 109 (26.1%) | 116 (27.4%) |
| Chronic pulmonary disease | 151 (18.0%) | 73 (17.5%) | 78 (18.4%) |
| Chronic kidney disease (stage 1 to 3) | 55 (6.6%) | 21 (5.0%) | 34 (8.0%) |
| Auto-inflammatory disease | 41 (4.9%) | 17 (4.1%) | 24 (5.7%) |
| Malignant hemopathy | 36 (4.8%) | 17 (4.6%) | 19 (5.0%) |
| Chronic neurological disorder (including dementia) | 37 (4.4%) | 19 (4.6%) | 18 (4.3%) |
| Mild liver disease | 29 (3.5%) | 14 (3.4%) | 15 (3.5%) |
| Active malignant neoplasm | 28 (3.3%) | 13 (3.1%) | 15 (3.5%) |
| Transplantation | 11 (1.3%) | 2 (0.5%) | 9 (2.1%) |
| Asplenia | 4 (0.5%) | 1 (0.2%) | 3 (0.7%) |
| AIDS / HIV not on HAART | 2 (0.2%) | 0 (0.0%) | 2 (0.5%) |
| Smoking (current or former) | 144 (18.0%) | 75 (18.8%) | 69 (17.1%) |
| Current smoker | 32 (4%) | 15 (3.8%) | 17 (4.2%) |
| **Median time from symptoms onset to randomization\*** — days [IQR] | 9.0 [7.0-12.0] | 9.0 [7.0-11.0] | 9.0 [7.0-12.0] |
| **Severity of COVID-19 at randomization** — no. (%) |  |  |  |
| Moderate | 511 (60.6%) | 256 (61.0%) | 255 (60.3%) |
| Severe | 332 (39.4%) | 164 (39.0%) | 168 (39.7%) |
| **Ventilatory support at randomization** — no. (%) |  |  |  |
| Room air | 12 (1.4%) | 6 (1.4%) | 6 (1.4%) |
| Oxygen support (Nasal canula, face mask) | 499 (59.2%) | 250 (59.5%) | 249 (58.9%) |
| High Flow Oxygen device | 149 (17.7%) | 72 (17.1%) | 77 (18.2%) |
| Non-invasive ventilation | 32 (3.8%) | 15 (3.6%) | 17 (4.0%) |
| Invasive Mechanical Ventilation | 149 (17.7%) | 77 (18.3%) | 72 (17.0%) |
| ECMO | 2 (0.2%) | 0 (0.0%) | 2 (0.5%) |
| **NEWS-2\*** – median [IQR] | 9.0 [7.0-11.0] | 9.0 [6.0-11.0] | 9.0 [7.0-11.0] |
| **7-point ordinal scale at baseline** — no. (%) |  |  |  |
| 3. Hospitalized, not requiring supplemental oxygen | 15 (1.8%) | 8 (1.9%) | 7 (1.7%) |
| 4. Hospitalized, requiring supplemental oxygen | 482 (57.2%) | 240 (57.1%) | 242 (57.2%) |
| 5. Hospitalized, on non-invasive ventilation or high flow oxygen devices | 192 (22.8%) | 94 (22.4%) | 98 (23.2%) |
| 6. Hospitalized, on invasive mechanical ventilation or ECMO | 154 (18.3%) | 78 (18.6%) | 76 (18.0%) |
| **Randomization site\* — no. (%)** |  |  |  |
| ICU | 368 (43.8%) | 184 (44.1%) | 184 (43.5%) |
| Conventional unit | 472 (56.2%) | 233 (55.9%) | 239 (56.5%) |
| **Median viral load on NPS at baseline\* — log<sub>10</sub> cp/10,000 cells [IQR]** | 3.2 [1.8-4.5] | 3.2 [1.7-4.5] | 3.2 [1.8-4.4] |
| **Biological data at baseline\* — median [IQR]** |  |  |  |
| Minimal lymphocyte count (10<sup>9</sup>/L) | 0.8 [0.6-1.2] | 0.8 [0.6-1.1] | 0.8 [0.6-1.2] |
| Maximal neutrophil count (10<sup>9</sup>/L) | 5.8 [3.9-8.3] | 6.0 [4.0-8.5] | 5.6 [3.8-8.0] |
| Maximal platelet count (10<sup>9</sup>/L) | 223.0 [171.0-297.5] | 224.5 [173.0-305.0] | 218.5 [167.0-291.0] |
| Maximal urea (mmol/L) | 6.0 [5.0-9.0] | 6.0 [5.0-9.0] | 6.0 [5.0-9.0] |
| Maximal creatininemia (μmol/L) | 74.0 [61.0-93.0] | 74.0 [60.0-93.0] | 75.0 [61.0-93.0] |
| Maximal AST / SGOT (U/L) | 46.0 [33.0-68.0] | 46.0 [33.0-69.0] | 46.0 [32.0-67.0] |
| Maximal ALT / SGPT (U/L) | 37.0 [24.0-58.0] | 36.0 [23.0-55.0] | 38.0 [24.0-61.0] |
| Maximal Total Bilirubin (μmol/L) | 8.6 [6.0-12.0] | 8.6 [6.0-12.0] | 8.7 [6.0-12.8] |
| Maximal International Normalized Ratio (INR) | 1.1 [1.0-1.2] | 1.1 [1.0-1.2] | 1.1 [1.0-1.2] |
| Maximal C-Reactive Protein (mg/L) | 105.0 [54.0-167.0] | 101.0 [53.0-157.0] | 109.0 [54.5-171.5] |
| Maximal D-Dimers (μg/L) | 944.0 [590.0-1710.0] | 905.0 [560.0-1530.0] | 1004.5 [600.0-1878.0] |
| Maximal Procalcitonin (g/mL) | 0.2 [0.1-0.8] | 0.2 [0.1-0.7] | 0.3 [0.1-0.9] |
| Maximal Ferritin (mg/L) | 797.5 [352.0-1596.0] | 873.0 [414.0-1703.0] | 742.0 [229.0-1550.0] |

NPS, Nasopharyngeal swabs; IQR, interquartile range; \* denotes variables with missing data.

At day 15, the distribution of the WHO ordinal scale was as follow in the remdesivir and control groups, respectively: Not hospitalized, no limitations on activities: 62/420 (14.8%) and 72/423 (17.0%); Not hospitalized, limitation on activities: 126/420 (30%) and 135/423 (31.9%); Hospitalized, not requiring supplemental oxygen: 56/420 (13.3%) and 31/423 (7.3%); Hospitalized, requiring supplemental oxygen: 75/420 (18%) and 65/423 (15.4%); Hospitalized, on non-invasive ventilation or high flow oxygen devices: 16/420 (3.8%) and 16/423 (3.8%); Hospitalized, on invasive mechanical ventilation or ECMO: 64/420 (15.2%) and 80/423 (19%); Death: 21/420 (5%) and 24/423 (6%). Overall, the ordinal scale was missing in less than 5% of participants at day 15 and in 7% of participants at day 29, without imbalance between groups; There was no significant difference between remdesivir and control (odds-ratio [OR], 1.02, 95% confidence interval [CI], 0.62 to 1.70, P=0.93) (Figure 2, Table 2 and Supplementary Figure S1). No significant difference was observed between remdesivir and control in subgroup analyses according to age, sex, duration of symptoms before randomisation, disease severity or country of randomisation (Supplementary Figure S2).

**Table 2.**
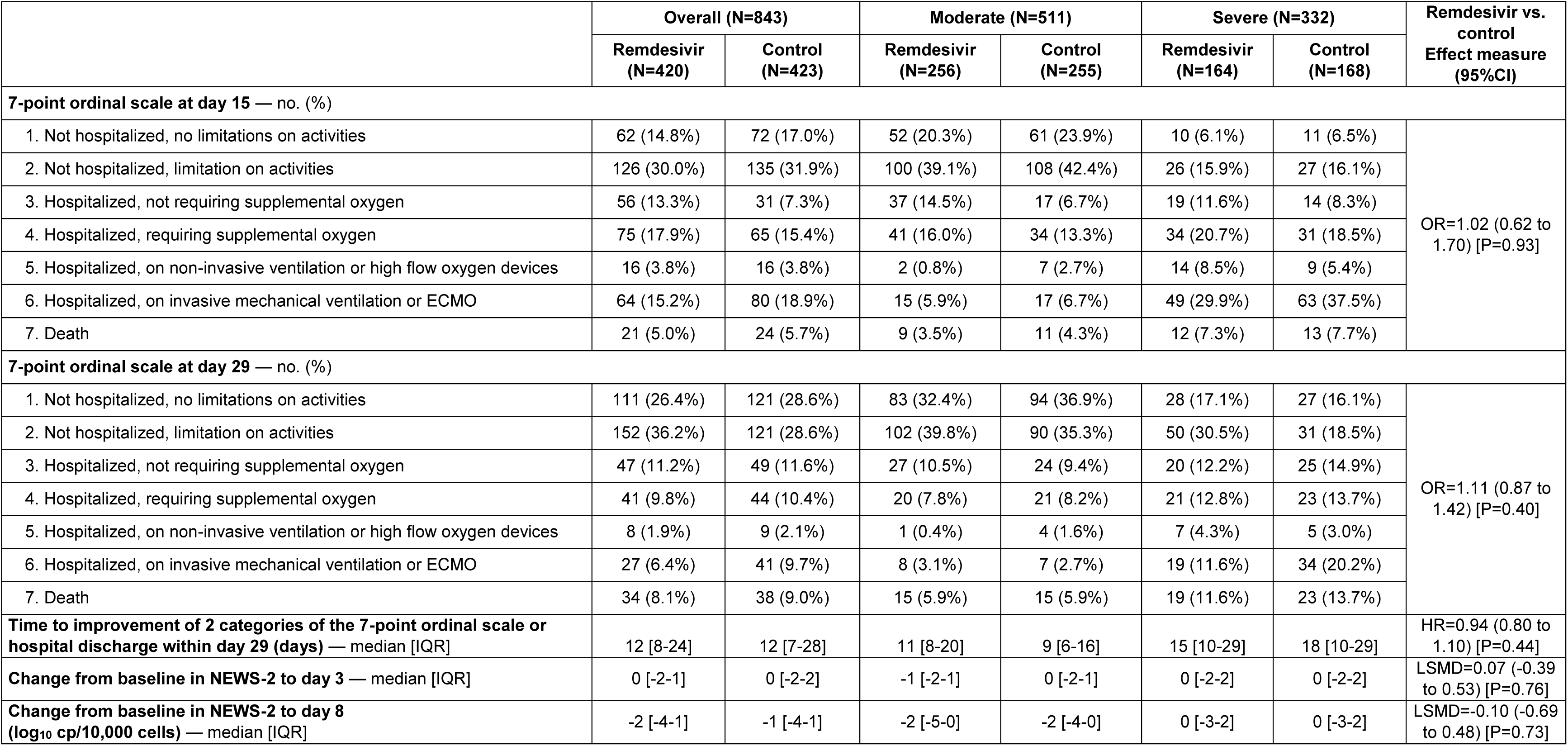
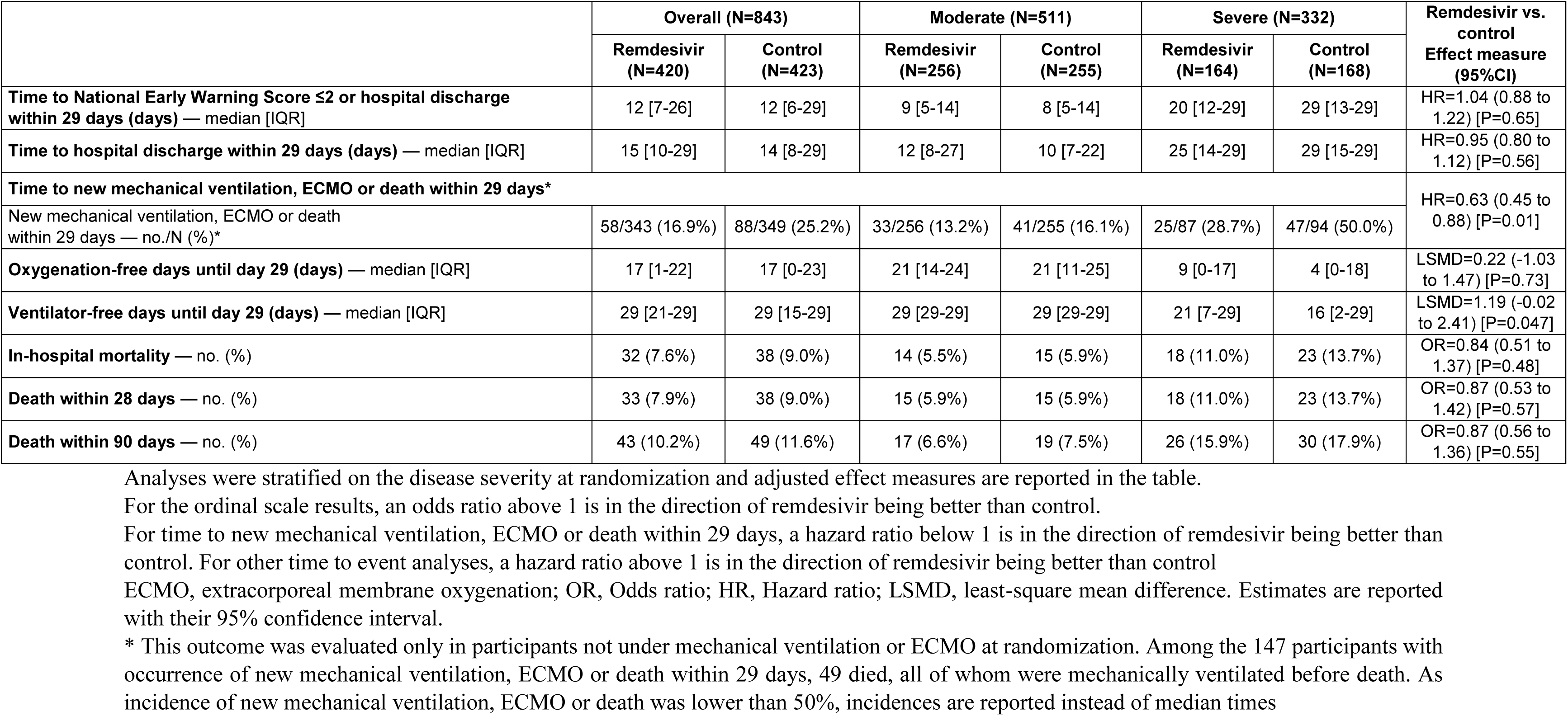
Primary and secondary outcomes in the intention-to-treat population of the DisCoVeRy trial, overall, according to randomization arm and to severity at randomization.

**Figure 2.**
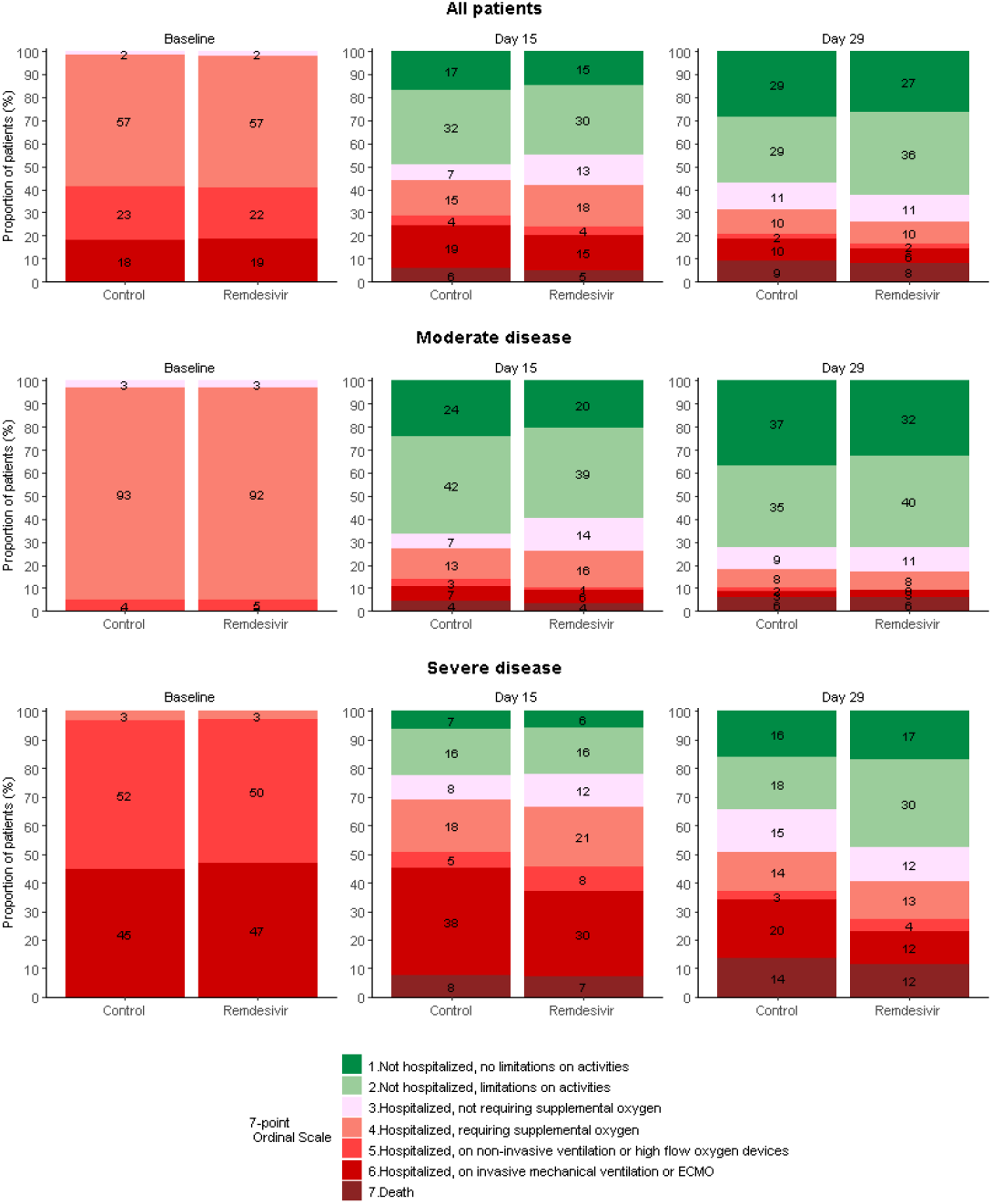
Clinical status at baseline, day 15 and day 29 in the intention-to-treat population of the DisCoVeRy trial, overall and according to randomization arm and disease severity at randomization. Overall, 24 participants were randomized as moderate participants but had their disease severity revised to severe at the baseline evaluation, and 21 participants were randomized as severe participants but had their disease severity revised to moderate at the baseline evaluation. Reported numbers refer to the proportion of participants with the corresponding level in each group.

There was no significant difference between remdesivir and control in the distribution of the 7-point ordinal scale at day 29 (OR, 1.11, 95% CI, 0.87 to 1.43, P=0.40) (Figure 2, Table 2 and Supplementary Figure S1).

In-hospital, day 28 and day 90 mortality rates were not different between remdesivir and control treatment groups (Table 2). In participants without mechanical ventilation nor ECMO at randomisation (n=692), the time to the composite endpoint of new mechanical ventilation, ECMO, or death was significantly longer in the remdesivir arm than in the control arm (Hazard ratio [HR], 0.63, 95% CI, 0.45 to 0.88, P=0.01, Table 2 and Supplementary Figure S3). In non-prespecified analyses, this effect was significant in participants with severe disease at randomisation (HR=0.49, 95%CI 0.30 to 0.80, P=0.004), but not in participants with moderate disease (HR=0.79, 95%CI, 0.50 to 1.25, P=0.31).

No other significant difference was observed for any other secondary outcomes between remdesivir and control (Table 2, Supplementary Table S4 and Supplementary Figures S4-S9). A total of 2,981 NP swabs were analysed in 694 participants. The median normalised viral loads were 3.2 log10 cp/10,000 cells (IQR, 1.7; 4.5) and 3.2 log10 cp/10,000 cells (IQR, 1.8; 4.4) in the remdesivir and control arms, respectively (Table 1). The median decrease in viral loads between baseline and day 3 was 0.5 log10 cp/10,000 cells (IQR −1.4; 0.0) and 0.5 log10 cp/10,000 cells (IQR −1.3; 0.1) in the remdesivir and control arms, respectively (Supplementary Table S5). There was no significant effect of remdesivir on the viral kinetics (effect of remdesivir on slope, −0.006 log10 cp/10,000 cells/day, 95% CI, −0.02 to 0.03, P=0.66; Figure 3, Supplementary Table S6 and Supplementary Figure S10). Similar results were obtained in subgroup analyses according to severity at randomisation or duration of symptoms (Supplementary Table S7). Accordingly, there was no significant difference in the proportion of participants with detectable viral loads at each sampling time (Supplementary Table S5).

**Figure 3.**
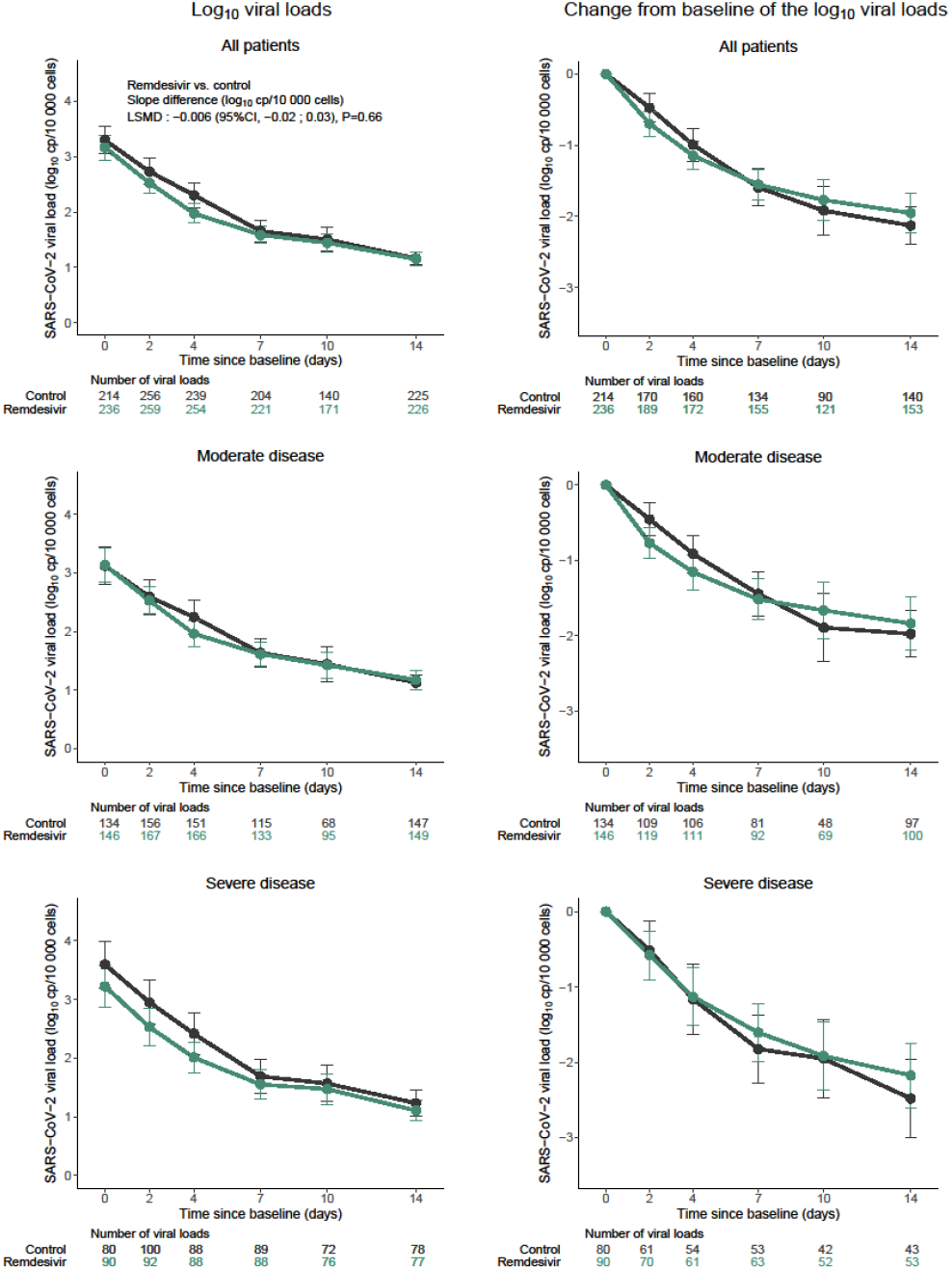
Evolution of the normalized SARS-CoV-2 viral load in nasopharyngeal swabs between baseline and day 15 in the intention-to-treat population of the DisCoVeRy trial. Data are presented as means (95%CI). Remdesivir (green line); control (black line). LSMD, least-square mean difference; 95%CI, 95% confidence interval.

Median post-infusion concentrations of remdesivir and GS-441524 at day 1 for a subset of 58 participants were 2,541 ng/mL (IQR, 1,417; 4,845) and 74 ng/mL (IQR, 51; 105), respectively. Trough plasma concentrations of remdesivir were below the limit of quantification for all participants, while median trough plasma concentrations of GS-441524 at days 2, 5 and 8 were stable at 66 ng/mL (n=49; IQR, 50; 102), 59 ng/mL (n=32; IQR, 46; 86) and 63 ng/mL (n=22; IQR 44; 87), respectively. Results according to disease severity at randomisation are presented in Supplementary Table S8.

A total of 833 participants were included in the safety analysis (remdesivir, n=410; control, n=423). Safety outcomes are presented in Table 3 and Supplementary Table S9. Among the 1896 reported AEs, 625 were graded 3 or 4 adverse events affecting 34.9% of participants (143/410) from the remdesivir arm and 35.5% (150/423) from the control arm (P=0.91). Five hundred forty-two serious adverse events (SAEs) were reported affecting 35.9% participants (147/410) from the remdesivir arm and 32.6% (138/423) from the control arm (P=0.29). Three deaths (ARDS, bacterial infection, and hepatorenal syndrome) were considered related to remdesivir by the investigators, but only one by the sponsor’s safety team (hepatorenal syndrome). The most frequently reported SAEs were ARDS (81/833, 10%), acute respiratory failure (77/833, 9.2%), - and acute kidney injury (33/833, 4%) (Table 3).

**Table 3.** Summary of adverse events in the modified intention to treat population of the DisCoVeRy trial, overall and according to randomization arm. Some patients had more than a single SAE. Analyses were performed on the modified Intention-to-treat population. OR, Odds ratio; SAE, Serious Adverse Event. * Excluding acute renal failures defined based on the RIFLE classification.

|  | **Remdesivir**<br>**(n=410)** | **Control**<br>**(n=423)** | **Remdesivir vs. Control**<br>**Effect measure (95%CI)** |
| --- | --- | --- | --- |
| **Any adverse events – no. (%)** | 256 (62.4%) | 248 (58.6%) | OR=1.19 (0.89 to 1.57)<br>[P=0.24] |
| **Any grade 3 and 4 adverse events – no. (%)** | 143 (34.9%) | 150 (35.5%) | OR=0.98 (0.73 to 1.32)<br>[P=0.91] |
| **Any serious adverse events – no. (%)** | 147 (35.9%) | 138 (32.6%) | OR=1.17 (0.87 to 1.57)<br>[P=0.29] |
| **Most relevant SAE – no. (%)** |  |  |  |
| Acute kidney injury* | 15 (4%) | 18 (4%) |  |
| Acute renal failure based on the RIFLE classification | 1 (0%) | 4 (1%) |  |
| Acute respiratory distress syndrom | 40 (10%) | 41 (10%) |  |
| Acute respiratory failure | 26 (6%) | 51 (12%) |  |
| Sepsis | 9 (2%) | 10 (2%) |  |
| Arrhythmia | 15 (4%) | 8 (2%) |  |
| Transaminases increased | 11 (3%) | 3 (1%) |  |
| Pulmonary embolism | 10 (2%) | 12 (3%) |  |
| Cholestasis | 0 (0%) | 0 (0%) |  |

## Discussion

Here, we report the final results of the DisCoVeRy trial comparing remdesivir to control in hospitalised patients with COVID-19, which confirm what was observed in preliminary analyses. Remdesivir administration was well tolerated but was neither associated with a better clinical outcome at day 15 and 29 nor with a faster viral clearance.

Regarding day-15 clinical status, the discrepancy observed between the present results and those from the ACTT-1 (10) (which contributed to obtain EUA) may be explained by the differences in study populations: in ACTT-1, a smaller percentage of participants required oxygen support at baseline (87% in ACTT-1 *vs*. 98% in DisCoVeRy, which might be due to differences in participants’ inclusion criteria and severity at inclusion) and fewer received corticosteroids (23% received corticosteroids in ACTT-1 *vs*. 35% in DisCoVeRy). In DisCoVery, among the subset of participants without mechanical ventilation or ECMO at randomisation, remdesivir significantly delayed the need of new mechanical ventilation or ECMO, or death, consistent with what was reported in ACTT-1 (10). This suggests that remdesivir could delay the worsening of respiratory disorders. Nevertheless, the decision to implement mechanical ventilation or ECMO is prone to vary based on investigator’s judgement and centre practices. In addition, this effect was not observed for other secondary outcomes of respiratory status, such as the NEWS-2 score, and it did not translate into a reduced mortality rate at day 28, similar to what Solidarity trial reported on in-hospital mortality (11). In the meta-analysis of four trials which compared remdesivir to control, the conclusion was that remdesivir might have little or no effect on mortality (11).

In DisCoVeRy, SARS-CoV-2 kinetic assessments were centralised and normalised to ensure consistency throughout centres. There was no effect of remdesivir on SARS-CoV-2 viral kinetics, consistent with previous results (9,20). This could be due to a genuine lack of effect but could also reflect that treatment was administered too late to be effective (median of nine days after onset of symptoms). Modelling studies of SARS-CoV-2 infection (21,22) have suggested that antiviral efficacy depends on early administration, before attaining the peak viral load (23). Consistently, clinical studies on influenza have demonstrated that the administration of oseltamivir within 48 hours after the onset of symptoms is required to ensure decreased viral shedding (24). Recent results obtained with an alternative antiviral approach through infusion of anti-SARS-CoV-2 antibodies confirm this need of early treatment to ensure effectiveness (25–28). Of note, however, when restricting the viral kinetics analysis to participants who initiated treatment within seven days after onset of symptoms, still no effect of remdesivir on SARS-CoV-2 clearance was observed.

Post-infusion plasma concentrations of remdesivir were consistent with those reported in healthy volunteers (29), and trough plasma concentrations after day 1 were undetectable in all participants, consistent with the estimated 1-hour elimination half-life (29). This is probably due to rapid entry of remdesivir into cells. GS-441524 is one intracellular remdesivir metabolite able to cross cellular membranes and whose levels can be measured in plasma. It is renally eliminated unchanged (27-hour half-life). Over the study period, trough concentrations of GS-441524 were consistent with those previously reported (30). Although we were not able to measure the active tri phosphorylated compound, the lack of viral efficacy is not likely attributable to inappropriate drug levels.

The trial has some limitations. It was open, and not placebo-controlled. Indeed, several treatments were concomitantly evaluated at the beginning of the trial, and blinding was thus impossible due to the different modes of administration (intravenous, subcutaneous or oral) of the different treatment arms. This might have introduced bias in the follow up and management of patients, and in the evaluation of endpoints whose assessment contains elements of subjectivity: decision to begin corticosteroids in patients management or to begin mechanical ventilation might have been influenced by the knowledge of the treatment arm, even unconsciously. This risk of bias is however mitigated for the viral load, which was analysed blindly from treatment arm. Next, viral load assessment was not available for 18% of participants (nearly half at baseline). However, the proportions of participants with available viral loads at each sampling time were similar in both experimental groups, suggesting that NP sampling was not guided by the allocated treatment. Finally, plasma concentrations of the prodrug remdesivir and GS-441524 were assessed in only 10% of participants and the concentrations of its intracellular active metabolite were not measured. Although the trial was not designed as a pharmacokinetic study, it provides currently lacking data on remdesivir exposure in patients hospitalised with COVID-19.

## Conclusion

In this randomised controlled trial, the use of remdesivir for the treatment of hospitalised patients with COVID-19 was not associated with clinical improvement at day 15 or day 29, nor with a reduction in mortality, nor with a reduction in SARS-CoV-2 RNA.

## Data Availability

All data produced in the present study are available upon reasonable request to the authors

## Data Availability

All data produced in the present study are available upon reasonable request to the authors

## Contributors

FA, NPS, JPo, MBD, GP, BLi, DC, YY, CB, FM were involved in the design, establishment, and day-to-day management and implementation of the trial. FA, MH, CD, JS, DC, YY and FM obtained funding for the trial. FA, MBD, MH, NPS, JPo, MPL, GP, ST, XL, EF, SN, JCR, FW, FG, BLe, AK, FR, BG, JR, JPL, CA, YZ, FBS, BM, FD, YR, RCJ, VLM, KK, KL, GMB, FVB, AC, JMT, LP, MB, EBN, AGB, GT, FB, RGa, JM, SG, DG, KB, LE, SJ, AG, GV, SB, JMFR, MJ, TS, AA, RGr, AE, MN, RRA, JAP, BLi included participants in the trial. MBD and AG were responsible for the virological analyses. MPL and GP were responsible for the pharmacological analyses. ADi, ADe, CF, NM, PV, TA were in charge of data curation. They had full access to the data. DB, JG, DC, CB and FM were involved in the statistical analyses. FA, ADi, MH, MBD, CB, FM wrote the original draft of the manuscript, which was reviewed and edited by NPS, JPo, DB, MPL, GP, DC, YY. All authors contributed to refinement of and approved this manuscript. The corresponding author had full access to all the data in the study and had final responsibility for the decision to submit for publication.

## Data sharing

With publication, deidentified, individual participant data that underlie this Article, along with a data dictionary describing variables in the dataset, will be made available to researchers whose proposed purpose of use is approved by the DisCoVeRy Steering Committee. To request the dataset, please address directly to the Corresponding author or to the Sponsor’s representative to obtain a data access form. All requests will be evaluated by the Trial Management Team and the DisCoVeRy Steering Committee. For accepted requests, data will be shared after signing a Data Transfer Agreement with the Study Sponsor. Data will be shared directly or through access on the Inserm repository. Related documents, such as the study protocol, statistical analysis plan, and informed consent form, will be made available (with publication) on request to the Corresponding author or to the Sponsor’s representative. The data will be open access for the informed consent form, protocol, and statistical analysis plan.

## Acknowledgements

This work received funding from several sources:

- *Europe*: European Union’s Horizon 2020 research and innovation programme
- *Austria*: AGMT gGmbH
- *Belgium*: Belgian Health Care Knowledge Centre; Fonds Erasme-COVID-ULB
- *France*: REACTing, a French multi-disciplinary collaborative network working on emerging infectious diseases; Ministry of Health; Paris Ile-de-France Region
- *Luxemburg*: European Regional Development Fund
- *Portugal*: Ministry of Health; Agency for Clinical Research and Biomedical Innovation

Remdesivir was provided by Gilead free of charge.

We thank all participants who accepted to enrol in the trial, as well as all study and site staff whose indispensable assistance made possible the conduct of the DisCoVeRy trial. They are all listed in the Supplementary Appendix.

## References

1. Ader F. Protocol for the DisCoVeRy trial: multicentre, adaptive, randomised trial of the safety and efficacy of treatments for COVID-19 in hospitalised adults. BMJ Open. 21 sept 2020;10(9):e041437.

2. Ader F, Peiffer-Smadja N, Poissy J, Bouscambert-Duchamp M, Belhadi D, Diallo A, et al. An open-label randomized controlled trial of the effect of lopinavir/ritonavir, lopinavir/ritonavir plus IFN-β-1a and hydroxychloroquine in hospitalized patients with COVID-19. Clinical Microbiology and Infection. éc 2021;27(12):1826–37.

3. Ader F, Peiffer-Smadja N, Poissy J, Bouscambert-Duchamp M, Belhadi D, Diallo A, et al. An open-label randomized, controlled trial of the effect of lopinavir/ritonavir, lopinavir/ritonavir plus IFN-beta-1a and hydroxychloroquine in hospitalized patients with COVID-19 - Final results from the DisCoVeRy trial [Internet]. Infectious Diseases (except HIV/AIDS); 2022 févr [cité 22 févr 2022]. Disponible sur: http://medrxiv.org/lookup/doi/10.1101/2022.02.16.22271064

4. Lê Mp, Le Hingrat Q, Jaquet P, Wicky P-H, Bunel V, Massias L, et al. Removal of Remdesivir’s Metabolite GS-441524 by Hemodialysis in a Double Lung Transplant Recipient with COVID-19. Antimicrob Agents Chemother. 20 oct 2020;64(11):e01521–20.

5. European Medicine Agency. Summary on compassionate use [Internet]. 2020. Disponible sur: https://www.ema.europa.eu/en/documents/other/summary-compassionate-use-remdesivir-gilead_en.pdf

6. Warren TK, Jordan R, Lo MK, Ray AS, Mackman RL, Soloveva V, et al. Therapeutic efficacy of the small molecule GS-5734 against Ebola virus in rhesus monkeys. Nature. 17 mars 2016;531(7594):381–5.

7. Wang M, Cao R, Zhang L, Yang X, Liu J, Xu M, et al. Remdesivir and chloroquine effectively inhibit the recently emerged novel coronavirus (2019-nCoV) in vitro. Cell Res. mars 2020;30(3):269–71.

8. Williamson BN, Feldmann F, Schwarz B, Meade-White K, Porter DP, Schulz J, et al. Clinical benefit of remdesivir in rhesus macaques infected with SARS-CoV-2. Nature. sept 2020;585(7824):273–6.

9. Wang Y, Zhang D, Du G, Du R, Zhao J, Jin Y, et al. Remdesivir in adults with severe COVID-19: a randomised, double-blind, placebo-controlled, multicentre trial. Lancet. 16 mai 2020;395(10236):1569–78.

10. Beigel JH, Tomashek KM, Dodd LE, Mehta AK, Zingman BS, Kalil AC, et al. Remdesivir for the Treatment of Covid-19 - Final Report. N Engl J Med. 5 nov 2020;383(19):1813–26.

11. Pan H, Peto R, Henao-Restrepo A-M, Preziosi M-P, Sathiyamoorthy V, Abdool Karim Q, et al. Repurposed Antiviral Drugs for Covid-19 - Interim WHO Solidarity Trial Results. N Engl J Med. 11 févr 2021;384(6):497–511.

12. Ader F, Bouscambert-Duchamp M, Hites M, Peiffer-Smadja N, Poissy J, Belhadi D, et al. Remdesivir plus standard of care versus standard of care alone for the treatment of patients admitted to hospital with COVID-19 (DisCoVeRy): a phase 3, randomised, controlled, open-label trial. The Lancet Infectious Diseases. févr 2022;22(2):209–21.

13. RECOVERY Collaborative Group, Horby P, Lim WS, Emberson JR, Mafham M, Bell JL, et al. Dexamethasone in Hospitalized Patients with Covid-19. N Engl J Med. 25 févr 2021;384(8):693–704.

14. The WHO Rapid Evidence Appraisal for COVID-19 Therapies (REACT) Working Group, Sterne JAC, Murthy S, Diaz JV, Slutsky AS, Villar J, et al. Association Between Administration of Systemic Corticosteroids and Mortality Among Critically Ill Patients With COVID-19: A Meta-analysis. JAMA. 6 oct 2020;324(13):1330.

15. Villar J, Ferrando C, Martínez D, Ambrós A, Muñoz T, Soler JA, et al. Dexamethasone treatment for the acute respiratory distress syndrome: a multicentre, randomised controlled trial. Lancet Respir Med. mars 2020;8(3):267–76.

16. Tang N, Bai H, Chen X, Gong J, Li D, Sun Z. Anticoagulant treatment is associated with decreased mortality in severe coronavirus disease 2019 patients with coagulopathy. J Thromb Haemost. mai 2020;18(5):1094–9.

17. Paranjpe I, Fuster V, Lala A, Russak AJ, Glicksberg BS, Levin MA, et al. Association of Treatment Dose Anticoagulation With In-Hospital Survival Among Hospitalized Patients With COVID-19. J Am Coll Cardiol. 7 juill 2020;76(1):122–4.

18. Etievant S, Bal A, Escuret V, Brengel-Pesce K, Bouscambert M, Cheynet V, et al. Performance Assessment of SARS-CoV-2 PCR Assays Developed by WHO Referral Laboratories. J Clin Med. 16 juin 2020;9(6):E1871.

19. Avataneo V, de Nicolò A, Cusato J, Antonucci M, Manca A, Palermiti A, et al. Development and validation of a UHPLC-MS/MS method for quantification of the prodrug remdesivir and its metabolite GS-441524: a tool for clinical pharmacokinetics of SARS-CoV-2/COVID-19 and Ebola virus disease. Journal of Antimicrobial Chemotherapy. 1 juill 2020;75(7):1772–7.

20. Barratt-Due A, Olsen IC, Nezvalova-Henriksen K, Kåsine T, Lund-Johansen F, Hoel H, et al. Evaluation of the Effects of Remdesivir and Hydroxychloroquine on Viral Clearance in COVID-19 : A Randomized Trial. Ann Intern Med. 13 juill 2021;174(9):1261–9.

21. Gonçalves A, Bertrand J, Ke R, Comets E, de Lamballerie X, Malvy D, et al. Timing of Antiviral Treatment Initiation is Critical to Reduce SARS-CoV-2 Viral Load. CPT Pharmacometrics Syst Pharmacol. sept 2020;9(9):509–14.

22. Néant N, Lingas G, Le Hingrat Q, Ghosn J, Engelmann I, Lepiller Q, et al. Modeling SARS-CoV-2 viral kinetics and association with mortality in hospitalized patients from the French COVID cohort. Proc Natl Acad Sci U S A. 23 févr 2021;118(8):e2017962118.

23. Rasmussen AL, Popescu SV. SARS-CoV-2 transmission without symptoms. Science. 19 mars 2021;371(6535):1206–7.

24. Nicholson KG, Aoki FY, Osterhaus AD, Trottier S, Carewicz O, Mercier CH, et al. Efficacy and safety of oseltamivir in treatment of acute influenza: a randomised controlled trial. Neuraminidase Inhibitor Flu Treatment Investigator Group. Lancet. 27 mai 2000;355(9218):1845–50.

25. Gottlieb RL, Nirula A, Chen P, Boscia J, Heller B, Morris J, et al. Effect of Bamlanivimab as Monotherapy or in Combination With Etesevimab on Viral Load in Patients With Mild to Moderate COVID-19: A Randomized Clinical Trial. JAMA. 16 févr 2021;325(7):632.

26. Libster R, Pérez Marc G, Wappner D, Coviello S, Bianchi A, Braem V, et al. Early High-Titer Plasma Therapy to Prevent Severe Covid-19 in Older Adults. N Engl J Med. 18 févr 2021;384(7):610–8.

27. Dougan M, Nirula A, Azizad M, Mocherla B, Gottlieb RL, Chen P, et al. Bamlanivimab plus Etesevimab in Mild or Moderate Covid-19. N Engl J Med. 14 juill 2021;385(15):1382–92.

28. Weinreich DM, Sivapalasingam S, Norton T, Ali S, Gao H, Bhore R, et al. REGN-COV2, a Neutralizing Antibody Cocktail, in Outpatients with Covid-19. N Engl J Med. 21 janv 2021;384(3):238–51.

29. Humeniuk R, Mathias A, Cao H, Osinusi A, Shen G, Chng E, et al. Safety, Tolerability, and Pharmacokinetics of Remdesivir, An Antiviral for Treatment of COVID-19, in Healthy Subjects. Clin Transl Sci. sept 2020;13(5):896–906.

30. Humeniuk R, Mathias A, Kirby BJ, Lutz JD, Cao H, Osinusi A, et al. Pharmacokinetic, Pharmacodynamic, and Drug-Interaction Profile of Remdesivir, a SARS-CoV-2 Replication Inhibitor. Clin Pharmacokinet. mai 2021;60(5):569–83.

