## Supplemental material for "Remdesivir for the treatment of hospitalised patients with COVID-19: final results from the DisCoVeRy randomised, controlled, open-label trial"

by Ader, F. et al.

#### Supplementary Methods – Virological Methods

NP swabs were collected through validated devices containing flocked swabs and virus transport medium. Respiratory samples were collected in sterile containers and were sent, as swabs, to the laboratory to be processed within 24 hours after sampling for SARS-CoV-2 detection, blind from allocated treatment. Each sample was then sent after freezing, to the National Centre for Viral Respiratory Infections (Hospices Civils de Lyon, France) for the determination of the normalised viral load. The analysis of the NP swab was performed blinded to treatment arm. RNA extraction was performed on the EMAG® platform (bioMerieux, Marcy-l'Étoile, France), using manufacturer's instructions. RNA was extracted from 200 µL of sample eluted in 50 µL of elution buffer. Then, SARS-CoV-2 viral load was measured by quantitative RT-PCR using the RT-PCR RdRp-IP4 developed by the French National Centre for Viral Respiratory Infections (Institut Pasteur, Paris) widely used in French laboratories at the beginning of the pandemic. The amplification protocol was developed for viral quantification using QuantStudio 5 *rt*PCR Systems (Thermo Fisher Scientific, Waltham, Massachusetts, USA). This RT-PCR targets the RdRp gene and is one of the most sensitive methods available [1] . SARS-COV-2 viral loads were measured according to a scale of calibrated in-house plasmids, harbouring the RdRp target and developed specifically for this quantification. The 10-fold diluted calibrated plasmid ranges from $5.10^2$ to $5.10^6$ copies/5 µL. The strict validation criteria applied to each PCR allowed for a reproductible quantification run after run. The quality of NP swabs was checked using the CELL Control r-gene® kit (Argene_BioMérieux, Marcy-l'Étoile, France) targeting the human gene HPRT-1. This kit is provided with a quantified plasmid for cellular quantification. If cell quantification was below 500 cells/5µL, the sample was considered of poor quality for viral load to be measured. To homogenise the viral loads determinations between participants and sampling times, we computed a normalised SARS-CoV-2 viral load by dividing the viral load measured in the sample by the number of cells measured, and expressed it in $\log_{10}$ of RNA copies per 10 000 cells. The limit of detection (LoD) of RT-PCR for the viral load was 4 copies / reaction, which corresponds to a limit of detection for normalised viral loads of 1 $\log_{10}$ copies/100 000 cells. We estimated the LoD using a probit analysis by analysing dilutions of a quantified cell culture supernatants. Probit analysis consists of describing the relationship between the probability of detection and concentration using a cumulative probability curve. For each dilution, the ratio (the hit rate) is computed as the number of replicates with a detected outcome per the total number of replicates tested. These hit rates are converted mathematically into cumulative normal probability units (probits) and fitted using a regression model *vs.* their respective concentrations. The LoD is defined as the lowest amount of viral genome that can be detected with a 95% hit rate. In the study, all viral loads strictly under 1 $\log_{10}$ copies/100 000 cells were considered under the LoD and were reported in the e-CRF as a negative result. If a rebound in viral excretion was observed, the sample was retested for confirmation.

#### Supplementary References

1 Etievant S, Bal A, Escuret V, *et al.* Performance Assessment of SARS-CoV-2 PCR Assays Developed by WHO Referral Laboratories. *J Clin Med* 2020; **9**. http://www.ncbi.nlm.nih.gov/pubmed/32560044.

**Supplementary Table S1. Baseline characteristics of participants included in the intention to treat population of the DisCoVeRy trial, overall and according to randomization arm and to disease severity at randomization.**
NPS, Nasopharyngeal swabs. * denotes variables with missing data.

| | Moderate | | | Severe | | |
|---|---|---|---|---|---|---|
| | **Overall (N=511)** | **Remdesivir (N=256)** | **Control (N=255)** | **Overall (N=332)** | **Remdesivir (N=164)** | **Control (N=168)** |
| **Median age** — yr [IQR] | 63 [53-73] | 62 [54-74] | 64 [52-73] | 65 [56-72] | 65 [56-73] | 66 [56-72] |
| **Median BMI** — kg/m2 [IQR] | 28 [25-33] | 28 [25-33] | 28 [25-32] | 29 [26-33] | 29 [26-33] | 29 [26-33] |
| **Male sex** — no. (%) | 340 (66.5%) | 166 (64.8%) | 174 (68.2%) | 246 (74.1%) | 128 (78.0%) | 118 (70.2%) |
| **Ethnicity*** — no. (%) | | | | | | |
| - Caucasian | 293 (63.3%) | 141 (60.8%) | 152 (65.8%) | 215 (77.6%) | 107 (78.7%) | 108 (76.6%) |
| - North African | 73 (15.8%) | 35 (15.1%) | 38 (16.5%) | 41 (14.8%) | 16 (11.8%) | 25 (17.7%) |
| - Sub-Saharan African | 37 (8.0%) | 25 (10.8%) | 12 (5.2%) | 10 (3.6%) | 5 (3.7%) | 5 (3.5%) |
| - Other | 60 (13.0%) | 31 (13.4%) | 29 (12.6%) | 11 (4.0%) | 8 (5.9%) | 3 (2.1%) |
| **Number of coexisting conditions*** — no. (%) | | | | | | |
| - 0 | 135 (26.5%) | 66 (25.9%) | 69 (27.1%) | 87 (26.4%) | 46 (28.4%) | 41 (24.4%) |
| - 1 | 177 (34.7%) | 90 (35.3%) | 87 (34.1%) | 104 (31.5%) | 54 (33.3%) | 50 (29.8%) |
| - 2 | 118 (23.1%) | 67 (26.3%) | 51 (20.0%) | 80 (24.2%) | 33 (20.4%) | 47 (28.0%) |
| - >2 | 80 (15.7%) | 32 (12.5%) | 48 (18.8%) | 59 (17.9%) | 29 (17.9%) | 30 (17.9%) |
| **Coexisting condition*** — no. (%) | | | | | | |
| - Obesity | 152 (30.2%) | 77 (30.8%) | 75 (29.6%) | 132 (40.0%) | 63 (38.9%) | 69 (41.1%) |
| - Chronic cardiac disease | 142 (27.8%) | 75 (29.4%) | 67 (26.3%) | 92 (28.0%) | 37 (23.0%) | 55 (32.7%) |
| - Diabetes mellitus | 140 (27.5%) | 71 (27.8%) | 69 (27.1%) | 85 (25.0%) | 38 (23.5%) | 47 (28.0%) |
| - Chronic pulmonary disease | 96 (18.8%) | 45 (17.6%) | 51 (20.0%) | 55 (16.7%) | 28 (17.4%) | 27 (16.1%) |
| - Chronic kidney disease (stage 1 to 3) | 31 (6.1%) | 12 (4.7%) | 19 (7.5%) | 24 (7.3%) | 9 (5.6%) | 15 (8.9%) |
| - Auto-inflammatory disease | 26 (5.1%) | 11 (4.3%) | 15 (5.9%) | 15 (4.6%) | 6 (3.7%) | 9 (5.4%) |
| - Malignant hemopathy | 17 (3.7%) | 6 (2.6%) | 11 (4.7%) | 19 (6.7%) | 11 (7.9%) | 8 (5.5%) |
| - Chronic neurological disorder (including dementia) | 24 (4.7%) | 10 (3.9%) | 14 (5.5%) | 13 (4.0%) | 9 (5.6%) | 4 (2.4%) |
| - Mild liver disease | 18 (3.5%) | 8 (3.1%) | 10 (3.9%) | 11 (3.3%) | 6 (3.7%) | 5 (3.0%) |
| - Active malignant neoplasm | 22 (4.3%) | 10 (3.9%) | 12 (4.7%) | 6 (1.8%) | 3 (1.9%) | 3 (1.8%) |
| - Transplantation | 5 (1.0%) | 1 (0.4%) | 4 (1.6%) | 6 (1.8%) | 1 (0.6%) | 5 (3.0%) |
| - Asplenia | 3 (0.6%) | 0 (0.0%) | 3 (1.2%) | 1 (0.3%) | 1 (0.6%) | 0 (0.0%) |
| - AIDS / HIV not on HAART | 2 (0.4%) | 0 (0.0%) | 2 (0.8%) | 0 (0.0%) | 0 (0.0%) | 0 (0.0%) |
| - Smoking status (current or former) | 86 (17.6%) | 47 (19.4%) | 39 (15.9%) | 58 (18.5%) | 28 (17.9%) | 30 (19.0%) |
| - Smoking status (current) | 19 (3.9%) | 10 (4.1%) | 9 (3.7%) | 13 (4.1%) | 5 (3.2%) | 8 (5.1%) |
| **Median time from symptoms onset to** | 8.0 [6.0-11.0] | 9.0 [6.0-11.0] | 8.0 [6.0-11.0] | 9.5 [7.0-12.0] | 9.0 [7.0-11.0] | 10.0 [7.0; 12.0] |

| | Moderate | | | Severe | | |
|---|---|---|---|---|---|---|
| | **Overall (N=511)** | **Remdesivir (N=256)** | **Control (N=255)** | **Overall (N=332)** | **Remdesivir (N=164)** | **Control (N=168)** |
| **randomization*** — days [IQR] | | | | | | |
| **Severity of COVID-19 at randomization** — no. (%) | | | | | | |
| - Moderate | 511 (100.0%) | 256 (100.0%) | 255 (100.0%) | 0 (0.0%) | 0 (0.0%) | 0 (0.0%) |
| - Severe | 0 (0.0%) | 0 (0.0%) | 0 (0.0%) | 332 (100.0%) | 164 (100.0%) | 168 (100.0%) |
| **Ventilatory support at randomization** — no. (%) | | | | | | |
| - Room air | 12 (2.3%) | 6 (2.3%) | 6 (2.4%) | 0 (0.0%) | 0 (0.0%) | 0 (0.0%) |
| - Oxygen support (Nasal cannula, face mask) | 499 (97.7%) | 250 (97.7%) | 249 (97.6%) | 0 (0.0%) | 0 (0.0%) | 0 (0.0%) |
| - High Flow Oxygen device (e.g. optiflow) | 0 (0.0%) | 0 (0.0%) | 0 (0.0%) | 149 (44.9%) | 72 (43.9%) | 77 (45.8%) |
| - Non-Invasive ventilation | 0 (0.0%) | 0 (0.0%) | 0 (0.0%) | 32 (9.6%) | 15 (9.1%) | 17 (10.1%) |
| - Invasive Mechanical Ventilation | 0 (0.0%) | 0 (0.0%) | 0 (0.0%) | 149 (44.9%) | 77 (47.0%) | 72 (42.9%) |
| - ECMO | 0 (0.0%) | 0 (0.0%) | 0 (0.0%) | 2 (0.6%) | 0 (0.0%) | 2 (1.2%) |
| **NEWS-2*** — no. (%) | 7.0 [6.0-9.0] | 7.0 [5.0-9.0] | 7.0 [6.0-9.0] | 10.0 [9.0-13.0] | 10.0 [8.0-13.0] | 10.0 [9.0-13.0] |
| **7-point ordinal scale at baseline** — no. (%) | | | | | | |
| 3. Hospitalized, not requiring supplemental oxygen | 15 (2.9%) | 8 (3.1%) | 7 (2.7%) | 0 (0.0%) | 0 (0.0%) | 0 (0.0%) |
| 4. Hospitalized, requiring supplemental oxygen | 471 (92.2%) | 235 (91.8%) | 236 (92.5%) | 11 (3.3%) | 5 (3.0%) | 6 (3.6%) |
| 5. Hospitalized, on non-invasive ventilation or high flow oxygen devices | 23 (4.5%) | 12 (4.7%) | 11 (4.3%) | 169 (50.9%) | 82 (50.0%) | 87 (51.8%) |
| 6. Hospitalized, on invasive mechanical ventilation or ECMO | 2 (0.4%) | 1 (0.4%) | 1 (0.4%) | 152 (45.8%) | 77 (47.0%) | 75 (44.6%) |
| **Randomization site*** — no. (%) | | | | | | |
| - ICU | 50 (9.8%) | 28 (11.0%) | 22 (8.6%) | 318 (96.4%) | 156 (96.3%) | 162 (96.4%) |
| - Conventional unit | 460 (90.2%) | 227 (89.0%) | 233 (91.4%) | 12 (3.6%) | 6 (3.7%) | 6 (3.6%) |
| **Median viral load on NPS at baseline*** – $log_{10}$ cp/10,000 cells [IQR] | 3.1 [1.7-4.4] | 3.2 [1.7-4.5] | 2.8 [1.7-4.3] | 3.4 [1.9-4.7] | 3.3 [1.8-4.5] | 3.6 [2.1-4.9] |
| **Biological data at baseline*** – median [IQR] | | | | | | |
| - Minimal lymphocyte count ($10^9$/L) | 0.9 [0.7-1.3] | 0.9 [0.6-1.2] | 1.0 [0.7-1.4] | 0.7 [0.5-0.9] | 0.7 [0.5-1.0] | 0.7 [0.5; 0.9] |
| - Maximal neutrophil count ($10^9$/L) | 4.8 [3.3-7.3] | 4.7 [3.3-7.8] | 4.8 [3.2-6.8] | 7.1 [5.1-9.8] | 7.2 [5.8-9.7] | 6.7 [4.6-10.0] |
| - Maximal platelet count ($10^9$/L) | 209.0 [163.0-286.5] | 210.0 [168.0-295.0] | 205.0 [159.0-286.0] | 244.0 [183.0-306.0] | 251.0 [179.0-322.0] | 244.0 [185.0-298.0] |
| - Maximal urea (mmol/L) | 6.0 [4.0-8.0] | 6.0 [4.0-8.0] | 6.0 [4.0-8.0] | 8.0 [5.5-11.0] | 8.0 [6.0-11.0] | 8.0 [5.0-11.0] |
| - Maximal creatininemia (µmol/L) | 75.0 [61.0-91.0] | 75.0 [60.0-92.0] | 75.0 [61.0-91.0] | 74.0 [61.0-98.0] | 72.0 [60.0-98.0] | 74.0 [61.0-98.5] |
| - Maximal AST / SGOT (U/L) | 44.0 [32.0-65.0] | 44.0 [32.0-67.5] | 44.0 [31.0-60.0] | 49.0 [36.0-72.5] | 49.0 [35.0-69.0] | 50.0 [36.5-73.0] |
| - Maximal ALT / SGPT (U/L) | 36.0 [23.0-58.0] | 36.0 [24.0-55.0] | 37.0 [23.0-59.0] | 38.0 [24.0-58.0] | 36.5 [23.0-53.0] | 40.0 [26.0-64.0] |
| - Maximal Total Bilirubin (µmol/L) | 8.6 [6.0-12.0] | 8.6 [6.0-12.0] | 8.7 [6.2-12.0 | 8.7 [6.0-12.9] | 8.6 [6.0-12.0] | 8.7 [6.0-13.0] |
| - Maximal International Normalized Ratio | 1.1 [1.0-1.2] | 1.1 [1.0-1.2] | 1.1 [1.0-1.2] | 1.1 [1.0-1.2] | 1.1 [1.0-1.2] | 1.1 [1.1-1.3] |
| - Maximal C-Reactive Protein (mg/L) | 91.0 [49.0-151.0] | 89.0 [48.0-144.0] | 96.0 [49.0-155.0] | 122.0 [73.0-192.0] | 118.0 [73.0-179.0] | 129.5 [72.5-203.5] |

| | **Moderate** | | | **Severe** | | |
|---|---|---|---|---|---|---|
| | **Overall (N=511)** | **Remdesivir (N=256)** | **Control (N=255)** | **Overall (N=332)** | **Remdesivir (N=164)** | **Control (N=168)** |
| - Maximal D-Dimers (µg/L) | 829.0 [540.0-1320.0] | 800.0 [499.0-1230.0] | 930.0 [591.0-1583.0] | 1170.0 [620.0-2405.0] | 1208.5 [681.0-2277.5] | 1080.0 [620.0-2680.0] |
| - Maximal Procalcitonin (g/mL) | 0.2 [0.1-0.4] | 0.2 [0.1-0.4] | 0.2 [0.1-0.4] | 0.3 [0.1-1.2] | 0.3 [0.1-1.0] | 0.4 [0.2-1.6] |
| - Maximal Ferritin max - mg/L) | 632.0 [286.0-1334.0] | 886.0 [407.0-1763.0] | 610.5 [99.0-1262.5] | 962.0 [418.0-1883.0] | 1088.5 [546.5-2250.5] | 886.0 [407.0-1763.0] |

**Supplementary Table S2. Concomitant treatments received during the study course in the patients included in the intention-to-treat population of the DisCoVeRy trial, overall, according to randomization arm and to disease severity at randomization.**

| | All patients | | | Moderate | | | Severe | | |
|---|---|---|---|---|---|---|---|---|---|
| | **Overall (N=843)** | **Remdesivir (N=420)** | **Control (N=423)** | **Overall (N=511)** | **Remdesivir (N=256)** | **Control (N=255)** | **Overall (N=332)** | **Remdesivir (N=164)** | **Control (N=168)** |
| **Corticosteroids (general route)** — no. (%) | 296 (35.1%) | 147 (35.0%) | 149 (35.2%) | 171 (33.5%) | 82 (32.0%) | 89 (34.9%) | 125 (37.7%) | 65 (39.6%) | 60 (35.7%) |
| - Dexamethasone | 227 (26.9%) | 119 (28.3%) | 108 (25.5%) | 131 (25.6%) | 69 (27.0%) | 62 (24.3%) | 96 (28.9%) | 50 (30.5%) | 46 (27.4%) |
| - Hydrocortisone | 22 (2.6%) | 7 (1.7%) | 15 (3.5%) | 8 (1.6%) | 3 (1.2%) | 5 (2.0%) | 14 (4.2%) | 4 (2.4%) | 10 (6.0%) |
| - Methylprednisolone | 46 (5.5%) | 22 (5.2%) | 24 (5.7%) | 24 (4.7%) | 9 (3.5%) | 15 (5.9%) | 22 (6.6%) | 13 (7.9%) | 9 (5.4%) |
| **Corticosteroids (inhaled route)** — no. (%) | 62 (7.4%) | 29 (6.9%) | 33 (7.8%) | 43 (8.4%) | 22 (8.6%) | 21 (8.2%) | 19 (5.7%) | 7 (4.3%) | 12 (7.1%) |
| **Interleukin-6 inhibitors** — no. (%) | 7 (0.8%) | 5 (1.2%) | 2 (0.5%) | 5 (1.0%) | 4 (1.6%) | 1 (0.4%) | 2 (0.6%) | 1 (0.6%) | 1 (0.6%) |
| - Tocilizumab | 7 (0.8%) | 5 (1.2%) | 2 (0.5%) | 5 (1.0%) | 4 (1.6%) | 1 (0.4%) | 2 (0.6%) | 1 (0.6%) | 1 (0.6%) |
| **Interleukin-1 inhibitors** — no. (%) | 4 (0.5%) | 3 (0.7%) | 1 (0.2%) | 3 (0.6%) | 2 (0.8%) | 1 (0.4%) | 1 (0.3%) | 1 (0.6%) | 0 (0.0%) |
| **Angiotensin-receptor blockers** — no. (%) | 81 (9.6%) | 36 (8.6%) | 45 (10.6%) | 54 (10.6%) | 25 (9.8%) | 29 (11.4%) | 27 (8.1%) | 11 (6.7%) | 16 (9.5%) |
| **Antibiotics** — no. (%) | 354 (42.0%) | 181 (43.1%) | 173 (40.9%) | 193 (37.8%) | 97 (37.9%) | 96 (37.6%) | 161 (48.5%) | 84 (51.2%) | 77 (45.8%) |
| - Azithromycin | 15 (1.8%) | 11 (2.6%) | 4 (0.9%) | 14 (2.7%) | 10 (3.9%) | 4 (1.6%) | 1 (0.3%) | 1 (0.6%) | 0 (0.0%) |
| **Anticoagulants** — no. (%) | 446 (52.9%) | 216 (51.4%) | 230 (54.4%) | 271 (53.0%) | 134 (52.3%) | 137 (53.7%) | 175 (52.7%) | 82 (50.0%) | 93 (55.4%) |
| **Parenteral or enteral nutrition** — no. (%) | 271 (32.1%) | 128 (30.5%) | 143 (33.8%) | 71 (13.9%) | 34 (13.3%) | 37 (14.5%) | 200 (60.2%) | 94 (57.3%) | 106 (63.1%) |
| **Vasopressors** — no. (%) | 229 (27.2%) | 107 (25.5%) | 122 (28.8%) | 47 (9.2%) | 24 (9.4%) | 23 (9.0%) | 182 (54.8%) | 83 (50.6%) | 99 (58.9%) |
| **Extra-renal replacement/hemofiltration** — no. (%) | 34 (4.0%) | 10 (2.4%) | 24 (5.7%) | 8 (1.6%) | 3 (1.2%) | 5 (2.0%) | 26 (7.8%) | 7 (4.3%) | 19 (11.3%) |
| **Neuromuscular blocking agents** — no. (%) | 213 (25.3%) | 96 (22.9%) | 117 (27.7%) | 48 (9.4%) | 22 (8.6%) | 26 (10.2%) | 165 (49.7%) | 74 (45.1%) | 91 (54.2%) |
| **Inhaled nitric oxide** — no. (%) | 32 (3.8%) | 15 (3.6%) | 17 (4.0%) | 8 (1.6%) | 4 (1.6%) | 4 (1.6%) | 24 (7.2%) | 11 (6.7%) | 13 (7.7%) |
| **Prone positioning** — no. (%) | 200 (23.7%) | 96 (22.9%) | 104 (24.6%) | 44 (8.6%) | 24 (9.4%) | 20 (7.8%) | 156 (47.0%) | 72 (43.9%) | 84 (50.0%) |

**Supplementary Table S3. Administration of corticosteroids by general route, before and after June, 30th 2020 in the intention-to-treat population of the DisCoVeRy trial, overall and according to randomization arm and to disease severity at randomization.**
On June 30th, 2020, results from the RECOVERY trial reported the efficacy of dexamethasone in reducing mortality of patients hospitalized with COVID-19.

| **Corticosteroids (general route)** — no. (%) | **All patients** | | | **Moderate** | | | **Severe** | | |
|---|---|---|---|---|---|---|---|---|---|
| | **Overall (N=843)** | **Remdesivir (N=420)** | **Control (N=423)** | **Overall (N=511)** | **Remdesivir (N=256)** | **Control (N=255)** | **Overall (N=332)** | **Remdesivir (N=164)** | **Control (N=168)** |
| Patients randomized before June 30, 2020 | 80/301 (26.6%) | 39/152 (25.7%) | 41/149 (27.5%) | 43/190 (22.6%) | 20/95 (21.1%) | 23/95 (24.2%) | 37/111 (33.3%) | 19/57 (33.3%) | 18/54 (33.3%) |
| Patients randomized after June 30, 2020 | 216/542 (39.9%) | 110/271 (40.6%) | 108/274 (39.4%) | 128/321 (39.9%) | 62/161 (38.5%) | 66/160 (41.3%) | 88/221 (39.8%) | 46/107 (43.0%) | 42/114 (36.8%) |

**Supplementary Table S4. Secondary outcomes in the intention-to-treat population of the DisCoVeRy trial, overall, according to randomization arm and to severity at randomization.**

Analyses were stratified on the disease severity at randomization and adjusted effect measures are reported in the table. ECMO, extracorporeal membrane oxygenation; OR, Odds-ratio; HR, Hazard ratio; LSMD, least-square mean difference. Estimates are reported with their 95% confidence interval.

| | **Overall (N=843)** | | **Moderate (N=511)** | | **Severe (N=332)** | | **Remdesivir vs. control Effect measure (95%CI)** |
|---|---|---|---|---|---|---|---|
| | **Remdesivir (N=420)** | **Control (N=423)** | **Remdesivir (N=256)** | **Control (N=255)** | **Remdesivir (N=164)** | **Control (N=168)** | |
| **7-point ordinal scale at day 3** — no. (%) | | | | | | | |
| 3. Hospitalized, not requiring supplemental oxygen | 22 (5.2%) | 37 (8.7%) | 22 (8.6%) | 37 (14.5%) | 0 (0.0%) | 0 (0.0%) | OR=0.95 (0.72 to 1.26) [P=0.72] |
| 4. Hospitalized, requiring supplemental oxygen | 192 (45.7%) | 184 (43.5%) | 183 (71.5%) | 174 (68.2%) | 9 (5.5%) | 10 (6.0%) | |
| 5. Hospitalized, on non-invasive ventilation or high flow oxygen devices | 102 (24.3%) | 79 (18.7%) | 37 (14.5%) | 27 (10.6%) | 65 (39.6%) | 52 (31.0%) | |
| 6. Hospitalized, on invasive mechanical ventilation or ECMO | 102 (24.3%) | 120 (28.4%) | 14 (5.5%) | 17 (6.7%) | 88 (53.7%) | 103 (61.3%) | |
| 7. Death | 2 (0.5%) | 3 (0.7%) | 0 (0.0%) | 0 (0.0%) | 2 (1.2%) | 3 (1.8%) | |
| **7-point ordinal scale at day 5** — no. (%) | | | | | | | |
| 1. Not hospitalized, no limitations on activities | 1 (0.2%) | 5 (1.2%) | 1 (0.4%) | 5 (2.0%) | 0 (0.0%) | 0 (0.0%) | OR=1.07 (0.83 to 1.39) [P=0.59] |
| 2. Not hospitalized, limitation on activities | 1 (0.2%) | 14 (3.3%) | 1 (0.4%) | 14 (5.5%) | 0 (0.0%) | 0 (0.0%) | |
| 3. Hospitalized, not requiring supplemental oxygen | 71 (16.9%) | 58 (13.7%) | 65 (25.4%) | 53 (20.8%) | 6 (3.7%) | 5 (3.0%) | |
| 4. Hospitalized, requiring supplemental oxygen | 159 (37.9%) | 155 (36.6%) | 138 (53.9%) | 134 (52.5%) | 21 (12.8%) | 21 (12.5%) | |
| 5. Hospitalized, on non-invasive ventilation or high flow oxygen devices | 77 (18.3%) | 57 (13.5%) | 31 (12.1%) | 27 (10.6%) | 46 (28.0%) | 30 (17.9%) | |
| 6. Hospitalized, on invasive mechanical ventilation or ECMO | 106 (25.2%) | 125 (29.6%) | 18 (7.0%) | 20 (7.8%) | 88 (53.7%) | 105 (62.5%) | |
| 7. Death | 5 (1.2%) | 9 (2.1%) | 2 (0.8%) | 2 (0.8%) | 3 (1.8%) | 7 (4.2%) | |
| **7-point ordinal scale at day 8** — no. (%) | | | | | | | |
| 1. Not hospitalized, no limitations on activities | 13 (3.1%) | 23 (5.4%) | 12 (4.7%) | 23 (9.0%) | 1 (0.6%) | 0 (0.0%) | OR=0.92 (0.72 to 1.17) [P=0.50] |
| 2. Not hospitalized, limitation on activities | 39 (9.3%) | 58 (13.7%) | 36 (14.1%) | 53 (20.8%) | 3 (1.8%) | 5 (3.0%) | |
| 3. Hospitalized, not requiring supplemental oxygen | 81 (19.3%) | 70 (16.5%) | 68 (26.6%) | 56 (22.0%) | 13 (7.9%) | 14 (8.3%) | |
| 4. Hospitalized, requiring supplemental oxygen | 141 (33.6%) | 112 (26.5%) | 102 (39.8%) | 80 (31.4%) | 39 (23.8%) | 32 (19.0%) | |
| 5. Hospitalized, on non-invasive ventilation or high flow oxygen devices | 42 (10.0%) | 35 (8.3%) | 16 (6.3%) | 16 (6.3%) | 26 (15.9%) | 19 (11.3%) | |
| 6. Hospitalized, on invasive mechanical ventilation or ECMO | 93 (22.1%) | 112 (26.5%) | 18 (7.0%) | 22 (8.6%) | 75 (45.7%) | 90 (53.6%) | |
| 7. Death | 11 (2.6%) | 13 (3.1%) | 4 (1.6%) | 5 (2.0%) | 7 (4.3%) | 8 (4.8%) | |
| **7-point ordinal scale at day 11** — no. (%) | | | | | | | |
| 1. Not hospitalized, no limitations on activities | 25 (6.0%) | 34 (8.0%) | 20 (7.8%) | 30 (11.8%) | 5 (3.0%) | 4 (2.4%) | OR=0.97 (0.76 to 1.23) [P=0.79] |
| 2. Not hospitalized, limitation on activities | 91 (21.7%) | 114 (27.0%) | 78 (30.5%) | 99 (38.8%) | 13 (7.9%) | 15 (8.9%) | |
| 3. Hospitalized, not requiring supplemental oxygen | 83 (19.8%) | 49 (11.6%) | 65 (25.4%) | 35 (13.7%) | 18 (11.0%) | 14 (8.3%) | |
| 4. Hospitalized, requiring supplemental oxygen | 103 (24.5%) | 83 (19.6%) | 65 (25.4%) | 49 (19.2%) | 38 (23.2%) | 34 (20.2%) | |
| 5. Hospitalized, on non-invasive ventilation or high flow oxygen devices | 25 (6.0%) | 24 (5.7%) | 5 (2.0%) | 12 (4.7%) | 20 (12.2%) | 12 (7.1%) | |
| 6. Hospitalized, on invasive mechanical ventilation or ECMO | 78 (18.6%) | 99 (23.4%) | 17 (6.6%) | 20 (7.8%) | 61 (37.2%) | 79 (47.0%) | |
| 7. Death | 15 (3.6%) | 20 (4.7%) | 6 (2.3%) | 10 (3.9%) | 9 (5.5%) | 10 (6.0%) | |

| | **Overall (N=843)** | | **Moderate (N=511)** | | **Severe (N=332)** | | **Remdesivir vs. control Effect measure (95%CI)** |
|---|---|---|---|---|---|---|---|
| | **Remdesivir (N=420)** | **Control (N=423)** | **Remdesivir (N=256)** | **Control (N=255)** | **Remdesivir (N=164)** | **Control (N=168)** | |
| **Time to improvement of 1 category of the 7-point ordinal scale within day 29** — median (IQR) | 12 [8-24] | 12 [7-28] | 11 [8-20] | 9 [6-16] | 16 [10-29] | 18 [10-29] | HR=1.08 (0.93 to 1.26) [P=0.30] |
| **Change from baseline in the ordinal scale** — median (IQR) | | | | | | | |
| – to day 3 | 0 [0-0] | 0 [0-0] | 0 [0-0] | 0 [0-0] | 0 [0-0] | 0 [0-0] | LSMD=0.00 (-0.08 to 0.08) [P=0.98] |
| – to day 5 | 0 [0-0] | 0 [0-0] | 0 [-1-0] | 0 [-1-0] | 0 [0-0] | 0 [0-1] | LSMD=-0.00 (-0.12 to 0.12) [P=0.94] |
| – to day 8 | 0 [-1-0] | 0 [-2-0] | 0 [-1-0] | -1 [-2-0] | 0 [-1-0] | 0 [-1-1] | LSMD=0.05 (-0.12 to 0.22) [P=0.58] |
| – to day 11 | -1 [-2-0] | -1 [-2-0] | -1 [-2-0] | -2 [-2-0] | -1 [-2-0] | 0 [-2-1] | LSMD=-0.05 (-0.25 to 0.15) [P=0.60] |
| – to day 15 | -2 [-2-0] | -2 [-3-0] | -2 [-2-0] | -2 [-3-0] | -1 [-3-0] | -1 [-3-0] | LSMD=-0.05 (-0.27 to 0.18) [P=0.68] |
| – to day 29 | -2 [-3--1] | -2 [-3--1] | -2 [-3--1] | -2 [-3--1] | -3 [-4-0] | -2 [-3-0] | LSMD=-0.18 (-0.42 to 0.06) [P=0.14] |
| **Change from baseline in NEWS2** — median (IQR) | | | | | | | |
| – to day 5 | -1 [-3-1] | -1 [-3-1] | -1 [-4-1] | -1 [-4-1] | 0 [-2-2] | 0 [-2-2] | LSMD=0.04 (-0.48 to 0.57) [P=0.87] |
| – to day 11 | -3 [-5-1] | -2 [-4-1] | -3 [-5-0] | -2 [-5-0] | -1 [-4-2] | -1 [-4-3] | LSMD=-0.42 (-1.09 to 0.25) [P=0.22] |
| – to day 15 | -3 [-6-0] | -3 [-6-0] | -4 [-6--1] | -4 [-6--1] | -2 [-6-2] | -1 [-5-3] | LSMD=-0.37 (-1.11 to 0.36) [P=0.32] |
| – to day 29 | -4 [-7--1] | -4 [-7-0] | -4 [-7--2] | -4 [-7--2] | -4 [-8-0] | -3 [-7-2] | LSMD=-0.37 (-1.20 to 0.47) [P=0.39] |

**Supplementary Table S5. Proportion of patients with a detectable viral load, normalized viral load, change from baseline of normalized viral loads in the nasopharyngeal swabs at each sampling time in the intention-to-treat population of the DisCoVeRy trial, overall, according to randomization arm and to disease severity at randomization.**
Analyses of detectable viral load were stratified on the disease severity at randomization and reported OR are adjusted on disease severity.
NP, nasopharyngeal; OR, odds ratio.

| | All patients | | | Moderate | | | Severe | | | Remdesivir vs. control Effect measure (95%CI) |
|---|---|---|---|---|---|---|---|---|---|---|
| | Overall (N=843) | Remdesivir (N=420) | Control (N=423) | Overall (N=511) | Remdesivir (N=256) | Control (N=255) | Overall (N=332) | Remdesivir (N=164) | Control (N=168) | |
| **Detectable viral load in NP swabs** – n/N (%) | | | | | | | | | | |
| - at baseline | 388/447 (86.8%) | 199/235 (84.7%) | 189/212 (89.2%) | 238/280 (85.0%) | 122/146 (83.6%) | 116/134 (86.6%) | 150/167 (89.8%) | 77/89 (86.5%) | 73/78 (93.6%) | - |
| - at day 3 | 387/509 (76.0%) | 201/259 (77.6%) | 186/250 (74.4%) | 237/320 (74.1%) | 130/167 (77.8%) | 107/153 (69.9%) | 150/189 (79.4%) | 71/92 (77.2%) | 79/97 (81.4%) | OR=1.20 (0.80 to 1.81) [P=0.37] |
| - at day 5 | 321/489 (65.6%) | 165/252 (65.5%) | 156/237 (65.8%) | 196/315 (62.2%) | 102/165 (61.8%) | 94/150 (62.7%) | 125/174 (71.8%) | 63/87 (72.4%) | 62/87 (71.3%) | OR=0.99 (0.68 to 1.45) [P=0.97] |
| - at day 8 | 209/419 (49.9%) | 109/218 (50.0%) | 100/201 (49.8%) | 123/248 (49.6%) | 67/133 (50.4%) | 56/115 (48.7%) | 86/171 (50.3%) | 42/85 (49.4%) | 44/86 (51.2%) | OR=1.01 (0.69 to 1.48) [P=0.96] |
| - at day 11 | 134/309 (43.4%) | 77/171 (45.0%) | 57/138 (41.3%) | 72/164 (43.9%) | 45/95 (47.4%) | 27/69 (39.1%) | 62/145 (42.8%) | 32/76 (42.1%) | 30/69 (43.5%) | OR=1.16 (0.74 to 1.83) [P=0.52] |
| - at day 15 | 142/450 (31.6%) | 66/226 (29.2%) | 76/224 (33.9%) | 92/295 (31.2%) | 44/149 (29.5%) | 48/146 (32.9%) | 50/155 (32.3%) | 22/77 (28.6%) | 28/78 (35.9%) | OR=0.80 (0.54 to 1.20) [P=0.28] |
| - at day 29 | 36/358 (10.1%) | 17/174 (9.8%) | 19/184 (10.3%) | 25/254 (9.8%) | 11/123 (8.9%) | 14/131 (10.7%) | 11/104 (10.6%) | 6/51 (11.8%) | 5/53 (9.4%) | OR=0.94 (0.47 to 1.87) [P=0.86] |
| **Median viral load in NP swabs**, $log_{10}$ cp/10,000 cells [IQR] | | | | | | | | | | |
| - at baseline | 3.2 [1.8-4.5] | 3.2 [1.7-4.5] | 3.2 [1.8-4.4] | 3.1 [1.7-4.4] | 3.2 [1.7-4.5] | 2.8 [1.7-4.3] | 3.4 [1.9-4.7] | 3.3 [1.8-4.5] | 3.6 [2.1-4.9] | |
| - at day 3 | 2.3 [1.0-3.7] | 2.4 [1.2-3.3] | 2.2 [0.7-3.9] | 2.2 [0.7-3.5] | 2.3 [1.2-3.2] | 2.2 [0.7-3.7] | 2.5 [1.2-3.9] | 2.5 [1.1-3.6] | 2.5 [1.2-4.2] | |
| - at day 5 | 1.7 [0.7-3.1] | 1.6 [0.7-2.9] | 1.8 [0.7-3.5] | 1.6 [0.7-3.0] | 1.5 [0.7-2.9] | 1.6 [0.7-3.4] | 1.9 [0.7-3.2] | 1.8 [0.7-2.9] | 2.1 [0.7-3.7] | |
| - at day 8 | 1.0 [0.7-2.2] | 1.0 [0.7-2.2] | 0.7 [0.7-2.2] | 1.0 [0.7-2.2] | 1.0 [0.7-2.2] | 0.7 [0.7-2.2] | 0.7 [0.7-2.2] | 0.9 [0.7-2.2] | 0.7 [0.7-2.3] | |
| - at day 11 | 0.7 [0.7-2.0] | 0.7 [0.7-1.9] | 0.7 [0.7-2.0] | 0.7 [0.7-2.0] | 0.7 [0.7-2.0] | 0.7 [0.7-1.9] | 0.7 [0.7-2.0] | 0.7 [0.7-1.9] | 0.7 [0.7-2.1] | |
| - at day 15 | 0.7 [0.7-1.2] | 0.7 [0.7-1.0] | 0.7 [0.7-1.3] | 0.7 [0.7-1.2] | 0.7 [0.7-1.0] | 0.7 [0.7-1.2] | 0.7 [0.7-1.3] | 0.7 [0.7-1.0] | 0.7 [0.7-1.4] | |

| | All patients | | | Moderate | | | Severe | | | Remdesivir vs. control Effect measure (95%CI) |
|---|---|---|---|---|---|---|---|---|---|---|
| | **Overall (N=843)** | **Remdesivir (N=420)** | **Control (N=423)** | **Overall (N=511)** | **Remdesivir (N=256)** | **Control (N=255)** | **Overall (N=332)** | **Remdesivir (N=164)** | **Control (N=168)** | |
| **Change from baseline of the normalized viral load in NP swabs**, $\log_{10}$ cp/10,000 cells [IQR] | | | | | | | | | | |
| - at day 3 | -0.5 [-1.3-0.0] | -0.5 [-1.4-0.0] | -0.5 [-1.3-0.1] | -0.5 [-1.2-0.0] | -0.6 [-1.5-0.0] | -0.4 [-1.2-0.0] | -0.5 [-1.4-0.3] | -0.4 [-1.4-0.3] | -0.6 [-1.5-0.2] | |
| - at day 5 | -1.0 [-1.9-0.0] | -1.1 [-1.9-0.0] | -1.0 [-1.9-0.0] | -1.0 [-1.8-0.0] | -1.1 [-1.9-0.0] | -0.9 [-1.8-0.0] | -1.1 [-2.1-0.0] | -1.0 [-1.9-0.0] | -1.1 [-2.2--0.4] | |
| - at day 8 | -1.6 [-2.6--0.4] | -1.6 [-2.6--0.4] | -1.6 [-2.5--0.3] | -1.5 [-2.5--0.3] | -1.5 [-2.4--0.5] | -1.5 [-2.5--0.2] | -1.8 [-2.8--0.4] | -1.8 [-2.9--0.3] | -1.8 [-2.7--0.4] | |
| - at day 11 | -1.9 [-3.0--0.5] | -2.0 [-3.0--0.6] | -1.8 [-3.0--0.5] | -1.9 [-2.8--0.6] | -2.0 [-2.8--0.6] | -1.5 [-3.1--0.5] | -1.9 [-3.2--0.5] | -1.9 [-3.2--0.4] | -1.9 [-2.8--0.5] | |
| - at day 15 | -1.9 [-3.2--0.7] | -2.0 [-3.2--0.7] | -1.8 [-3.3--0.9] | -1.7 [-3.2--0.5] | -1.8 [-3.1--0.4] | -1.6 [-3.2--0.7] | -2.4 [-3.6--1.1] | -2.4 [-3.2--1.2] | -2.4 [-3.7--1.1] | |

**Supplementary Table S6. Estimated intercepts and slopes of viral load decrease and difference of slopes between randomization arms, according to disease severity at randomization.** Analyses were stratified on the disease severity at randomization and reported effect measure is adjusted on disease severity at randomization. Intercepts and slopes are reported with their standard errors; difference of slopes is reported with its 95% confidence interval.

| | **Remdesivir** | | **Control** | | **Remdesivir vs. control Effect measure (95%CI)** |
|---|---|---|---|---|---|
| | Moderate | Severe | Moderate | Severe | |
| Intercept, $log_{10}$ cp/10,000 cells | 2.85 (0.11) | 3.01 (0.13) | 3.04 (0.11) | 3.18 (0.13) | - |
| Slope, $log_{10}$ cp/10,000 cells/day | 0.13 (0.01) | 0.15 (0.01) | 0.14 (0.01) | 0.15 (0.01) | -0.006 (-0.02 to 0.03) [P=0.66] |

**Supplementary Table S7. Estimated intercepts and slopes of viral load decrease and difference of slopes between randomization arms, according to disease severity at randomization and to the duration of symptoms before randomization.**
Intercepts and slopes are reported with their standard errors; differences of slopes are reported with their 95% confidence interval.

| | | Remdesivir | Control | Remdesivir vs. control Effect measure (95%CI) |
|---|---|---|---|---|
| **Disease Severity** | | | | |
| Moderate | Intercept, $log_{10}$ cp/10,000 cells | 2.90 (0.12) | 2.97 (0.13) | - |
| | Slope, $log_{10}$ cp/10,000 cells/day | 0.13 (0.01) | 0.13 (0.01) | 0.002 (-0.03 to 0.04) [P=0.93] |
| Severe | Intercept, $log_{10}$ cp/10,000 cells | 2.93 (0.15) | 3.26 (0.15) | - |
| | Slope, $log_{10}$ cp/10,000 cells/day | 0.14 (0.01) | 0.15 (0.01) | -0.013 (-0.03 to 0.05) [P=0.54] |
| **Duration of symptoms prior to randomization** | | | | |
| ≤ 7 days | Intercept, $log_{10}$ cp/10,000 cells | 3.33 (0.18) | 3.82 (0.18) | - |
| | Slope, $log_{10}$ cp/10,000 cells/day | 0.16 (0.02) | 0.18 (0.02) | -0.014 (-0.03 to 0.06) [P=0.55] |
| 7 – 14 days | Intercept, $log_{10}$ cp/10,000 cells | 2.67 (0.11) | 2.71 (0.12) | - |
| | Slope, $log_{10}$ cp/10,000 cells/day | 0.12 (0.01) | 0.12 (0.01) | -0.006 (-0.03 to 0.04) [P=0.71] |
| >14 days | Intercept, $log_{10}$ cp/10,000 cells | 3.0 (0.41) | 2.61 (0.34) | - |
| | Slope, $log_{10}$ cp/10,000 cells/day | 0.14 (0.05) | 0.10 (0.04) | 0.033 (-0.16 to 0.09) [P=0.60] |

**Supplementary Table S8. Plasma concentrations of remdesivir and its metabolite GS-441524, overall and according to disease severity at randomization, in remdesivir-treated participants from the DisCoVeRy trial.**

| | | Overall | Moderate | Severe |
|---|---|---|---|---|
| **Median post-infusion plasma concentration** — ng/mL [IQR] | | | | |
| at day 1 | - n | 58 | 44 | 14 |
| | - remdesivir | 2,541 [1,417; 4,845] | 2,387 [806; 4,682] | 2,617 [2,240; 5,180] |
| | - GS-441524 | 74 [51; 105] | 72 [51; 103] | 81 [59; 107] |
| **Median trough plasma concentration** — ng/mL [IQR] | | | | |
| at day 2 | - n | 49 | 37 | 12 |
| | - GS-441524 | 66 [50; 102] | 67 [51; 105] | 63 [50; 95] |
| at day 5 | - n | 32 | 21 | 11 |
| | - GS-441524 | 59 [46; 86] | 72 [56; 94] | 53 [42; 69] |
| at day 8 | - n | 22 | 11 | 11 |
| | - GS-441524 | 63 [44; 87] | 70 [62; 121] | 49 [37; 68] |

**Supplementary Table S9. Summary of adverse events according to treatment group in the modified intention-to-treat population of the DisCoVeRy trial, overall and according to randomization arm and to disease severity at randomization.**

Numbers refer to number of patients (%). Some patients had more than a single AE. Analyses were performed on the modified Intention-to-treat population.

| no (%) | Remdesivir (n=410) | Control (n=423) |
|---|---|---|
| **Blood and lymphatic system disorders** | 11 (3%) | 13 (3%) |
| - Anaemia | 8 (2%) | 11 (2%) |
| - Haemolytic anaemia | 1 (0%) | 0 (0%) |
| - Lymphopenia | 1 (0%) | 1 (0%) |
| - Thrombocytopenia | 0 (0%) | 1 (0%) |
| - Thrombocytosis | 1 (0%) | 0 (0%) |
| **Cardiac disorders** | 13 (3%) | 19 (4%) |
| - Acute coronary syndrome | 0 (0%) | 1 (0%) |
| - Arrhythmia | 5 (1%) | 3 (1%) |
| - Bradycardia | 3 (1%) | 2 (0%) |
| - Cardiac arrest | 1 (0%) | 4 (1%) |
| - Cardiac failure | 0 (0%) | 3 (1%) |
| - Congestive cardiomyopathy | 0 (0%) | 1 (0%) |
| - Left ventricular failure | 0 (0%) | 0 (0%) |
| - Myocardial ischaemia | 1 (0%) | 0 (0%) |
| - Myocarditis | 0 (0%) | 1 (0%) |
| - Tachycardia | 4 (1%) | 2 (0%) |
| - Ventricular tachycardia | 0 (0%) | 2 (0%) |
| **Gastrointestinal disorders** | 3 (1%) | 7 (2%) |
| - Bezoar | 0 (0%) | 1 (0%) |
| - Faecaloma | 1 (0%) | 0 (0%) |
| - Gastrointestinal haemorrhage | 0 (0%) | 3 (1%) |
| - Gastrointestinal ulcer | 0 (0%) | 2 (0%) |
| - Intestinal ischaemia | 1 (0%) | 1 (0%) |
| - Oesophageal ulcer haemorrhage | 1 (0%) | 0 (0%) |
| **General disorders and administration site conditions** | 5 (1%) | 10 (2%) |
| - Chest pain | 1 (0%) | 0 (0%) |
| - General physical health deterioration | 0 (0%) | 1 (0%) |
| - Malaise | 0 (0%) | 1 (0%) |
| - Multiple organ dysfunction syndrom | 4 (1%) | 7 (2%) |
| - Oedema | 0 (0%) | 1 (0%) |
| **Hepatobiliary disorders** | 1 (0%) | 3 (1%) |
| - Cholangitis | 0 (0%) | 1 (0%) |
| - Hepatocellular injury | 0 (0%) | 1 (0%) |
| - Hepatorenal syndrome | 1 (0%) | 1 (0%) |
| **Infections and infestations** | 14 (3%) | 18 (4%) |
| - Aspergillus infection | 0 (0%) | 0 (0%) |
| - Bacterial infection | 2 (0%) | 1 (0%) |
| - Bronchitis | 1 (0%) | 0 (0%) |
| - Dengue fever | 1 (0%) | 0 (0%) |
| - Hepatitis viral | 0 (0%) | 1 (0%) |
| - Infection | 1 (0%) | 0 (0%) |
| - Pneumonia | 7 (2%) | 13 (3%) |
| - Sepsis | 2 (0%) | 2 (0%) |
| - Superinfection fungal | 0(0%) | 0 (0%) |
| - Urinary tract infection | 0 (0%) | 1 (0%) |
| **Injury, poisoning and procedural complications** | 16 (4%) | 2 (0%) |

| no (%) | Remdesivir (n=410) | Control (n=423) |
|---|---|---|
| - Mechanical ventilation complication | 1 (0%) | 0 (0%) |
| - Overdose | 1 (0%) | 1 (0%) |
| - Product administration error | 14 (3%) | 0 (0%) |
| - Sedation complication | 0 (0%) | 1 (0%) |
| **Investigations** | 16 (4%) | 3 (1%) |
| - Aspartate aminotransferase increase | 1 (0%) | 0 (0%) |
| - Blood creatine phosphokinase increase | 2 (0%) | 0 (0%) |
| - Blood glucose increased | 1 (0%) | 0 (0%) |
| - Gamma-glutamyltransferase increase | 1 (0%) | 0 (0%) |
| - Glomerular filtration rate decrease | 1 (0%) | 0 (0%) |
| - Haemoglobin decreased | 0 (0%) | 1 (0%) |
| - Lipase increased | 0 (0%) | 1 (0%) |
| - Oxygen consumption increased | 0 (0%) | 1 (0%) |
| - Oxygen saturation decreased | 1 (0%) | 0 (0%) |
| - Serum creatinine increased | 0 (0%) | 0 (0%) |
| - Transaminases increased | 9 (2%) | 0 (0%) |
| **Metabolism and nutrition disorders** | 9 (2%) | 11 (3%) |
| - Alkalosis | 1 (0%) | 2 (0%) |
| - Decreased appetite | 0 (0%) | 1 (0%) |
| - Dehydration | 1 (0%) | 0 (0%) |
| - Hyperglycaemia | 5 (1%) | 4 (1%) |
| - Hyperkalaemia | 0 (0%) | 1 (0%) |
| - Hypernatremia | 1 (0%) | 0 (0%) |
| - Hypalbuminaemia | 1 (0%) | 1 (0%) |
| - Hyponatraemia | 0 (0%) | 1 (0%) |
| - Malnutrition | 0 (0%) | 1 (0%) |
| **Neoplasms benign, malignant and unspecified** | 3 (1%) | 2 (0%) |
| - Hepatic cancer | 0 (0%) | 1 (0%) |
| - Hepatocellular carcinoma | 1 (0%) | 0 (0%) |
| - Lung cancer metastatic | 1 (0%) | 0 (0%) |
| - Lung neoplasm malignant | 0 (0%) | 1 (0%) |
| - Malignant neoplasm progression | 1 (0%) | 0 (0%) |
| **Nervous system disorders** | 7 (2%) | 1 (0%) |
| - Acute haemorrhagic leukoencephalitis | 1 (0%) | 0 (0%) |
| - Cerebrovascular accident | 2 (0%) | 0 (0%) |
| - Encephalopathy | 2 (0%) | 0 (0%) |
| - Extrapyramidal disorder | 1 (0%) | 0 (0%) |
| - Hepatic encephalopathy | 1 (0%) | 0 (0%) |
| - Neuropathy peripheral | 0 (0%) | 0 (0%) |
| - Somnolence | 0 (0%) | 1 (0%) |
| **Renal and urinary disorders** | 20 (5%) | 26 (6%) |
| - Acute kidney injury | 16 (4%) | 21 (5%) |
| - Hepatorenal syndrome | 1 (0%) | 1 (0%) |
| - Renal failure | 2 (0%) | 3 (1%) |
| - Renal tubular disorder | 1 (0%) | 0 (0%) |
| - Urinary retention | 0 (0%) | 1 (0%) |
| **Respiratory, thoracic and mediastinal disorders** | 76 (18%) | 83 (20%) |
| - Acute respiratory distress syndrome | 40 (10%) | 41 (10%) |
| - Acute respiratory failure | 5 (1%) | 11 (3%) |
| - Dyspnoea | 4 (1%) | 3 (1%) |
| - Dyspnoea exertional | 1 (0%) | 0 (0%) |
| - Hypoxia | 5 (1%) | 4 (1%) |
| - Lung disorder | 0 (0%) | 1 (0%) |
| - Pleural effusion | 6 (1%) | 4 (1%) |
| - Pneumomediastinum | 0 (0%) | 1 (0%) |

| no (%) | Remdesivir (n=410) | Control (n=423) |
|---|---|---|
| - Pneumothorax | 4 (0%) | 4 (1%) |
| - Pulmonary embolism | 10 (2%) | 12 (3%) |
| - Pulmonary oedema | 0 (0%) | 1 (0%) |
| - Respiratory acidosis | 1 (0%) | 0 (0%) |
| - Respiratory alkalosis | 0 (0%) | 1 (0%) |
| **Skin and subcutaneous tissue disorders** | 0 (0%) | 2 (0%) |
| - Rash | 0 (0%) | 1 (0%) |
| - Subcutaneous emphysema | 0 (0%) | 1 (0%) |
| **Vascular disorders** | 8 (2%) | 5 (1%) |
| - Device related thrombosis | 2 (0%) | 0 (0%) |
| - Haemodynamic instability | 1 (0%) | 0 (0%) |
| - Hypertension | 0 (0%) | 1 (0%) |
| - Hypotension | 2 (0%) | 0 (0%) |
| - Orthostatic hypotension | 1 (0%) | 0 (0%) |
| - Shock | 1 (0%) | 2 (0%) |
| - Thrombosis | 1 (0%) | 2 (0%) |

**Supplementary Figure S1. Clinical status, as measured by the 7-point ordinal scale, at day 15 and day 29 in the intention-to-treat population of the DisCoVeRy trial, in severe patients stratified by absence (top) or presence (bottom) of invasive mechanical ventilation or ECMO at randomization.**

Reported numbers refer to the proportion of patients with the corresponding level in each group.

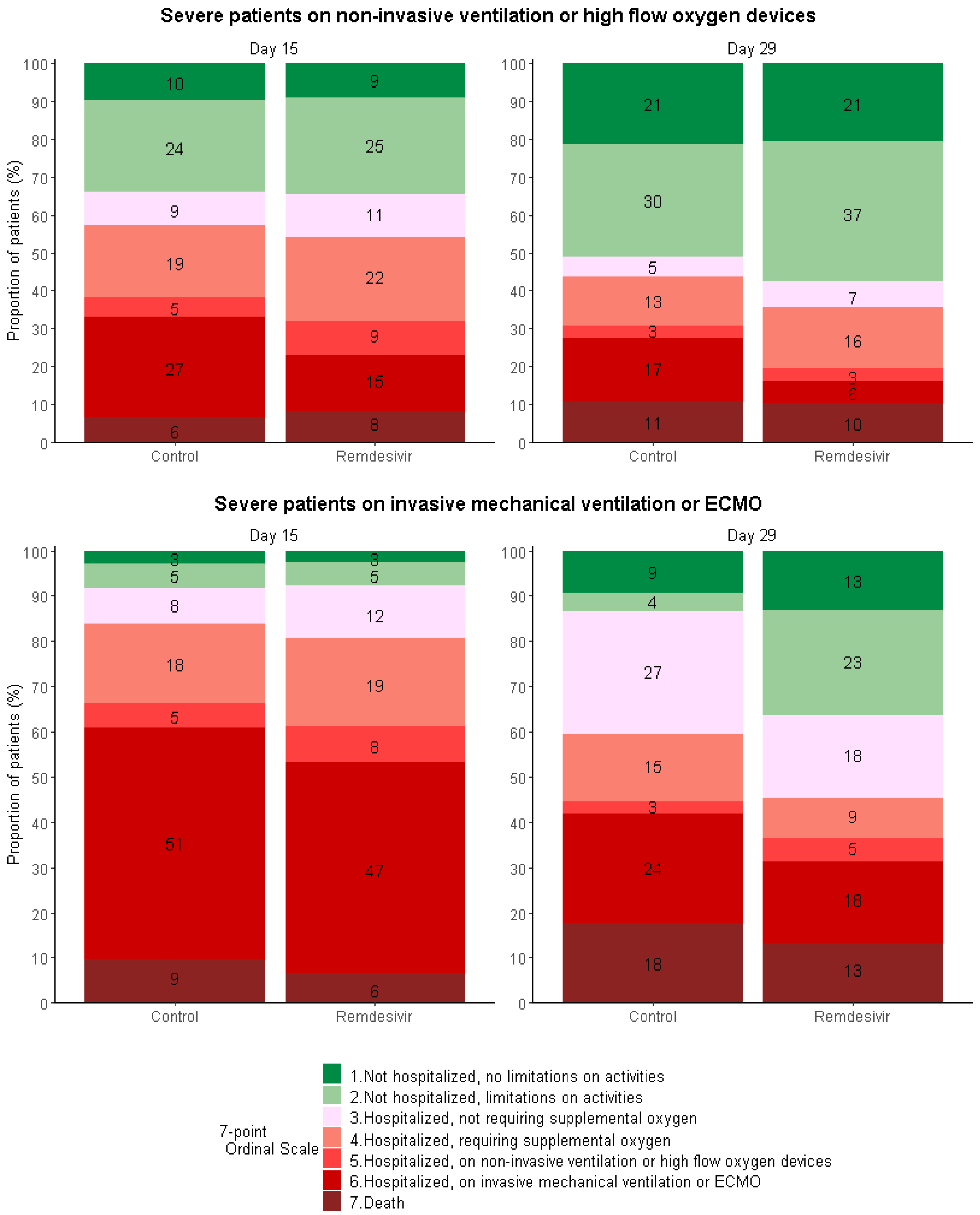

**Supplementary Figure S2. Forrest plot of subgroup analyses of the primary outcome in the intention-to-treat population of the DisCoVeRy trial.**
The widths of the confidence intervals have not been adjusted for multiplicity and therefore cannot be used to infer treatment effects.

| Subgroups | Number of patients | OR (95%CI) | P-value |
|---|---|---|---|
| **Age** | | | |
| < 50 | 135 | 0.99 (0.78 to 1.26) | 0.38 |
| [50;70[ | 422 | 0.90 (0.64 to 1.27) | 0.56 |
| ≥ 70 | 286 | 0.98 (0.65 to 1.47) | 0.92 |
| **Sex** | | | |
| Men | 586 | 0.92 (0.69 to 1.22) | 0.55 |
| Women | 257 | 1.20 (0.78 to 1.86) | 0.41 |
| **Duration of symptoms prior to randomization** | | | |
| ≤ 7 days | 269 | 1.12 (0.74 to 1.72) | 0.57 |
| 8-14 days | 488 | 1.01 (0.73 to 1.38) | 0.95 |
| >14 days | 69 | 0.46 (0.19 to 1.10) | 0.08 |
| **Severity at randomization** | | | |
| Moderate | 511 | 0.85 (0.62 to 1.17) | 0.33 |
| Severe | 332 | 1.18 (0.80 to 1.73) | 0.40 |
| **Country** | | | |
| France | 715 | 1.06 (0.82 to 1.37) | 0.67 |
| Austria | 29 | 0.91 (0.24 to 3.42) | 0.89 |
| Portugal | 34 | 0.90 (0.26 to 3.09) | 0.87 |
| Belgium/Luxembourg | 65 | 0.47 (0.19 to 1.14) | 0.10 |
| **All patients** | **843** | **0.93 (0.78 to 1.26)** | **0.93** |

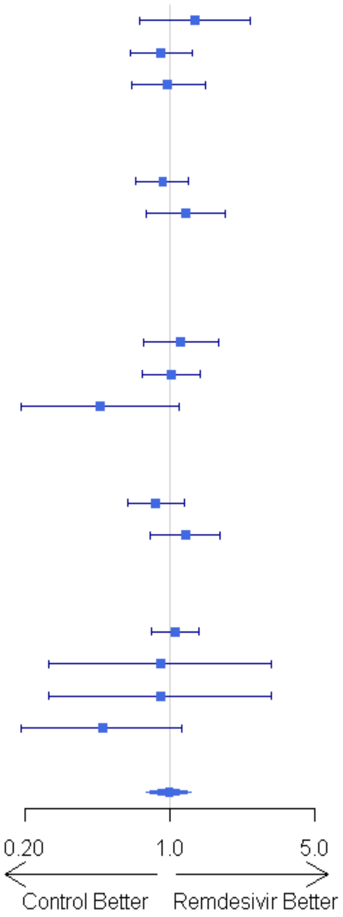

**Supplementary Figure S3. Time to new mechanical ventilation, new ECMO or death between baseline and day 29 in the intention-to-treat population of the DisCoVeRy trial, according to disease severity at randomization in moderate and severe disease without ventilation or ECMO at randomization (panel A), in participants with moderate disease at randomization (panel B) and in participants with severe disease without ventilation or ECMO at randomization (panel C).**

In the Panel A, analyses were stratified on the disease severity at randomization and reported hazard ratio is adjusted on disease severity at randomization.

In the panels B and C, we performed non-prespecified subgroup analyses according to the disease severity at baseline, and reported hazard ratios are specific to each disease severity subgroup.

Remdesivir (green line); control (black line). HR, Hazard ratio; 95%CI, 95% confidence interval.

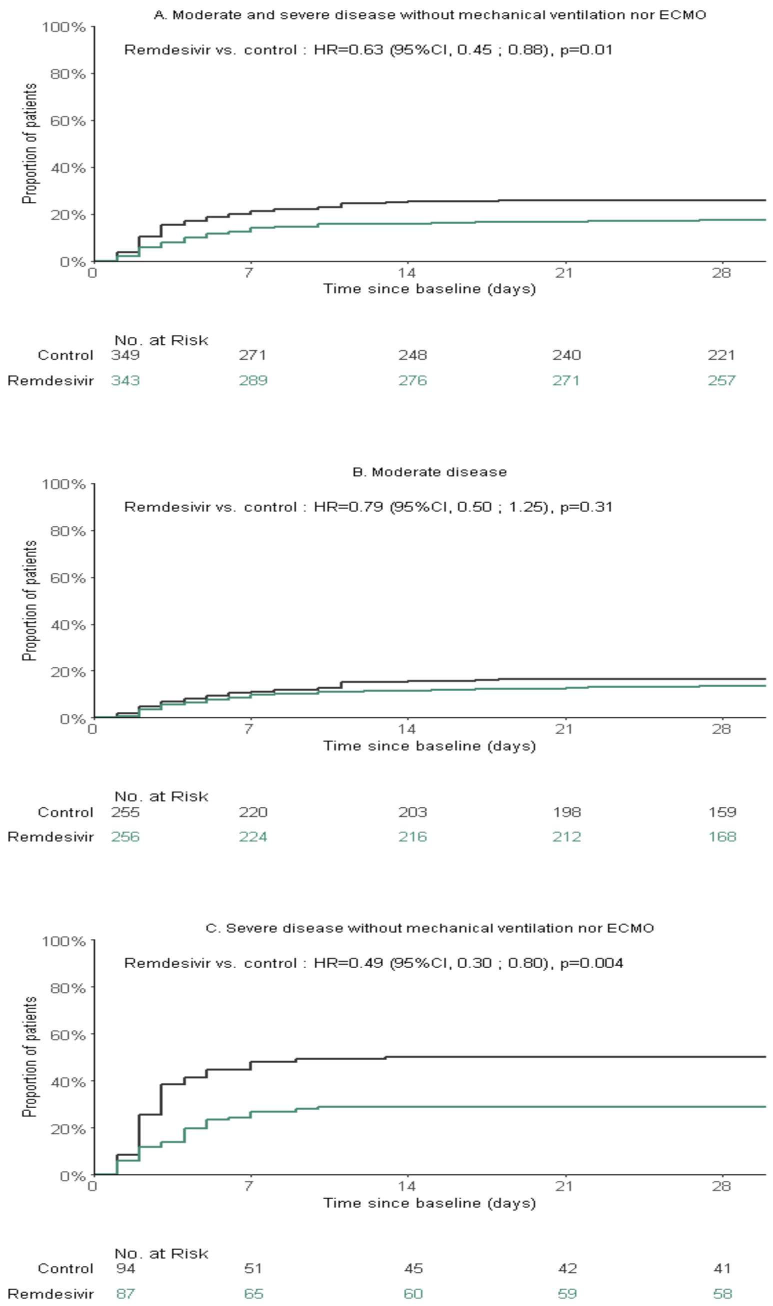

**Supplementary Figure S4. Time to improvement of at least 2 categories or the 7-point ordinal scale or hospital discharge between baseline and day 29 in the intention-to-treat population of the DisCoVeRy trial, according to disease severity at randomization in all participants (panel A), in participants with moderate disease at randomization (panel B) and in participants with severe disease at randomization (panel C).**
Analyses were stratified on the disease severity at randomization and reported aazard ratio is adjusted on disease severity at randomization. Remdesivir (green line); control (black line). HR, Hazard ratio; 95%CI, 95% confidence interval.

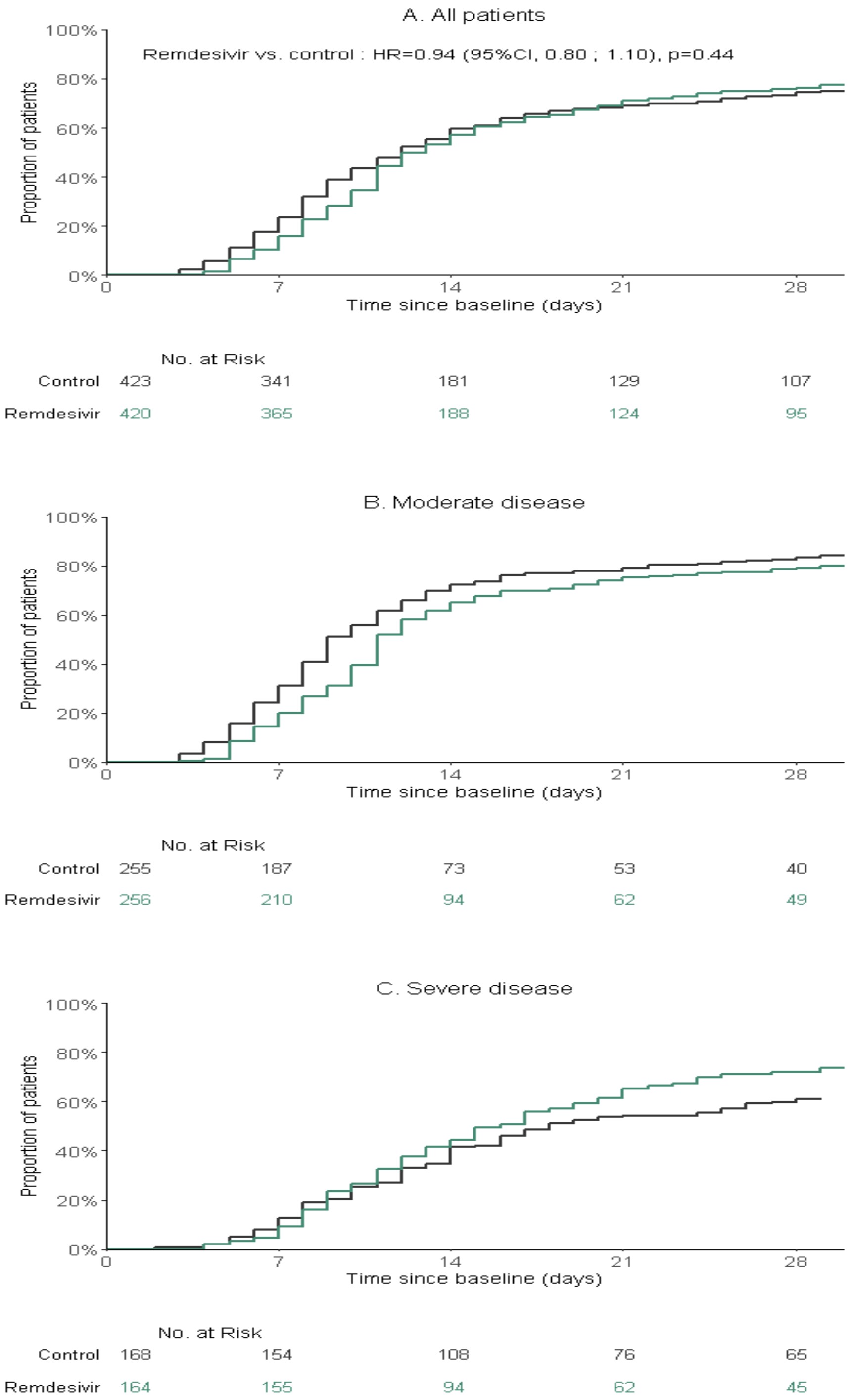

**Supplementary Figure S5. Time to improvement of at least 2 categories or the 7-point ordinal scale or hospital discharge between baseline and day 29 in the intention-to-treat population of the DisCoVeRy trial, in severe patients stratified by absence (left) or presence (right) of invasive mechanical ventilation or ECMO at randomisation.**
Remdesivir (green line); control (black line).

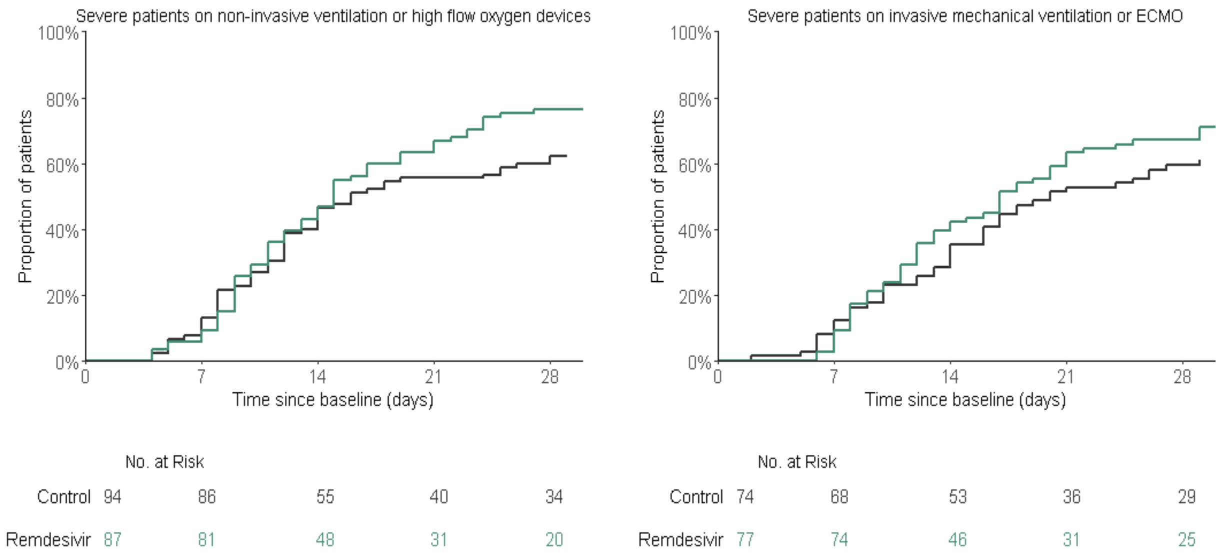

**Supplementary Figure S6. Time to National Early Warning Score ≤2 or hospital discharge between baseline and day 29 in the intention-to-treat population of the DisCoVeRy trial, according to disease severity at randomization in all participants (panel A), in participants with moderate disease at randomization (panel B) and in participants with severe disease at randomization (panel C).**

Remdesivir (green line); control (black line). HR, Hazard ratio; 95%CI, 95% confidence interval.

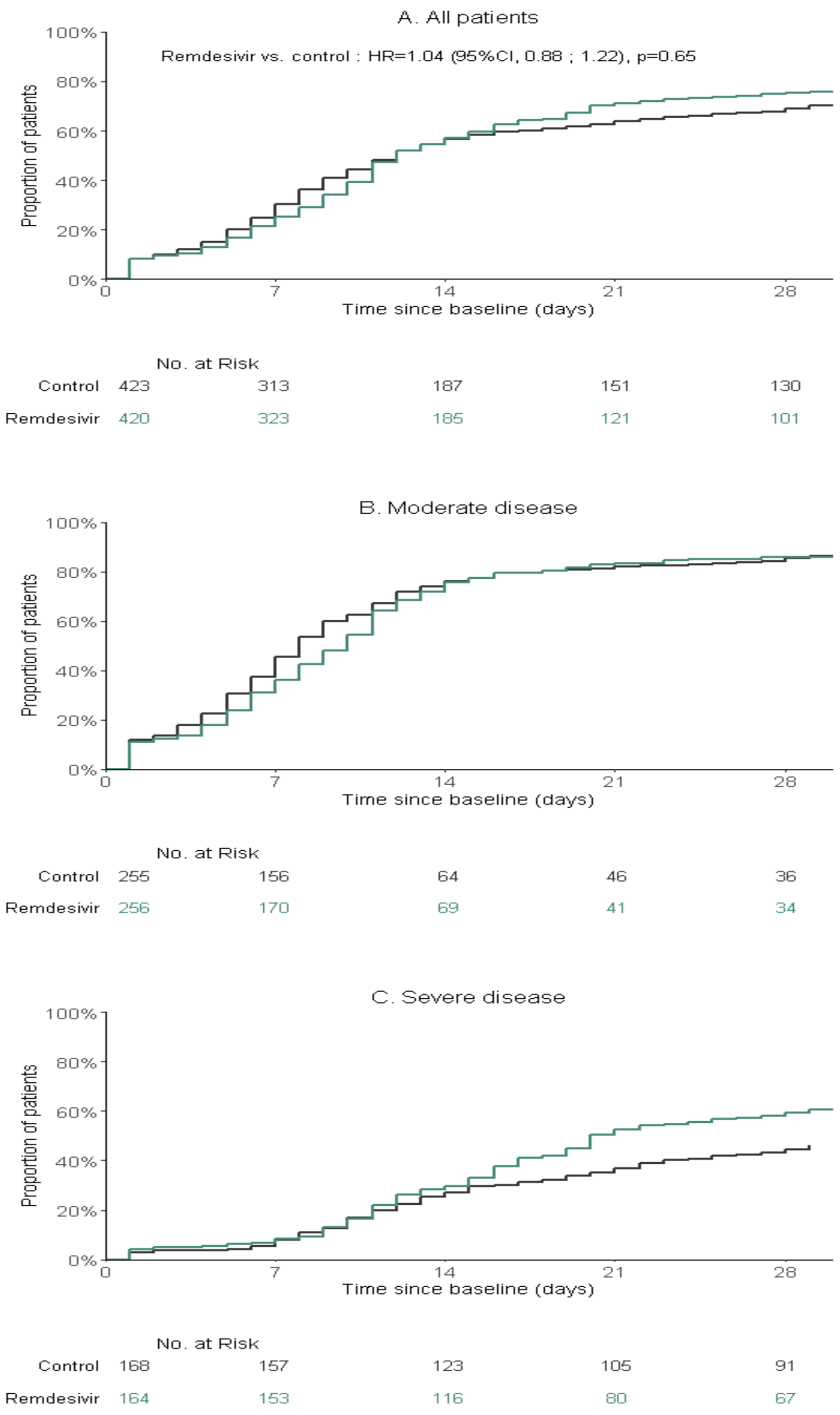

**Supplementary Figure S7. Time to National Early Warning Score ≤2 or hospital discharge between baseline and day 29 in the intention-to-treat population of the DisCoVeRy trial, in severe patients stratified by absence (left) or presence (right) of invasive mechanical ventilation or ECMO at randomisation.**
Remdesivir (green line); control (black line).

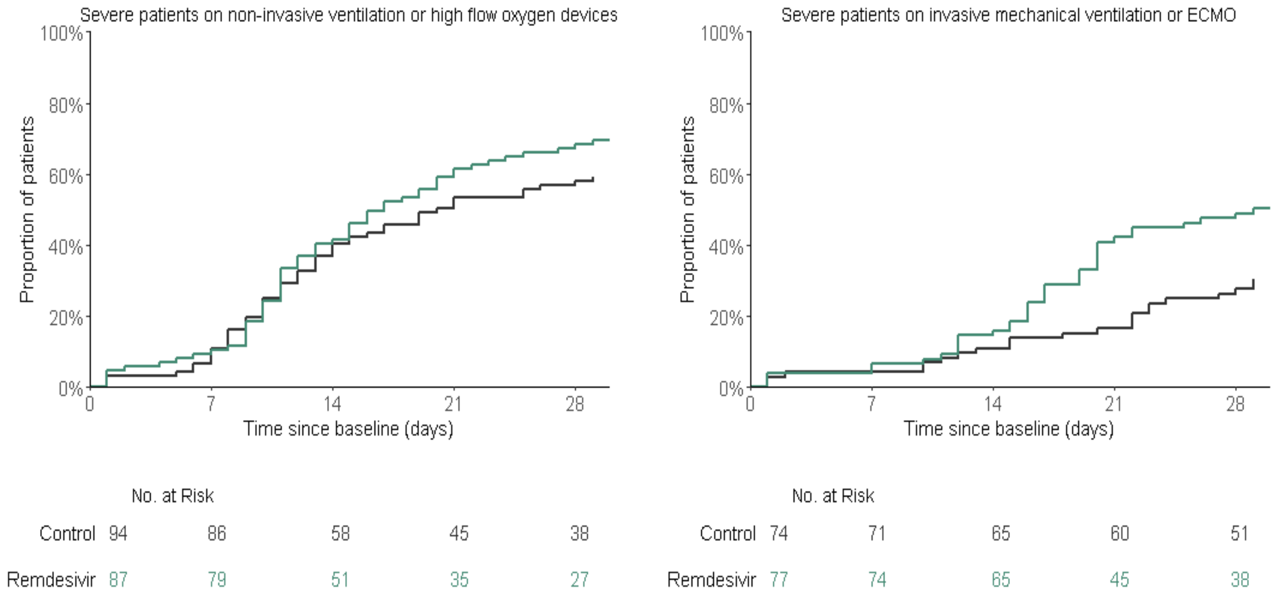

**Supplementary Figure S8. Time to hospital discharge between baseline and day 29 in the intention-to-treat population of the DisCoVeRy trial, according to disease severity at randomization in all participants (panel A), in participants with moderate disease at randomization (panel B) and in participants with severe disease at randomization (panel C).** Remdesivir (green line); control (black line). HR, Hazard ratio; 95%CI, 95% confidence interval.

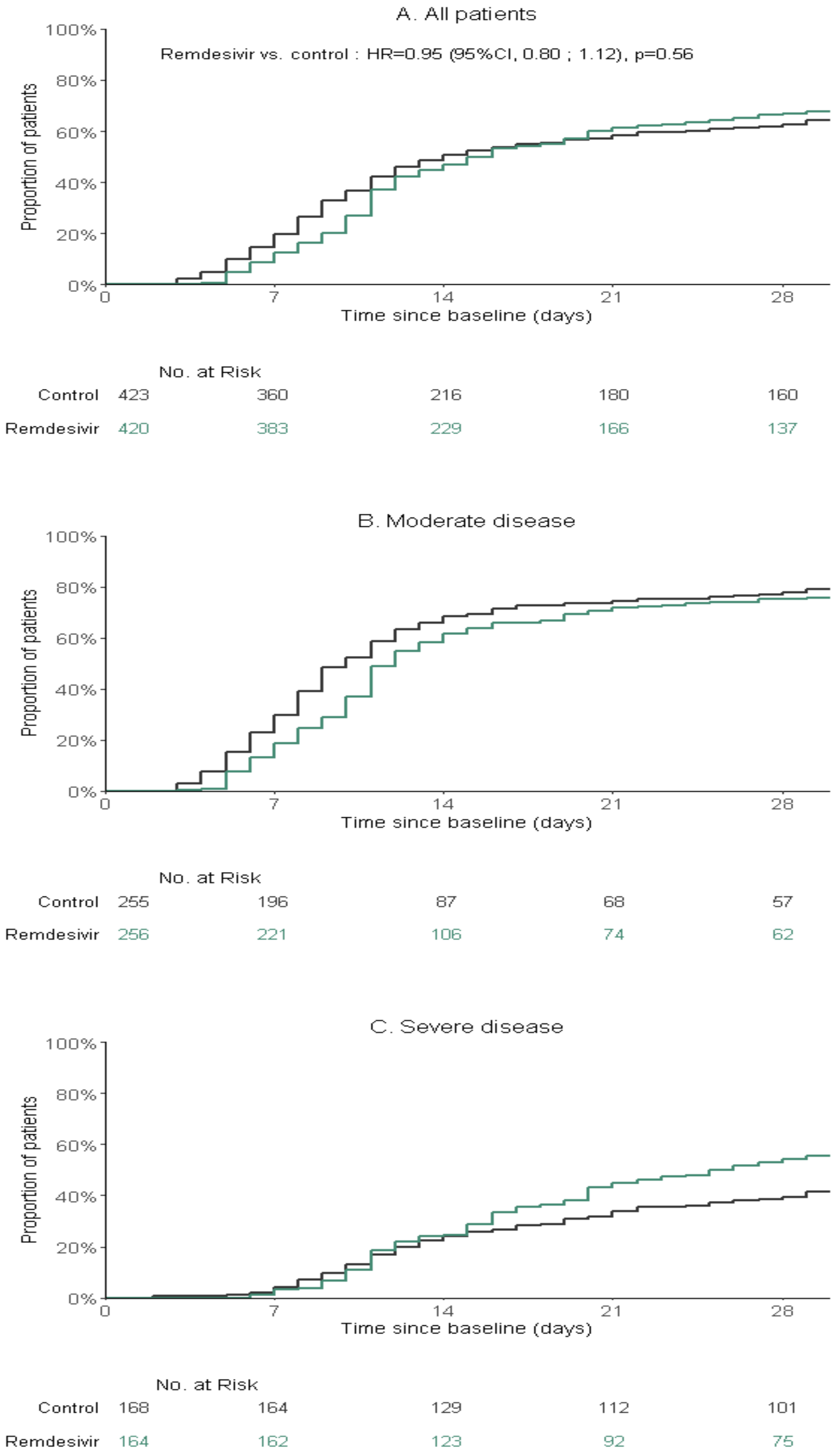

**Supplementary Figure S9. Time to hospital discharge between baseline and day 29 in the intention-to-treat population of the DisCoVeRy trial, in severe patients stratified by absence (left) or presence (right) of invasive mechanical ventilation or ECMO at randomisation.** Remdesivir (green line); control (black line).

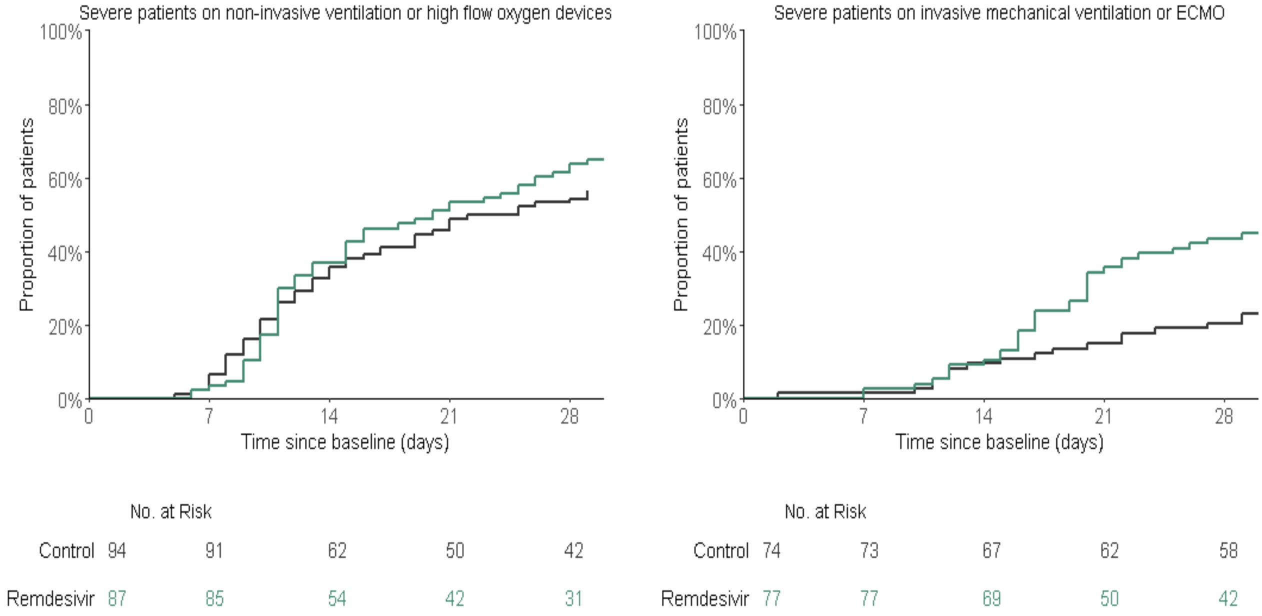

**Supplementary Figure S10. Evolution of the normalized SARS-CoV-2 viral load in nasopharyngeal swabs between baseline and day 15 in the intention-to-treat population of the DisCoVeRy trial, in severe patients stratified by absence (left) or presence (right) of invasive mechanical ventilation or ECMO at randomisation: $\log_{10}$ viral loads (left), change from baseline of the $\log_{10}$ viral loads (right).**

Data are presented as means (95%CI). Remdesivir (green line); control (black line).

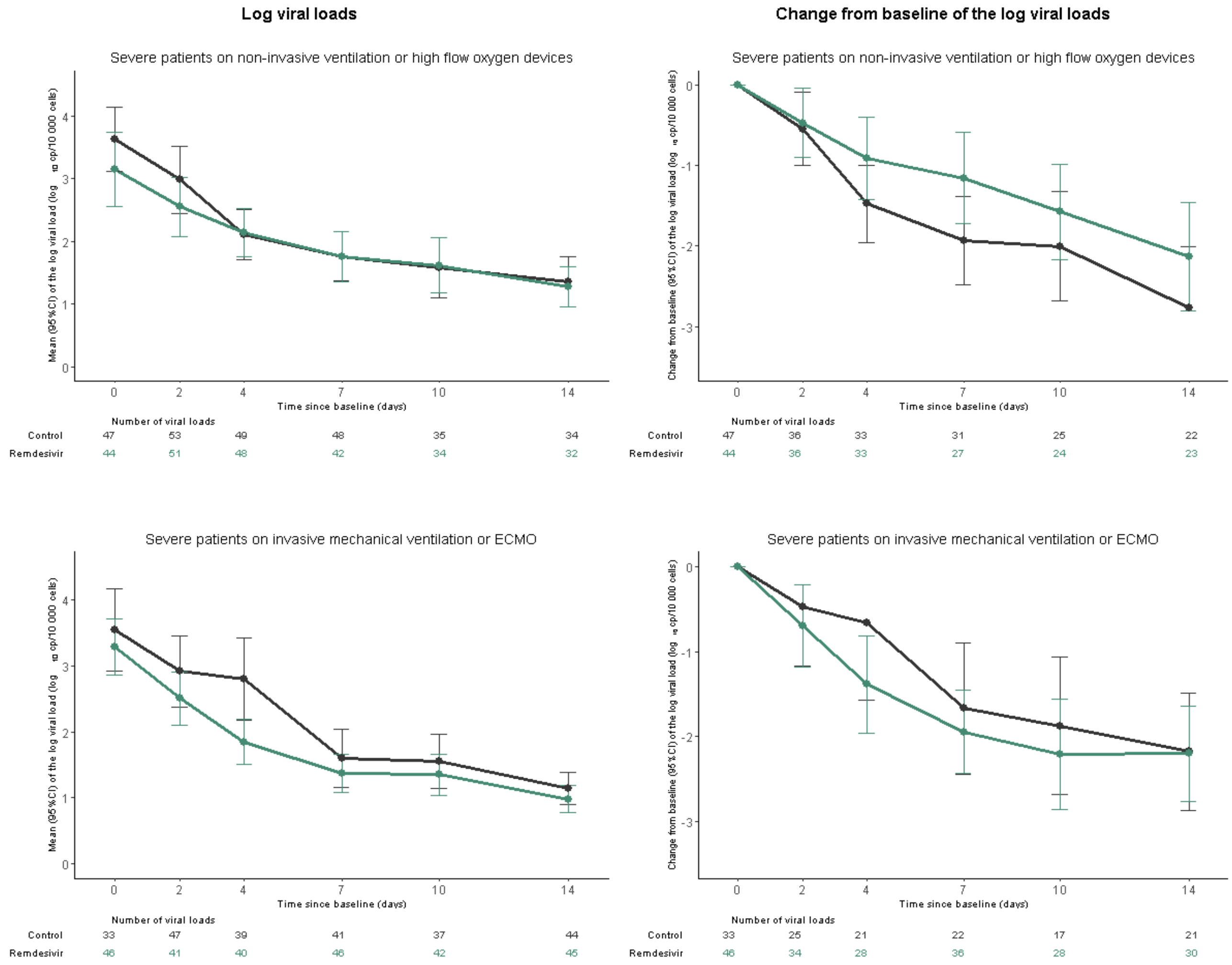

### CONSORT 2010 checklist of information to include when reporting a randomised trial*

| Section/Topic | Item No | Checklist item | Reported on page No |
|---|---|---|---|
| **Title and abstract** | | | |
| | 1a | Identification as a randomised trial in the title | 1 |
| | 1b | Structured summary of trial design, methods, results, and conclusions (for specific guidance see CONSORT for abstracts) | 9 |
| **Introduction** | | | |
| Background and objectives | 2a | Scientific background and explanation of rationale | 11 |
| | 2b | Specific objectives or hypotheses | 12 |
| **Methods** | | | |
| Trial design | 3a | Description of trial design (such as parallel, factorial) including allocation ratio | 12 |
| | 3b | Important changes to methods after trial commencement (such as eligibility criteria), with reasons | Suppl appendix |
| Participants | 4a | Eligibility criteria for participants | 12-13 |
| | 4b | Settings and locations where the data were collected | 12 |
| Interventions | 5 | The interventions for each group with sufficient details to allow replication, including how and when they were actually administered | 14 |
| Outcomes | 6a | Completely defined pre-specified primary and secondary outcome measures, including how and when they were assessed | 14-15 |
| | 6b | Any changes to trial outcomes after the trial commenced, with reasons | NA |
| Sample size | 7a | How sample size was determined | 16-17 |
| | 7b | When applicable, explanation of any interim analyses and stopping guidelines | 17 |
| Randomisation: | | | |
| Sequence generation | 8a | Method used to generate the random allocation sequence | 13 |
| | 8b | Type of randomisation; details of any restriction (such as blocking and block size) | 13 |
| Allocation concealment mechanism | 9 | Mechanism used to implement the random allocation sequence (such as sequentially numbered containers), describing any steps taken to conceal the sequence until interventions were assigned | 13 |
| Implementation | 10 | Who generated the random allocation sequence, who enrolled participants, and who assigned participants to interventions | 13 |
| Blinding | 11a | If done, who was blinded after assignment to interventions (for example, participants, care providers, those assessing outcomes) and how | 13 |
| | 11b | If relevant, description of the similarity of interventions | NA |

| | | | |
|---|---|---|---|
| Statistical methods | 12a | Statistical methods used to compare groups for primary and secondary outcomes | 18-19 |
| | 12b | Methods for additional analyses, such as subgroup analyses and adjusted analyses | 18-19 |
| **Results** | | | |
| Participant flow (a diagram is strongly recommended) | 13a | For each group, the numbers of participants who were randomly assigned, received intended treatment, and were analysed for the primary outcome | 19-20 |
| | 13b | For each group, losses and exclusions after randomisation, together with reasons | Figure 1 |
| Recruitment | 14a | Dates defining the periods of recruitment and follow-up | 19 |
| | 14b | Why the trial ended or was stopped | 17 |
| Baseline data | 15 | A table showing baseline demographic and clinical characteristics for each group | Table 1 |
| Numbers analysed | 16 | For each group, number of participants (denominator) included in each analysis and whether the analysis was by original assigned groups | 19-20 |
| Outcomes and estimation | 17a | For each primary and secondary outcome, results for each group, and the estimated effect size and its precision (such as 95% confidence interval) | 29-22, Table 2, Suppl Appendix |
| | 17b | For binary outcomes, presentation of both absolute and relative effect sizes is recommended | 29-22, Table 2, Suppl Appendix |
| Ancillary analyses | 18 | Results of any other analyses performed, including subgroup analyses and adjusted analyses, distinguishing pre-specified from exploratory | 29-22, Table 2, Suppl Appendix |
| Harms | 19 | All important harms or unintended effects in each group (for specific guidance see CONSORT for harms) | Table 3 |
| **Discussion** | | | |
| Limitations | 20 | Trial limitations, addressing sources of potential bias, imprecision, and, if relevant, multiplicity of analyses | 24-25 |
| Generalisability | 21 | Generalisability (external validity, applicability) of the trial findings | 24-25 |
| Interpretation | 22 | Interpretation consistent with results, balancing benefits and harms, and considering other relevant evidence | 22-25 |
| **Other information** | | | |
| Registration | 23 | Registration number and name of trial registry | 19 |
| Protocol | 24 | Where the full trial protocol can be accessed, if available | Suppl Appendix |
| Funding | 25 | Sources of funding and other support (such as supply of drugs), role of funders | 19, 28-29 |

*We strongly recommend reading this statement in conjunction with the CONSORT 2010 Explanation and Elaboration for important clarifications on all the items. If relevant, we also recommend reading CONSORT extensions for cluster randomised trials, non-inferiority and equivalence trials, non-pharmacological treatments, herbal interventions, and pragmatic trials. Additional extensions are forthcoming: for those and for up to date references relevant to this checklist, see www.consort-statement.org.

### DisCoVeRy Study Group

**Sponsor:** Sandrine Couffin-Cadiègues, Hélène Esperou (Pôle de Recherche Clinique, Inserm, Paris, France)

**Investigators** (countries in alphabetical order):

***Austria**:* Bernd Lamprecht (Kepler Universitätsklinikum Linz, Linz), Michael Joannidis (Medizinische Universität Innsbruck, Innsbruck), Alexander Egle, Richard Greil (Paracelsus Medical University Salzburg, SCRI-CCCI and AGMT)

***Belgium**:* Antoine Altdorfer, Vincent Fraipont (Centre Hospitalier Régional de la Citadelle, Liège), Leila Belkhir (Cliniques Universitaires de Saint Luc, Bruxelles), Maya Hites, Gil Verschelden (Hôpital Erasme, Cliniques Universitaires de Bruxelles)

***France***: Violaine Tolsma, David Bougon (Centre Hospitalier Annecy-Genevois) ; Agathe Delbove, Marie Gousseff (Centre Hospitalier Bretagne-Atlantique, Vannes) ; Nadia Saidani, Guilhem Wattecamps (Centre Hospitalier Cornouaille, Quimper) ; Félix Djossou, Loïc Epelboin (Centre Hospitalier de Cayenne Andrée Rosemon) ; Jean-Philippe Lanoix, Pierre-Alexandre Roger, Claire Andrejak, Yoann Zerbib (Centre Hospitalier Universitaire de Amiens) ; Kevin Bouiller, Catherine Chirouze, Jean-Christophe Navellou (Centre Hospitalier Universitaire de Besançon) ; Alexandre Boyer, Charles Cazanave, Alexandre Duvignaud, Didier Gruson, Denis Malvy (Centre Hospitalier Universitaire de Bordeaux) ; Henry Lessire, Martin Martinot (Hospices Civils de Colmar) ; Pascal Andreu, Mathieu Blot, Lionel Piroth, Jean Pierre Quenot (Centre Hospitalier Universitaire de Dijon) ; Olivier Epaulard, Nicolas Terzi (Centre Hospitalier Universitaire de Grenoble-Alpes) ; Karine Faure, Emmanuel Faure, Julien Poissy, Saad Nseir (Centre Hospitalier Régional Universitaire de Lille) ; Florence Ader, Laurent Argaud, Tristan Ferry, Thomas Perpoint, Vincent Piriou, Jean-Christophe Richard, Julien Textoris, Florent Valour, Florent Wallet (Hospices Civils de Lyon) ; André Cabié, Jean-Marie Turmel, Cyrille Chabartier (Centre Hospitalier Universitaire de Martinique, Fort-de-France) ; Rostane Gaci, Céline Robert (Centre Hospitalier Régional de Metz-Thionville) ; Alain Makinson, Vincent Le Moing, Kada Klouche (Centre Hospitalier Universitaire de Montpellier) ; Olivier Hinschberger, Joy Mootien (Centre Hospitalier de Mulhouse Sud-Alsace) ; Sébastien Gibot, François Goehringer, Antoine Kimmoun, Benjamin Lefevre (Centre Hospitalier Régional Universitaire de Nancy) ; David Boutoille, Emmanuel Canet, Benjamin Gaborit, Paul Le Turnier, François Raffi, Jean Reignier (Centre Hospitalier Universitaire de Nantes) ; Johan Courjon, Jean Dellamonica, Sylvie Leroy, Charles-Hugo Marquette (Centre Hospitalier Universitaire de Nice), Paul Loubet, Claire Roger, Albert Sotto (Centre Hospitalier Universitaire de Nîmes) ; Cédric Bruel, Benoît Pilmis (Groupe Hospitalier de Paris Saint-Joseph) ; Guillaume Geri, Elisabeth Rouveix-Nordon (Hôpital Ambroise Paré, Assistance Publique – Hôpitaux de Paris); Olivier Bouchaud (Hôpital Avicenne, Assistance Publique – Hôpitaux de Paris); Samy Figueiredo, Stéphane Jaureguiberry, Xavier Monnet (Hôpital Bicêtre, Assistance Publique – Hôpitaux de Paris); Lila Bouadma, François-

Xavier Lescure, Nathan Peiffer-Smadja, Jean-François Timsit, Yazdan Yazdanpanah (Hôpital Bichat - Claude Bernard, Assistance Publique – Hôpitaux de Paris); Solen Kerneis, Marie Lachâtre, Odile Launay, Jean-Paul Mira (Hôpital Cochin, Assistance Publique – Hôpitaux de Paris) ; Julien Mayaux, Valérie Pourcher (Hôpital de la Pitié-Salpêtrière, Assistance Publique – Hôpitaux de Paris); Jérôme Aboab, Flora Crockett, Naomi Sayre (Hôpital Delafontaine, Saint-Denis), Clément Dubost, Cécile Ficko (Hôpital d'Instruction des Armées Bégin, Saint Mandé) ; David Lebeaux (Hôpital Européen Georges-Pompidou, Assistance Publique – Hôpitaux de Paris) ; Sébastien Gallien, Armand Mekontso-Dessap (Hôpital Henri-Mondor, Assistance Publique – Hôpitaux de Paris) ; Jérôme Le Pavec, Francois Stefan (Hôpital Marie Lannelongue, Le Plessis-Robinson); Hafid Ait-Oufella, Karine Lacombe (Hôpital Saint-Antoine, Assistance Publique – Hôpitaux de Paris); Jean-Michel Molina (Hôpital Saint-Louis, Assistance Publique – Hôpitaux de Paris); Murielle Fartoukh, Gilles Pialoux (Hôpital Tenon, Assistance Publique – Hôpitaux de Paris) ; Firouzé Bani-Sadr, Bruno Mourvillier (Centre Hospitalier Universitaire de Reims) ; François Benezit, Fabrice Laine, Bruno Laviolle, Yves Le Tulzo, Matthieu Revest (Centre Hospitalier Universitaire de Rennes) ; Elisabeth Botelho-Nevers, Amandine Gagneux-Brunon, Guillaume Thiery (Centre Hospitalier Universitaire de Saint-Étienne) ; François Danion, Yves Hansmann, Ferhat Meziani, Walid Oulehri, Charles Tacquard (Centre Hospitalier Universitaire de Strasbourg) ; Fanny Bounes-Vardon, Guillaume Martin-Blondel, Marlène Murris-Espin, Béatrice Riu-Poulenc (Centre Hospitalier Universitaire de Toulouse) ; Vanessa Jeanmichel, Eric Senneville (Centre Hospitalier de Tourcoing) ; Louis Bernard, Denis Garot (Centre Hospitalier Universitaire de Tours).

***Luxembourg***: Jean Reuter, Thérèse Staub (Centre Hospitalier de Luxembourg) ; Marc Berna (Hôpitaux Robert Schuman).

***Portugal****:* Sandra Braz, Joao Miguel Ferreira Ribeiro (Centro Hospital Universitário de Lisboa Norte, Hospital de Santa Maria); José-Artur Paiva, Roberto Roncon-Albuquerque (Centro Hospitalar Universitário São João de Porto).

**Laboratory support** : Benjamin Leveau (CRB, Hospices Civils de Lyon)

### Acknowledgements

We acknowledge all the patients enrolled in the study and all the health-care workers involved in patients' management.

We acknowledge the members of the Data Safety Monitoring Board:
Stefano Vella (chair), Karen Barnes, Phaik Yeong Cheah, Guanhua Du, Mike Jacobs, Donata Medaglini, Stuart Pocock, Sylvie Van Der Werf, Patrick Yéni, and the independent statistician Tim Collier.

We acknowledge the DisCoVeRy Steering Committee (by alphabetical order):
Voting members: Florence Ader, Dominique Costagliola, Alpha Diallo, Alexander Egle, Hélène Espérou, Monika Halanova, Maya Hites, Bruno Lina, France Mentré, José-Artur Paiva, Marie-Thérèse Staub, Yazdan Yazdanpanah.
Non-voting members: Pascale Augé, Drifa Belhadi, Diana Brainard, Charles Burdet, Mireille Caralp, Sandrine Couffin-Cadiergues, Aline Dechanet, Marta Del Alamo, Christelle Delmas, Jacques Demotes, Marina Dumousseaux, Monica Ensini, Joe Eustace, Richard Greil, Christophe Hezode, Fionnuala Keane, Marie-Paule Kiény, Soizic Le Mestre, Bernd Muehlbauer, Mickael Ohana, Nathan Peiffer Smadja, Ventzislava Petrov-Sanchez, Gilles Peytavin, Julien Poissy, Isabel Püntmann, Cécile Rabian, Jean Reuter, Juliette Saillard, Benjamin Terrier, Vida Terzic.

We acknowledge the French DisCoVeRy Trial Management Team (by alphabetical order):
Laurent Abel, Florence Ader, Toni Alfaiate, Claire Andrejak, Basma Basli, Drifa Belhadi, Lila Bouadma, Maude Bouscambert, Charles Burdet, Carole Cagnot, Anissa Chair, Dominique Costagliola, Sandrine Couffin-Cadiergues, Eric D'Ortenzio, Aline Dechanet, Christelle Delmas, Alpha Diallo, Axelle Dupont, Hélène Esperou, Assia Ferrane, Claire Fougerou, Alexandre Gaymard, Ambre Gelley, Jérémie Guedj, Benjamin Hamze, Vinca Icard, Samira Laribi, Minh-Patrick Lê, Soizic Le Mestre, Delphine Lebrasseur- Longuet, François-Xavier Lescure, Benjamin Leveau, Bruno Lina, France Mentré, Noémie Mercier, Laëtitia Moinot, Florence Morfin-Sherpa, Marion Noret, Nathan Peiffer-Smadja, Ventzislava Petrov-Sanchez, Gilles Peytavin, Julien Poissy, Oriane Puéchal, Juliette Saillard, Marion Schneider, Caroline Semaille, Marie-Capucine Tellier, Jean-François Timsit, Sarah Tubiana, Priyanka Velou, Linda Wittkop, Yazdan Yazdanpanah.

We acknowledge the Working Groups of the DisCoVeRy Trial (by alphabetical order):
Clinical aspects: Florence Ader, Claire Andrejak, Charles Burdet, Christelle Delmas, Marina Dumousseaux, François-Xavier Lescure, Nathan Peiffer-Smadja, Julien Poissy, Juliette Saillard.

Study products: Aline Dechanet, Christelle Delmas, Marina Dumousseaux, Assia Ferrane, Benjamin Hamze, Juliette Saillard.

Methodology and Statistics: Drifa Belhadi, Charles Burdet, Dominique Costagliola, Axelle Dupont, France Mentré.

Data: Toni Alfaiate, Drifa Belhadi, Charles Burdet, Aline Dechanet, Christelle Delmas, Mouhamadou Diallo, Yakhara Diawara, Claire Fougerou-Leurent, Annabelle Metois, Priyanka Velou.

Radiology: Florence Ader, Claire Andrejak, Charles Burdet, Mickael Ohana, Nathan Peiffer-Smadja.

Virology: Maude Bouscambert, Charles Burdet, Alexandre Gaymard, Benjamin Leveau, Bruno Lina, France Mentré, Florence Morfin-Sherpa.

Pharmacology: Alpha Diallo, Minh Le, Benjamin Leveau, France Mentre, Nathan Peiffer-Smadja, Gilles Peytavin, Minh Le, Sarah Tubiana.

Immunology: Laurent Abel, Florence Ader, Alexandre Belot, Maude Bouscambert, Charles Burdet, Darragh Duffy, Véronique Fremeaux-Bacchi, François-Xavier Lescure, France Mentré, Nathan Peiffer-Smadja, David Smadja, Sophie Susen, Benjamin Terrier, Eric Vivier.

Genetics: Laurent Abel, Christelle Delmas, Benjamin Leveau, France Mentré, Sarah Tubiana.

Collection of biological human samples: Aline Dechanet, Christelle Delmas, Vinca Icard, Ouifiya Kalif, Benjamin Leveau, Bruno Lina, Florence Morfin-Sherpa, Valentine Piquard, Chaimae Saji, Céline Sakonda, Sarah Tubiana.

Safety matters: Drifa Belhadi, Aline Dechanet, Christelle Delmas, Alpha Diallo, Noemie Mercier, Laurence Moachon, Nathan Peiffer-Smadja, Vida Terzic.

Regulatory affairs, Monitoring and Financial management: Carole Cagnot, Sandrine Couffin-Cadiergues, Aline Dechanet, Christelle Delmas, Marina Dumousseaux, Hélène Esperou, Assia Ferrane, Claire Fougerou-Leurent, Delphine Lebrasseur, Soizic Le Mestre, Noemie Mercier, Marion Noret, Juliette Saillard, Vida Terzic.

Communication: Florence Ader, Sandrine Couffin-Cadiergues, Aline Dechanet, Christelle Delmas, Marina Dumousseaux, Hélène Esperou, Claire Fougerou-Leurent, France Mentré, Marion Noret, Nathan Peiffer-Smadja, Priscille Rivière, Juliette Saillard, Léa Surugue, Yazdan Yazdanpanah.

Drug supply: Johanna Guillon, Anne-Marie Taburet and Theradis PHARMA

Inserm-Transfert: Pascale Augé, Mireille Caralp, Nathalie Dugas, Florence Chung, Julia Lumbroso.

Clinical Research Unit CHU Bichat-Claude Bernard: Camille Couffignal.

Data hosting: the COVID-19 Partners Platform and Hervé Le Nagard.

We acknowledge the executive management staff of Inserm: Gilles Bloch (CEO), Claire Giry, Frédérique Le Saulnier, Jean-Christophe Hébert, Carine Delrieu.

We acknowledge the RENARCI network: Marion Noret, Albert Sotto and Pierre Tattevin.

We acknowledge the support of the Data management and Methodological centers:
ANRS (France Recherche Nord&Sud SIDA-HIV Hépatites), Paris: Ventzislava Petrov-Sanchez, Axel Levier, Chloé Birkle, Cécile Moins, Sandrine Gibowski, Elise Landry, Anaïs Le Goff, Laurence Moachon, Léna Wadouachi, Christelle Paul, Joséphine Balssa ; Inserm US19-Sc10/ U1018 CESP, Université Paris Saclay, Villejuif: Laurence Meyer, Maud Brossard, Asma Essat , Emmanuelle Netzer, Yoann Riault, Mathilde Ghislain ; CMG Inserm U1219, Bordeaux Population Health, Université de Bordeaux, Bordeaux: Linda Wittkop, Laetitia Moinot, Christine Betard ; Sorbonne Université, INSERM U1136, Institut Pierre Louis d'Épidémiologie et de Santé Publique (IPLESP), Paris: Lambert Assoumou, Lydie Béniguel, Michèle Génin et Lina Gouichiche ; CIC1414, Inserm, CHU Rennes: Bruno Laviolle, Claire Fougerou.

We acknowledge the monitoring management team: Beniguel Lydie, Betard Christine, Birklé Chloé, Brossard Maud, Cameli Charlotte, Caro Alain, Essat Asma, Génin Michèle, Ghislain Mathilde, Grubner Mélanie, Levier Axel, Moins Cécile, Netzer Emmanuelle, Ngo Um Tégué Marie-José, Pacaud Isabelle and Riault Yoann.

We acknowledge the Clinical research associates: *Autriche*: Esmaeilzadeh-Leithner Stephanie; *Belgium*:Van Sante Nathalie, Khalil Zineb; *France*: Ait Djoudi Malek, Antoine Lydie, Back Christelle, Bellonet Marcellin, Benlakhryfa Assia, Boudjoghra Nour, Bouhet Fabrice, Calmont Isabelle, Camara Sabine, Cherifi Asma, Collette Camille, De Lemos Alexandra, Diesel Marie, Donet Elodie, Douillet Marine, Dubois-Gache Caroline, Faillet Edith, Fanomezantsoa Volanantenaina, Flasquin Stéphanie, Fonooni Shervin, Fortes Lopes Euma, Gaudin Isabelle, Gautier Blandine, Gerome Quentin, Ghidi Marion, Ginoux Pauline, Gouichiche Lyna, Granjon Marie, Guerard Valérie, Haerrel Elina, Harpon Camille, Herbele Morgane, Lafuente Lorrie, Lagarde Dominique, Langlois Audrey, Le Breton Aude, Lejeune Stéphanie, Madiot Hend, Maniangou Bercelin, Marquis Eric, Martineau Anne-Sophie, Mejane Murielle, Mizejewski Béatrice, Mouanga Victoria, Mugnier Brigitte, Osman Issraa, Passageon Maxence, Pelkowski Manon, Pelonde-Erimée Véronique, Pintaric Christine, Pruvost Celina, Risse Brigitte, Rousseaux Justine, Schiano Christine, Seux Alexandra, Simon Marielle, Stupien Marie-Laure, Tallon Sophie, Tobia Jérémy, Traika Chaima, Tréhoux Solange, Verdier Alice, Wegang-Nzeufo Adele, Yatimi Rachida; *Luxembourg*: Montanes Gloria; Portugal: Madeira Catarina

We acknowledge the outstanding support of the Clinical Investigation Centers:
CIC1401, Inserm, CHU Bordeaux: Pierre-Olivier Girodet; CIC1403, Inserm, CHRU Lille: Dominique Deplanque; CIC1406, Inserm, CHU Grenoble: Jean-Luc Cracowski; CIC1407, Inserm, Hospices Civil de Lyon: Michel Ovize; CIC1408, Inserm, CHU Saint-Etienne: Bernard Tardy; CIC1411, Inserm, CHU Montpellier: Eric Renard; CIC1413, Inserm, CHU Nantes: Jean-Noël Trochu; CIC1414, Inserm, CHU Rennes: Bruno Laviolle; CIC1415, Inserm, CHU Tours: Wissam El Hage; CIC1417, Inserm, APHP Cochin: Odile Launay; CIC1418, Inserm, APHP HEGP: Jean-Sébastien Hulot; CIC1422,

Inserm, APHP Pitié-Salpétriêre: Jean-Christophe Corvol; CIC1424, Inserm, CH André Rosemon: Mathieu Nacher; CIC1424, Inserm, CHU de la Martinique: André Cabié; CIC1425, Inserm, APHP Bichat: Xavier Duval; CIC1430, Inserm, APHP Mondor: Philippe Le Corvoisier; CIC1431, Inserm, CHU Besançon: Emmanuel Haffen; CIC1432, Inserm, CHU Dijon: Marc Bardou; CIC1433, Inserm, CHU Nancy: Patrick Rossignol; CIC1434, Inserm, CHRU Strasbourg: Catherine Schmidt-Mutter; CIC1436, Inserm, CHU Toulouse: Olivier Rascol.

We acknowledge the pharmacists, the virologists and all others research staffs:
*Austria:* Bellmann Romuald, Ditlbacher Adelheid, Egle Alexander, Finkenstedt Armin, Greil Richard, Hauck Volker, Hirtenfelder Cornelia, Huemer Florian, Joannidis Michael, Klein Sebastian, Lamprecht Bernd, Langanger Margit, Lehner Georg, Morre Patrick, Peer Andreas, Radl Bianca, Rathmaier Sandra, Rinnerthaler Gabriel, Salzer Helmut, Schachner Michaela, Schlintl Verena, Sgonc Sabrina, Spitzlinger Karin, Van Sante Nathalie, Wolkersdorfer Martin, Zotter Klemens.
*Belgium:* Altdorfer Antoine, Barbezange Cyril, Belkhir Leila, Borgoens Philippe, Crine Michel, De Bievre Félix, De Greef Julien, Delhauteur Blaise, Deschampheleire Maud, Ducrot Jarode, Elmaouhab Kahina, Fagnoul Sorya, Fraipont Vincent, Gaillet Adeline, Geeraerts Els, Gouders Jean-François, Gougnard Thierry, Grimaldi David, Hallin Marie, Henry Angeline, Hites Maya, Jacobs Frédérique, Jacquet Sophie, Jonas Alexa, Kamun Mwinkeu Nina, Khalil Zineb, Kin Valérie, Lardinois Valentine, Lesenfant Valentine, Leunen Françoise, Marquez Kristel, Martiny Delphine, Mathonet Nathalie, Moerman Filip, Moonen Martial, Nardulli Sonia, Nguyen Thi, Nonis Romain, Opeind Grégory, Peters Annick, Raimo Michela, Rugwiro Robert, Scarniere Axelle, Serrano Gabriela, Vandenberghe Florence, Vandorpe Astrid, Verschelden Gil, Yombi Jean Cyr.
*France:* Abboud Philippe, Abel Sylvie, Abergel Hélène, Aboab Jérôme, Abou Arab Osama, Accart Bertrand, Accassat Sandrine, Achino Chloé, Adda Anne, Ader Florence, Adolle Anais, Adrait Arnaud, Agasse Carole, Aggoun Sabrina, Agranier Maxime, Ahmad Kais, Aissi Emmanuelle, Ait Hamou Zakaria, Ait Toufella Hafid, Alb Ional, Albaladejo Pierre, Albert Monique, Alfandari Serge, Alips Julie, Allaf Salima, Allain Jean-Sébastien, Allam Hayat, Allaouchiche Bernard, Allavena Clothilde, Alloui Ahmed-Chakib, Alvarez Muriel, Alviset Sophie, Alzina Camille, Amaru Priscilla, Ambert Audrey, Amiel Corinne, Amigues Sophie, Amirouche Melissa, Andrejak Claire, Andreu Pascal, Andriamanantena Dinaherisoa, Andry Mathilde, Anguel Nadia, Antakly-Hanon Yara, Aptel François, Aquereburu Quam, Arbadji Lila, Archinard Daphne, Arnoux Laetitia, Arrive Clémence, Asehnoune Karim, Assadi Maksud, Assaf Ady, Atchade-Thierry Enora, Atger Romain, Atik Nesrine, Attias Arie, Aubineau Magali, Aubrey Aurélie, Auchet Thomas, Audard Vincent, Audibert Gérard, Audoin Pierre, Aujoulat Olivier, Aussedat Marine, Auvray Christelle, Azria Philippe, Azzi Mathilde, Baba-Hamed Nabil, Baboi Loredana, Bachelard Antoine, Bachour Bassel, Bacque Camille , Bagate François, Bagouet Flora, Bailly Benoit, Bal Antonin, Balanca Baptiste, Baldeyrou Marion, Banisadr Firouze, Bansard Hélène, Bapteste Lionel, Barau Caroline, Barba Thomas, Bard Mathieu, Barosi Anna, Barrail

Tran Aurélie, Barraquier Isabelle, Barraud Damien, Barthelemi Laurie, Basille Damien, Bastides Frédéric, Baudin Aurélie, Baudry Thomas, Bayer Sophie, Beaucreux Charlotte, Bechoua Shaliha, Becker Agathe, Becker Guillaume, Becret Antoine, Bedague Damien, Beddi Sihame, Bedekovic Caroline, Belaube Nicolas, Belhadj Said, Belilita Ridha, Belkahla Sébile, Bellabes Ghezzoul, Bellegarde Justine, Bellenfant Florence, Beltramo Guillaume, Beltzung Gaétan, Ben Ghanem Sarah, Ben Hamou Adrien, Ben Salem Noussaier, Benaissa Mohamed Nassim, Benech Nicolas, Beneton Camille, Benezit François, Benis James, Benmammar Arezki, Benoist Florence, Benoit Philippe, Benrayana Raida, Berger Jean Luc, Beringuer Florence, Beringuer Hélène, Berkenou Jugurtha, Bernard Louis, Bernard Megguy, Bernez Julie, Bernigal Virginie, Bernon Silvia Juliana, Berns Marjory, Bert Arthur, Berthet Delteil Bryan, Bertholon Frédérique, Bertier Astrid, Berton Elodie, Bertrand Pierre-Jean, Bertrand Eliane, Besse Marie-Catherine, Beurton Alexandra, Bevilacqua Sybille, Bichard Iris, Bielefeld Philip, Bienvenu Boris, Biferi Pascal, Bigaillon Christine, Bigeard Bastien, Bignet Laurie, Billard Muriel, Billaud Geneviève, Birr Virginie, Bistoquet Marie, Bitker Laurent, Blanc François-Xavier, Blanc Myriam, Blanc Stéphane, Blanchard Julien, Blanchet Denis, Blanvillain Jean-Baptiste, Bleibtreu Alexandre, Blin Stephanie, Bloch Frédéric, Blondiaux Nicolas, Boczek Christelle, Boddaert Sophie, Bodet Contentin Laetitia, Bodonian Carole, Bohe Julien, Boibieux André, Boiteux Guillaume, Bokulu - Ikona Patricia, Bollens Diane, Bonnard Bénédicte, Bonniaud Philippe, Bonnin Carine, Bonnivard Michel, Bontemps Jérémy, Bontemps Julien, Bordier Emmanuel, Bortolotti Périne, Bosseray Annick, Botelho-Nevers Elisabeth, Bouadma Lila, Bouali Sanae, Bouaziz Valérie, Bouchaud Olivier, Bouchez Sabelline, Bouchier Baptiste, Boudjema Noel, Boueilh Anna, Bouffard Catherine, Bougault Quentin, Bougnaud Joanna, Bougon David, Bouiller Kévin, Bounes - Vardon Fanny, Bouras Marwan, Bourbon Johan, Bourdic Katia, Bourdiol Alexandre, Boureau Anne-Sophie, Bourel Emilie, Bourgault Olivier, Bourgoin Hélène, Bourgoin Nicolas, Bourhis Marion, Bourlet Thomas, Bourrassaud Philippine, Boursier Justine, Bouskila Nadège, Bousquet Aurore, Boussairi Abdelghani, Boussairi Abdel, Boussekey Nicolas, Boutoille David, Boutrou Mathilde, Bouvard Eric, Bouvet Romain, Bouvier Magali, Bouvier Stéphanie, Bouzat Pierre, Boyer Alexandre, Boyer Laurent, Boyer Philippe, Boyer-Besseyre Marielle, Bozon Fabienne, Braghini Nathalie, Brahmi Sabrina, Brasseur Maureen, Breaud Sophie, Bressollette Céline, Bretonniere Cédric, Brice Sylvie, Brichler Segolene, Brick Lysiane, Brie Elodie, Brimboeuf Marjorie, Brion Jean-Paul, Brochier Corinne, Brouard Pascale, Bruel Cédric, Brunel Anne-Sophie, Brunel Pauline, Brunet Aurélie, Buffet Alexandre, Bui Hoang-Nam, Buisson Marielle, Bulifon Sophie, Buscot Matthieu, Buzon Julie, Cabie André, Cabon Mathieu, Cabras Ornella, Cadoz Cyril, Cadranel Jacques, Cailhol Johann, Calvas Fabienne, Camara Boubou, Camou Fabrice, Campagne Julien, Campana Valentine, Canac Benoit, Candille Clara, Canellas Anthony, Canet Emmanuel, Cantais Emmanuel, Cao Albert, Cariou Alain, Carlier Nicolas, Carre Valérie, Carrier Cédric, Carrillon Romain, Carriot Michèle, Carteaux Guillaume, Carvalho Verlinde Muriel, Carvalho-Schneider Claudia, Casel Antoine, Casez-Brasseur Myriam, Cassou Nicolas, Castelain Sandrine, Castel-Kremer Elisabeth, Castet-Nicolas Audrey, Castex

Cédric, Catala Hélène, Catherine François-Xavier, Cattan Léa, Cattelan Jessie, Caturla Laetitia, Caulier Thomas, Causel Astrid, Cavailles Arnaud, Cavalli Zoé, Cavelot Sébastien, Cazanave Charles, Cazorla Céline, Cellerin Laurent, Cerana Sophie, Cerf Amélie, Cerro Valérie, Cestac Philippe, Chabartier Cyrille, Chabert Margaux, Chabert Paul, Chabot Jean-François, Chabrier Amélie, Chaigne Benjamin, Chalal Lynda, Chalal Solaya, Chalancon Alize, Chambrin-Lauvray Hélène, Champenois Isabelle, Champenois Vanessa, Chansombat Malikhone, Chanzy Bruno, Chaouat Ari, Chapelet Guillaume, Chapurlat Roland, Charbit Jonathan, Charbonnieras François, Charbonnier-Beaupel Fanny, Charles Carole, Charles Pierre-Emmanuel, Charmillon Alexandre, Charpentier Claire, Charpentier Julien, Charretier Pierre-Alain, Chartier Delphine, Chas Julie, Chatelain Anne, Chatelain Emeric, Chaudier Caroline, Chauvelot Louis, Chauvelot Pierre, Chavanet Pascal, Chazot Guillaume, Chebib Nader, Chenaf Samir, Chenard Laura, Chene Anne-Laure, Cheref Kahina, Cheret Antoine, Chesnel Chrystel, Chiappini Dominique, Chiarabini Thibault, Chiche Jean-Daniel, Chirio David, Chirouze Catherine, Choinier Pascaline, Chopin Marie Charlotte, Chouquer Renaud, Chumbi Flores René, Churaqui Françoise, Cisse Salif, Clairet Anne-Laure, Claude Fréderic, Clavere Gaelle, Clere-Jehl Raphaël, Clouzeau Benjamin, Cochennec Sabrina, Cohen Régis, Cohen Yves, Cohen-Aubart Fleur, Coinus Leo, Collarino Rocco, Collomb-Muret Roselyne, Colombe Barbara, Combes Isabelle, Commandeur Diane, Conan Pierre Louis, Conil Jean-Marie, Conrad Anne, Conrad Marie, Contenti Julie, Corbut Sylvie, Cordel Hugues, Cordier Charlotte, Cornavin Pauline, Cortial Delphine, Corvol Jean Christophe, Cossec Stéphanie, Costa Isabelle, Costedoat Chlumeau Nathalie, Cottin Vincent, Couderc Auriane, Counille Flavien, Cour Martin, Couraud Sébastien, Courjon Johan, Court Fortune Isabelle, Courte Guilhem, Courtiade Mahler Juliette, Cousson Joël, Coustilleres François, Couvelaere Marie, Couzigou Carine, Coyard Hélène, Cransac Amélie, Cravoisy Aurélie, Credoz Isabelle, Crockett Flora, Crognier Laure, Crosby Laura, Crutu Adrian, Cua Eric, Cuisinier Adrien, Da Silva Daniel, Daconceicao Olivia, Dacosta-Noble Philippine, Dagod Geoffrey, Daguenel-Nguyen Anne, Dailler Frédéric, Danielou Adeline, Danion François, Dargent Auguste, Darien Marie, Darmon Arthur, Darrason Marie, Dartevel Anaïs, Darwiche Walid, Dauchy Edith, Dauriat Gaelle, David Guillaume, Davier Marina, Davoine Claire, De Briel Dominique, De Castro Nathalie, De Champes De Saint Leger Christophe, De Chevigny Alix, De Kergaradec Laurence, De La Salle Sylvie, De Magalhaes Elodie, De Monte Anne, De Montmollin Etienne, De Parades Vincent, De Rougemont Alexis, De Rpost Nicolas, De Seissan De Marignan Donatien, Debard Alexa, Debord Sophie, Debruyne Geraud, Declety Philippe, Degoul Samuel, Degoutte Aurélien, Dei Svaldi Charlotte, Deiler Véronique, Delagreverie Héloïse, Delamare Catherine, Delannoy Matthieu, Delannoy Pierre-Yves, Delattre Charlotte, Delattre Claire, Delaunay Mickael, Delaveuve Eric, Delcourte Claire, Delecrin Virginie, Deleris Robin, Deletombe Baptiste, Delignette Marie-Charlotte, Dellamonica Jean, Delobel Pierre, Delorme Estelle, Delwarde Benjamin, Demaeght François, Demar Magalie, Demeret Sophie, Demeure Dominique, Demonchy Elisa, Demoule Alexandre, Denis Blandine, Dequin Pierre-François, Derdour Vincent, Deroche Claire, Derradji Ouda, Derridj Rekkeb Lydia, Dersigny Adrien, Dervieux Benjamin,

Descamps Diane, Descamps Agathe, Deschamvres Colin, Desclaux Arnaud, Desille-Dugast Mireille, Desmure Pierre-François, Desmurs-Clavel Hélène, Dessertaine Géraldine, Destras Grégory, Detoc Maëlle, Devigne Bertrand, Devilliers Herve, Devouassoux Gilles, Devun Flavien, Dewitte Antoine, Dewitte Marie, Dhenin Sandrine, Diata Anne-Claire, Didier Agnès, Didier Kevin, Didier Marion, Diehl Jean-Luc, Diesel Marie, Dieu Maeva, Dieye Thierno, Di-Filippo Sarah, Dignoire Charlotte, Digumber Marc, Dinot Vincent, Diop Aminata, Dirou Stéphanie, Djahieche Zahia, Djossou Félix, Doh Sébastien, D'oiron Roseline, Dolidon Samuel, Dollat Marion, Doncesco Régine, Dorard-Chabrier Marie, Dorez Didier, Dormoy Alexandre, Douache Ouarda, Doyen Denis, Dozier Aurélie, Drenou Bernard, Dres Martin, Dreyer Chantal, Droguete Gomes Nelson, Dubar Véronique, Dubois Jacqueline, Dubos-Lascu Georgeta, Dubost Clément, Ducoux Grégoire, Ducrocq Gregory, Dujon Rita, Duong Michel, Dupland Pierre, Dupont Clarisse, Dupont Herve, Duprez Mathilde, Dupuy Olivier, Duran Clara, Durand Michel, Durand Philippe, Durant Jacques, Duranteau Lise, Durel Aurélie, Durel Cécile-Audrey, Duret-Aupy Nina, Durieu Isabelle, Duruisseaux Michaël, Durupt Stéphane, Duthoit Juliette, Eberst Guillaume, Ehrmann Stephan, El Houfia Abderrahim, El Karoui Khalil, El Kfif Najat, Elmenkouri Ismahan, Elotmani Loubna, Emeyriat Nicolas, Emile Jean-François, Emmerich Joseph, Empis De Vendin Ombeline, Engelmann Ilka, Epaulard Olivier, Escaut Lélia, Escavy Annabelle, Eschapasse Emmanuel, Escudier Etienne, Escuret Vanessa, Esteve Clémentine, Esteve Laure, Evano Fabienne, Evrard Philippe, Eyvrard Frédéric, Fafi-Kremer Samira, Fajole Gaëlle, Falcon Dominique, Fallet Barthélemy, Fallet Vincent, Fangous Marie-Sarah, Fanton Annlyse, Fares Nabila, Farines Raffoul Magali, Fartoukh Muriel, Fathallah Nadia, Fattoum Jihane, Faure Alexandre, Faure Emmanuel, Faure Karine, Faurie-Grepon Antoine, Faurous William, Fave Gersende, Faviez Guillaume, Favre Laurine, Favrolt Nicolas, Fayol Antoine, Fayolle Pierre-Marie, Feldman Laurent, Ferdinand Marie Paule, Ferge Jean-Louis, Fernandez Céline, Ferre Virginie, Ferreira Luis, Ferrier Laurence, Ferroukhi Tania, Feuillet Soummer Séverine, Feurer Elodie, Feyeux Delphine, Ficko Cécile, Figueiredo Samy, Filizzola Clea, Fiorentino Arianna, Fischer Quentin, Fite Charlotte, Flandrin Jennifer, Flattres Duchaussoy Delphine, Flet Laurent, Florea Valentina, Florens Nans, Fofana Djeneba, Foignot Clément, Fois Elena, Folliet Laure, Fontaine Candice, Fontaine-Delaruelle Clara, Fontana Aurélie, Forestier Chantal, Fort Romain, Fouche Myrtille, Foulongne Vincent, Fourati Slim, Fournel Emmanuelle, Fournier Julien, Fourreau Alain, Franchineau Guillaume, Francois Catherine, Francois Nelly, Francony Gilles, Frelat Alais, Frentiu Emilia, Fresard Anne, Freymond Nathalie, Friggeri Arnaud, Frobert Emilie, Froidure Marie, Fuentes Axelle, Gabassi Audrey, Gaborit Benjamin, Gaci Rostane, Gagneux-Brunon Amandine, Gaide-Chevronnay Lucie, Gaillard Emmanuel, Galassi Yolande, Galerneau Louis Marie, Galitzky Monique, Gallien Sébastien, Gallot-Lavallee Jean, Gally Josette, Galtier Florence, Galvan Valérie, Gambier Nicolas, Ganansia Olivier, Gandia Peggy, Garnier Camille, Garnier Maud, Garnon Sophie, Garot Denis, Garret Charlotte, Garrier Ludivine, Garrigou Sonia, Gatel Julie, Gaudy-Graffin Catherine, Gautron Flavien, Gauzit Remy, Gavaud Ariane, Gay Samuel, Gaymard Alexandre, Gazengel Pierre, Gazoppi Loïc, Genet Roxana, George Karen, Georges

Bernard, Georges Hugues, Georges Marjolaine, Georges Valérie, Gerardin Alexandre, Gerber Athenaïs, Geri Guillaume, Geriniere Laurence, Gerlier Camille, Germain Johanne, Gerset Sandrine, Ghaleh Marzban Taghi Bijan, Ghionda Carine, Giambra Laure, Gibelin Cécilia, Gigandon Anne, Gilbert Marie, Gilg Morgane, Gil-Jardine Cédric, Gillet Benjamin, Ginoux Marylise, Girerd Barbara, Giroudot Lauriane, Giroux Fanny, Gissot Valérie, Glavnik Boris, Gniadek Claudine, Gobert Florent, Godard Elodie, Godefroy Nagisa, Godon Alexandre, Goehringer François, Gohier Sandrine, Gominet Marie, Gonfrier Géraldine, Gonzalez Jésus, Gormand Frédéric, Goueffic Yann, Goupil De Bouille Jeanne, Gourraud Maeva, Goury Antoine, Gousseff Marie, Grange Claire, Granger Agathe, Granier Sandra, Gras Emmanuelle, Gras Guillaume, Grateau Gilles, Gravet Alain, Gravier Simon, Greaud Cécile, Greigert Valentin, Griffonnet Jennifer, Grigoli Maxime, Grillot Nicolas, Grimaldi Florian, Grimbert Philippe, Grinea Alexandra, Gros Alexandre, Groul-Viaud Céline, Grumet Pierre, Gruson Didier, Guelminger Raphaële, Guennoun Samia, Guerci Philippe, Guerder Antoine, Guerin Corinne, Guerin Franck, Guerin Laurent, Gueye Aboubakar, Guffroy Aurélien, Guibert Nicolas, Guillard Daniel, Guillaud Constance, Guillaume Ariane, Guillaumot Anne, Guillet Henri, Guillet Laura, Guillon Antoine, Guinard Solene, Guinchard Samuel, Guion-Dusserre Marc, Guiot Anaïs, Guitteaud Karine, Gutfreund Estelle, Gyselinck Marie, Haby Céline, Hadmifar Rebecca, Hadzic Biljana, Hammal Soumeya, Hamon-Bouer Catherine, Hanna Jérôme, Hansmann Yves, Harrois Anatole, Hartard Cédric, Haudebourg Luc, Hautefeuille Serge, Hebrard Amélie, Hecketsweiler Stephane, Helms Julie, Hemon Claire, Henard Sandrine, Hengy Baptiste, Henry Carole, Hentzien Maxime, Herault Marie-Christine, Herbert Marine, Hermida Jean-Sylvain, Hermitte Alexia, Herpe Yves-Edouard, Herve Fabien, Himbert Berthe-Marie, Himbert Berthe-Marie, Himbert Dominique, Hinschberger Olivier, Hocquet Didier, Hodique Florine, Hostachy Camille, Hot Arnaud, Hourcastagnou Edith, Hourmant Yannick, Hublain Paméla, Huguenin Gabriel, Huguet Raphaelle, Huleux Thomas, Hulot Jean Sébastien, Huntzinger Julien, Hustache Mathieu Laurent, Hutt-Clauss Anne, Huynh Sophie, Hyvernat Hervé, Icard Vinca, Ichai Carole, Ilic Dejan, Imbert Audrey, Ingiliz Patrick, Ion Ciprian, Ionescu Carmen, Ioos Vincent, Isernia Valentina, Itani Oula, Ivanova Dessislava, Izopet Jacques, Jacquin Laurent, Jacquot Audrey, Jaffal Karim, Jaffre Sandrine, Jager Marion, Jais Xavier, Jambon Julie, Jandeaux Louise-Marie, Janssen Cécile, Jaquet Pierre, Jardin Suzcs Meriam, Jarrassier Audrey, Jarry Nathalie, Jaspart-Le Du Valentine, Jaubert Paul, Jaud-Fischer Coline, Jaureguiberry Stéphane, Jaureguy Maïté, Jaussaud Roland, Jay Lucille, Jean-Baptiste Sylvain, Jeanmaire Eliette, Jean-Marie Janick, Jean-Michel Vanessa, Jeanpetit Julien, Jegousso Quentin, Jegu Anne-Lise, Jesel Laurence, Jeudy Adeline, Jeulin Hélène, Jevnikar Mitja, Joffredo Emilie, Joher Nizar, Jolas Stéphanie, Jolivot Anne, Jondot Marie, Joseph Cédric, Josset Laurence, Jouan Fanny, Jouan Mathilde, Jouan Youenn, Jouault Anne-Sophie, Joubert Virginie, Journois Didier, Jouzeau Jean-Yves, Jozwiak Mathieu, Juarez Sophie, Juillard Laurent, Julia Zélie, Julien Gautier, Jullien Aurélie, Jullien Thomas, Jung Boris, Junquera Fernandez Ramon, Kabala Anna, Kaddari Fatima, Kaeuffer Charlotte, Kafif Ouifiya, Kahn Jean-Emmanuel, Kaiser Jean- Daniel, Karachi Carine, Kayser Damien,

Kearns Kevin, Kebir Amina, Kerneis Solen, Khatchatourian Lydie, Khebbeb Samy, Khenifer Safia, Khuu Alexandre, Kiakouama Lize, Kim Byeul-A, Kimmoun Antoine, Kirchhoffer Mylène, Klement Elise, Klouche Kada, Komla-Soukha Isabelle, Kortchinsky Laurent Talna, Koszutski Matthieu, Kotokpoyoukou Gaella, Kramer Laura, Krause Jessica, Kreitmann Louis, Kroemer Marie, Kummerlen Christine, Kuteifan Khaldoun, Labetoulle Marc, Labro Guylaine, Labruyere Marie, Lacasse Marion, Lachatre Marie, Lacombe Karine, Lafitte Blandine, Lafon Marie-Edith, Lafondesmurs Barthélémy, Lafont Anne-Gaëlle, Lagarrigue Sandra, Laine Fabrice, Laine Laurent, Laithier François-Xavier, Lalande Laure, Lamaignere Jean-Louis, Lamara Fariza, Lamarque Julie, Lambert-Dessoy Dorothée, Lambotte Olivier, Lamouche Wilquin Pauline, Lamrani Lilia, Lanier Sylvie, Lanoix Jean Philippe, Lanotte Philippe, Lansalot-Matras Pauline, Laplanche Sophie, Larcher Romaric, Larrat Charlotte, Lascarrou Jean-Baptiste, Lassel Ludovic, Lateb Mael, Laugel Elodie, Launay Odile, Laurant-Noel Violaine, Laviolle Bruno, Lazareth Isabelle, Le Bel Cecilia, Le Bihan Clément, Le Bras Morgane, Le Brazic Melaine, Le Cam Manuela, Le Cam Pierre, Le Corre Pascal, Le Donge Marie, Le Duff Florence, Le Febvre De Nailly Delphine, Le Gac Sylvie, Le Goas Carole, Le Grand Jennifer, Le Hir Isabelle, Le Lorc'h Erwan, Le Marec Julien, Le Marechal Marion, Le Moing Vincent, Le Noel Anne, Le Pavec Jérôme, Le Pluart Diane, Le Puil Séverine, Le Tacon Serge, Le Tiec Clotilde, Le Tulzo Yves, Le Turnier Paul, Le Vavasseur Benjamin, Lebacle Cédric, Lebeaux David, Lebouc Marie, Lebreton Thibault, Lecocq Maxime, Lecomte Raphael, Lecronier Marie, Lefebvre Bénédicte, Lefebvre Nicolas, Lefevre Benjamin, Lefevre Lucie, Lefort Carine, Lefrancois Remi, Lega Jean-Christophe, Legoff Antoine, Legrand Amélie, Legras Annick, Le-Guennec Loic, Lehmann Audrey, Lehoux Mélanie, Lehur Anne Claire-Marie, Leiterer Caroline, Lelaidier Rodolphe, Lelievre Jean-Daniel, Lelievre Lucie, Lemaignen Adrien, Lemaire Hélène, Lemarie Jérémie, Lemoel Fabien, Lemoine Kimberly, Lemonnier Alban, Lepape Alain, Lepiller Quentin, Lepilliez Marie-Laurence, Lermuzeaux Mathilde, Leroy Audrey, Leroy Olivier, Leroy Sylvie, Lescure François-Xavier, Lesens Olivier, Lessire Henry, Letaillandier Cyrielle, Letertre Paule, Leveau Benjamin, Levrard Mélanie, Levrat Albrice, Levraut Jacques, Levy Bruno, Levy Michael, Lhonneur Anne Sophie, Li Chung Tong Stéphanie, Liegeon Geoffroy, Lim Pascal, Limal Nicolas, Lion Sylvie, Llontop Claudia, Lloret Sophie, Lobbes Hervé, Locatelli-Sanchez Myriam, Loewert Sébastien, Lohmann Caroline, Lomazzi Sandra, Longere Benoit, Lorber Julien, Loriau Jérôme, Lortat Jacob Brice, Losser Marie-Reine, Louapre Céline, Loubet Paul, Louis Guillaume, Louni Françoise, Louyot Marie, Lucena E Silva Ibrantina, Lukaszewicz Anne-Claire, Luong Liem, Lupo Julien, Lutz Marie-France, Luyt Charles Edouard, Maakaroun Zoha, Macheda Gabriel, Madelon Aurélie, Mafunahenry Nomonde, Mahe Pierre-Joachim, Mahevas Matthieu, Mahiou Sabrina, Mahy Sophie, Maillet Mylène, Maillot François, Maizel Julien, Makinson Alain, Malapert Amélie, Malaplate Catherine, Malaquin Stéphanie, Malvy Denis, Mangin Coralie, Maniel Charlotte, Mankikian Stefan, Mansard Catherine, Mansuy Jean-Michel, Manuel Aldric, Marchand Adam Sylvain, Marchionni Sandra, Marcotte Guillaume, Marey Jonathan, Mariani Louise-Laure, Marie Manon, Marien Marie-Christine, Mariller Laure, Marin

Nathalie, Marino Maria Rosaria, Marlet Julien, Marois Clémence, Marquette Charles Hugo, Marquise Athéna, Martel Nora, Martin Aurélie, Martin Maëlle, Martin-Blondel Guillaume, Martin-Gaujard Géraldine, Martinot Martin, Marty Olivier, Marty-Quinternet Solène, Marzak Halim, Mathien Cyril, Mathiotte Emilie, Matignon Marie, Mattei Mathieu, Matton Lise, Maugars Diane, Maugendre Karine, Mauguin Pauline, Maurer Agnes, May Thierry, Mayala Kanda Clementine, Mayaux Julien, Maynard Marianne, Mazet Julien, Mazieres Julien, Mechai Fréderic, Medrano Chloé, Meguig Sayah, Mehawej Hanane, Mehdaoui Hossein, Meister Charlotte, Mekontso Dessap Armand, Melica Giovanna, Melin Marjolaine, Mellati Nouchan, Melzani Alessia, Memain Nathalie, Merazga Salima, Merchaoui Zied, Mercier Emmanuelle, Merdji Hamid, Mergeay-Fabre Mayka, Merlet Audrey, Mespoulh Pauline, Metz Carole, Metzger Catherine, Meunier Alexandre, Meybeck Agnès, Meyer Christian, Meynard Jean-Luc, Meyzenc Juliette, Meziani Ferhat, Mezidi Mehdi, Mhanna Laurent, Miailhe Arnaud-Félix, Miailhes Patrick, Miatello Jordi, Michel Constance, Michel Marc, Milia Julie, Milleron Olivier, Minier Marine, Minoves Mélanie, Mira Jean Paul, Mistral Thomas, Mohamed Shirine, Mohseni-Zadeh Mahsa, Molina Jean-Michel, Molinari Domitille, Mondain Véronique, Mongardon Nicolas, Monnet Xavier, Monnier Alexandra, Monnier Gwenaël, Monphile Maëlle, Monsel Gentiane, Montagne Samantha, Montange Damien, Montani David, Montes Brigitte, Montoya-Ferrer Ana, Montravers Philippe, Mootien Joy, Morand-Joubert Laurence, Morawiec Elise, Morel Jérôme, Morelot-Panzini Capucine, Morfin Sherpa Florence, Morice Sophie, Moriconi Mikael, Morin Jean, Morin Luc, Morisseau Marlene, Morlet Julien, Morquin David, Moulinet Thomas, Mounayar Anne-Laure, Mourad Jean-Jacques, Mourguet Morgane, Mouries Martin Suzanne, Mourvillier Bruno, Mouthon Luc, Moyet Julien, Mugnier Sophie, Muller Michel, Muller Géraldine, Muret Patrice, Murris-Espin Marlène, Mutter Catherine, Naccache Jean-Marc, Nadal Marine, Nadaud Julien, Naitammar Houria, Najioullah Fatiha, Nardi Annelyse, Nasr Mouin, Natella Pierre-André, Nau Antoine, Navellou Jean Christophe, Ndiaye Awa, Ndiaye Diama, Ndinga – Mondze Sandrine, Neau Didier, Nemlaghi Safaa, Nemoz Benjamin, Nerini Krystel, Neuillet Camille, Neviere Rémi, Ngo Stéphanie, Nguyen Caroline, Nguyen Lee, N'guyen Yohan, Nicaise Blaise, Nicolas Pierre, Nieszkowka Anna, Nkontcho Flaubert, Noel Nicolas, Noelsavina Elise, Not Adeline, Nourry Emilie, Nove-Josserand Raphaële, Novello Cécile, Novy Emmanuel, Nseir Saadalla, Occelli Céline, Odin Cécile, Ogoudjobi Stanislas, Oniskoff Guillaume, Oniszczuk Julie, Orain Jérémie, Orban Jean Christophe, Orquevaux Pauline, Osman David, Ottavy Grégoire, Ouali Fariza, Ouamara Nadia, Oudeville Pierre, Oudotte Adrien, Ouisse Carole, Oulehri Walid, Pacanowski Jérôme, Paccoud Olivier, Pagadoy Maïder, Pain Jean-Baptiste, Palacios Christia, Palich Romain, Palmieri Lola Jade, Panaget Sophie, Panel Nathalie, Panh Sylvain, Pansu Nathalie, Paquet Valérie, Parat Rachel, Parent Florence, Parmentier Laurie, Parrot Antoine, Parry Elodie, Parthonneau Jessica, Pasqualoni Elisa, Pasquier Jérémie, Patin Florence, Patoz Pierre, Patrat-Delon Solène, Patrier Juliette, Pavese Patricia, Pavot Arthur, Pavy Stephan, Payen Jean-François, Peigney Isabelle, Pellerin Sarah, Pelton Oriane, Pene Frédéric, Pere Hélène, Perez Lucas, Perez Pascale, Perez Pierre, Perez Yonatan, Pernet Antoine,

Pernot Corinne, Perny Jessica, Perpoint Thomas, Perraud Hélène, Perreau Pauline, Perrein Adeline, Perrier Jean-François, Perrin Caroline, Perrot Emilie, Perry Marielle, Pertek Jean-Pierre, Pertue Alrick, Petitjean Thierry, Pezzi Agnès, Phibel Audrey, Philibert Lorène, Philippart François, Philit François, Phung Bao, Pialoux Gilles, Piard Vanessa, Picard Yoann, Picard Julien, Picchi Hugo, Pierre Isabelle, Pierre Michael, Pierre-Francois Sandrine, Piet Emilie, Pillet Sylvie, Pilmis Benoit, Pilorge Catherine, Pimentel Ana, Pineau Sylvie, Pinto Luis, Piot Anne, Piot Juliette, Piquard Valentine, Piroth Lionel, Plantier Laurent, Poidevin Antoine, Poindron Vincent, Poingant Lucile, Poinsignon Lydie, Pointurier Valentin, Poisay Marine, Poissy Julien, Pollet Angélina, Pontier-Marchandise Sandrine, Popov Isabelle, Porche Chloé, Porte Lydie, Poteau Maxime, Potriquier Stéphane, Pouderoux Cécile, Poumaroux Bénédicte, Poupard Marie, Pourcher Valérie, Poutrel Solène, Pouvaret Anne, Pouzet Brigitte, Pozzetto Bruno, Pradier Maxime, Pradier Pauline, Praneuf Joanne, Pregny Anaële, Prevel Renaud, Previlon Miresta, Prevot Grégoire, Prevot Michel, Prin Sébastien, Priollet Pascal, Pronina Nina, Protin Caroline, Prouvost-Keller Bernard, Provent Marion, Provoost Judith, Pugliese Pascal, Quelven-Bertin Isabelle, Quenot Jean-Pierre, Quentin Joane, Quetant Sébastien, Quint Isabelle, Quintard Hervé, Rabaud Christian, Rabec Claudio, Rabel Constance, Rabouel Yannick, Racheline Anne, Raffi François, Rahmani Hassene, Rameau Pascaline, Ramiere Christophe, Ramonda Véronique, Ramstein Marie, Rapp Jocelyn, Raux Maxime, Ray Alice, Rayez Mélanie, Raymond Eric, Raynal Florence, Razat Bénédicte, Razazi Keyvan, Razurel Anais, Reach Pauline, Rebaudet Stanislas, Refes Amel, Reignier Jean, Renaud Yanina, Renet Sophie, Resiere Dabor, Retail Maryvonne, Revest Matthieu, Rey David, Reynaud Quitterie, Reynes Jacques, Rezaiguia Delclaux Saida, Rhem Delphine, Richard Jean-Christophe, Richard Lucie, Richard Marion, Richard Pascale, Richard-Colmant Gaelle, Rigault Guillaume, Rigault Marie-Christine, Rimmele Thomas, Risal Aurélie, Risso Karine, Ritzenthaler Thomas, Riu Béatrice, Robbins Ailsa, Robert Céline, Robin Sylvaine, Robineau Olivier, Rocaboy Camila, Rochas Sophie, Roche Mélanie, Rodallec Audrey, Rodrigues Maura, Roger Claire, Roger Pierre-Alexandre, Rohaut Benjamin, Rolland Muriel, Romaru Juliette, Ronat Véronique, Rondelot Grégory, Roque Afonso Anne Marie, Roquilly Antoine, Rosique Pauline, Rosolen Béatrice, Rouaud Agnès, Roucher Aude, Roucou Aurélie, Roudaut Jean-Baptiste, Rouget Antoine, Rougier Estelle, Rougon Marion, Rouis-Bouabdallah Rabia, Rousseau Antoine, Rousseau Dominique, Roussel Catherine, Rousson Jessica, Rouveix-Nordon Elisabeth, Roux Pauline, Roux Sandrine, Roux Virginie, Rouzeau Céline, Roy Mahaut, Roy Ema Renée Flore, Royer Blandine, Royer Hugo, Rozenberg Flore, Ruch Yvon, Ruiz Stéphanie, Rulle Sandrine, Saada Noémie, Sabatier Brigitte, Sabri Ouafa, Saccheri Clément, Sacco Emmanuelle, Sacher Huvelin Sylvie, Sadaoui Thiziri, Saidani Nadia, Saint Gilles Max, Saint-Martin Léa, Salah Samia, Salas Virginie, Saleh-Mghir Samira, Sallaberry Stéphane, Salmeron Sergio, Salmon Gandonniere Charlotte, Salmon Rousseau Arnaud, Salomon Alexis, Salvadori Alexandre, Sambin Sara, Sambou Aramatoulaye, Sans Nicolas, Sarhaoui Badria, Sarton Benjamin, Saubion Amandine, Sautereau Aurélie, Sauvaget Elisabeth, Savan Veaceslav, Savieras Aurélie, Sayre Naomi, Sazio Charline, Schaeffer Céline, Schieber Pachart Anne, Schlemmer Frédéric,

Schmit Jean-Luc, Schroeder Zoé, Schvoerer Evelyne, Schwebel Carole, Schwiertz Vérane, Seferian Andrei Hora, Seguin Amélie, Seguin Thierry, Senechal Agathe, Senneville Eric, Serres Audrey, Serruya Rebecca, Servettaz Amélie, Seve Pascal, Sgarioto Amandine, Si Albdelkader Hasni, Sigaud Florian, Siguier Martin, Silva Sifontes Stein, Simeon Véronique, Simon Marie, Sinnah Fabrice, Sirodot Michel, Sitbon Olivier, Slama Michel, Smagghe Florian, Solis Morgane, Sonneville Romain, Soriot-Thomas Sandrine, Sorli Caroline, Soucemarianadin Myriam, Soulie Cathia, Sounni Adame, Souquet Pierre-Jean, Soussi Malika, Souweine Bertrand, Sow Khaly, Spanjaard Maximilien, Spinar Sylvie, Stefan François, Stefic Karl, Steg Philippe Gabriel, Stehle Thomas, Stephan Robin, Studer Antoine, Suez-Panama Valérie, Tacquard Charles Ambroise, Tadie Jean-Marc, Tahon Elsa, Taieb Sarah, Taillard Silvine, Talarmin Jean-Philippe, Tamayo Marion, Tan Boun Kim, Tanaka Sébastien, Tardy Bernard, Tashk Parvine, Tateo Mariagrazia, Tattevin Pierre, Taverna Xavier-Jean, Tazarourte Karim, Tching-Sin Martine, Tebano Gianpiero, Terrier Anne, Terrier Benjamin, Terzi Nicolas, Terzian Zaven, Tetart Macha, Textoris Julien, Teyssou Elisa, Thalamas Claire, Thebault Elise, Thellier Damien, Thervet Eric, Thibault Vincent, Thibaut Pélagie, Thiebaut Léonie, Thiebaut Marie-Marthe, Thiemele Alaki, Thiery Guillaume, Thill Pauline, Thiolliere Fabrice, Thiriet Claire, Thivilier Carine, Thomas Guillemette, Thorey Camille, Timsit Jean-François, Tissiere Pierre, Tissot Elise, Tissot Marion, Tissot Noémie, Tolsma Violaine, Tonnelier Alexandre, Touati Saber, Touitou Irit, Trabattoni Eloise, Traclet Julie, Tran Marc, Tran Thanh Huy Christian, Tran-Dinh Alexy, Traore Lalla, Trehan Anne, Tribeau Stéphanie, Triffault Fillit Claire, Tritz Thomas, Trosch Clémentine, Trosini-Desert Valery, Trouba Cécile, Trousset Capucine, Trouve-Buisson Thibaut, Truche Anne Sophie, Tuaillon Edouard, Tubiana Sarah, Turcu Alin, Turlin Bruno, Turmel Jean-Marie, Ubertino Déborah, Ursenbach Axel, Ushmorova Daria, Valentin Simon, Valero Régine, Valette Martine, Valin Nadia, Vallod Yann, Vallois Dorothée, Valour Florent, Vandamme Sylvie, Vardanega Julie, Varenio Sophie, Vauchy Charline, Vauloup Felous Christelle, Vazel Marion, Veloudou Vanessa, Venard Véronique, Verdier Alice, Verhaeghe Richa Floriane, Verstuyft Céline, Vettoretti Lucie, Veyer David, Viailly Vinciane, Vialatte De Pemille Clémentine, Vidal Barbara, Vieille Magalie, Vignaud Françoise, Villeneuve Laurent, Villot Solènne, Vindrios William, Vinour Hélène, Virolle Sara, Virot Edouard, Vitrat Virginie, Voiriot Guillaume, Voisin Olivier, Volkov Lev, Von Hunolstein Jean Jacques, Voyat Julien, Vuotto Fanny, Waldmann Victor, Wallet Florent, Wasielewski Eric, Wattecamps Guihem, Wehrlen-Pugliese Sylvia, Weiss Nicolas, Welfringer Pascal, Wicky Paul Henry, Wilson Roza, Wyplosz Benjamin, Yahya Debza, Yannoutsos Alexandra, Yazdanpanah Yazdan, Yelnik Cécile, Yemi Pierre-Yves, Yonis Hodanne, Yzet Thierry, Zahaf Imane, Zambon Olivier, Zame Thierry, Zappella Nathalie, Zaraa Ines, Zarhrate-Ghoul Aïda, Zerbib Yoann, Zerbit Jérémie, Zeyons Floriane, Zinninger Noémie, Zmihi Sylia, Zouak Ayoub, Zouzou Ghaleb, Zunarelli Romain.

***Luxembourg****:* Andlauer-Vialette Emmanuelle, Aouali Nassera, Bart Nathalie, Berna Marc, Frantzen Françoise, Gaudillot Gregory, Graas Jérôme, Joachimowicz Annie, Otto Anne, Rashid Mohamed Ally, Reuter Jean, Skhiri Lamia, Staub Thérèse.

***Portugal:*** Ana Lucia Jesus, Beaumont Helena, Bento Tatiana, Braz Sandra, Candeias Carlos, Cardoso Patricia, Carneiro Raquel, Carrondo Ana, Cecilio Susana, Ceia Filipa Susana, Codea Vanessa, Costa Reis Renato, Couto Marta, Dardenne Murielle, Fernandes Ana Margarida, Fernandes Susana, Ferreira Ribeiro Joao Miguel, Figueiredo Sónia, Gaibino Nuno, Guimaraes Tiago, Jorge Ruben, Lamas Filipa, Leao Ana Cláudia, Lima Ana, Lopes Ana, Medeiros Fábio, Melo Carla, Miranda Fernando, Monteiro Patricia, Mota Cristiana, Neves Inês, Pinheiro Ricardo, Pinto André, Ramos Daniela, Ribeiro Raquel, Roncon-Albuquerque Roberto, Rosinhas José, Sarmento António, Silva Marisa, Soares Dias Fátima, Trigueiros Frederico.

We acknowledge the support of GILEAD, SANOFI, MERCK and ABBVIE which provided the study drugs.

This work received funding from several sources:

- *Europe*: European Union’s Horizon 2020 research and innovation programme
- *Austria*: AGMT gGmbH
- *Belgium*: Belgian Health Care Knowledge Centre; Fonds Erasme-COVID-ULB
- *France*: REACTing, a French multi-disciplinary collaborative network working on emerging infectious diseases; Ministry of Health; Paris Ile-de-France Region
- *Luxemburg*: European Regional Development Fund
- *Portugal*: Ministry of Health; Agency for Clinical Research and Biomedical Innovation
